# Plasma S1P links to hypertension and biomarkers of inflammation and cardiovascular disease – findings from a translational investigation

**DOI:** 10.1101/2020.12.07.20245415

**Authors:** Amra Jujic, Frank Matthes, Lotte Vanherle, Henning Petzka, Marju Orho-Melander, Peter M Nilsson, Martin Magnusson, Anja Meissner

## Abstract

Sphingosine-1-phosphate (S1P) is an important regulator of immune cell trafficking and vascular dysfunction contributing to the development and progression of overt hypertension. Although targeting S1P signaling revealed therapeutic potential in different experimental hypertension studies, validations of S1P-blood pressure (BP) associations in humans are lacking. In a translational approach, we explored the associations between plasma S1P and BP in a family-based study cohort (Malmö Offspring (MOS) study; N=1026), and in a longitudinally conducted murine hypertension cohort.

In MOS, linear multivariate regression analyses showed that plasma S1P associates with increased systolic BP (β=1.06, P=0.015). Study subjects with systolic BP ≥140 mmHg presented with significantly higher S1P plasma concentrations compared to subjects with BP <120 mmHg independent of age and sex. The S1P-BP association was validated in a murine model where plasma S1P increased with systolic BP (r=0.7018, R^2^=0.4925; P<0.0001). In a sub-sample of MOS (N=444), proteomic profiling for markers of inflammation, metabolism and cardiovascular disease using proximity Extension Assays revealed multiple significant S1P associations, some of them with marked sex-specificity. *In vitro* and *ex vivo* validation of identified S1P associations disclosed augmented expression of different vascular dysfunction and inflammation markers in response to S1P.

Our translational findings show a link between plasma S1P and systolic BP as well as several inflammation and cardiovascular disease markers and suggest S1P’s biomarker potential. This encourages further studies to investigate its predictive capacity for hypertensive disease or the therapeutic potential of its signaling axis.

## Introduction

Hypertension is the most common preventable risk factor for cardiovascular disease (CVD) and leading single contributor to all-cause mortality and disability worldwide.^1^ Blood pressure (BP) control is therefore considered the gold standard approach to reducing the proportion of population burden of BP-induced CVD. The existing link between BP and cardiovascular risk^1, 2^ highlights the need to intervene early. Following the results of the SPRINT study,^3^ both the American College of Cardiology (ACC)/American Heart Association (AHA) and the European Society of Cardiology/European Society of Hypertension (ESC/ESH) have gradually updated their advice on diagnosis and treatment of arterial hypertension.^4, 5^ The implementation of lower BP thresholds^5^ ultimately increases the prevalence of hypertension substantially,^6^ with a large young low-risk population who are recommended antihypertensive treatment according to the new guidelines. Undoubtedly, this would result in treatment of a substantial amount of people with no or little benefit. Thus, the incorporation of biomarkers to aid in the selection of patients that would actually respond to antihypertensive therapy (personalized medicine)^7^ and for the overall assessment of cardiovascular risk has been proposed.^8–11^ Selecting from the numerous inflammation, metabolism and CVD biomarkers that associate with a higher cardiovascular risk might inflict bias, hence the use of biomarkers with association to both BP and established CVD, inflammation and metabolism markers might be advantageous.

As major regulator of vascular functions, inflammatory and metabolic processes relevant to the pathology of hypertension and associated cardiovascular events the bioactive sphingophospholipid sphingosine-1-phosphate (S1P) might possess such biomarker potential.^12–18^ Besides holding cell type- and receptor-specific vasomodulator potency,^12, 14^ S1P signaling critically regulates important immune system functions,^19–22^ is involved in barrier function control,^23, 24^ and largely affects lipid and glucose metabolism.^25–27^ In experimental studies, augmented S1P signaling has been linked to several cardiovascular condition including stroke, heart failure, atherosclerosis and hypertension.^12, 13, 15, 28^ Small scale biomarker studies associated plasma S1P levels with impaired left ventricular ejection fraction (N=74) and all-cause mortality (N=210) in systolic heart failure patients.^29, 30^ Plasma S1P was further used to predict the severity of coronary artery atherosclerosis (N=59).^31^ With respect to hypertension, S1P’s role in disease development and propagation is mainly elusive,^13^ despite a few reports that indicate altered sphingolipid metabolism in different forms of experimental hypertension.^15, 28, 32–35^ To date, human cohort-based studies investigating associations between S1P plasma concentrations and BP are lacking. Therefore, this study explored the relationship between plasma S1P and BP in a large family-based cohort study, clinically validating results obtained in an experimental murine hypertension model. Additionally, evaluation of S1P associations with established CVD, inflammation and metabolism biomarkers using multiplex proteomic profiling and *in vitro* and *ex vivo* validation approaches were performed to study novel associations with established biomarkers relevant to hypertensive disease.

## Methods

### Human study population

The Malmö Offspring Study (MOS) is an on-going family-based cohort study.^36^ The study population consists of adult children (MOS-G2) and grandchildren (MOS-G3) of participants from the Malmö Diet and Cancer Study – Cardiovascular Cohort (MDCS-CC).^37^ Participants were recruited using official register information from the Swedish Tax Agency. Not understanding information in Swedish was the only exclusion criteria. The study population used in the current investigation consisted of children to MDCS-CC participants (N=1326). Subjects with missing data in any of the co-variates used in analyses of associations between S1P and BP were excluded, resulting in 1046 eligible subject (**Supplemental Figure S1**). Ethical approval has been obtained for MOS at the Regional Ethics Committee in Lund (Dnr. 2012/594).

#### Clinical assessment

Participants height (cm) and weight (kg) were measured in light indoor clothing. Resting BP (mmHg) was measured after 10 minutes of rest in the supine position using an automatic device (Omron). A mean of two readings with one minute apart was calculated.

Smoking (yes/no), anti-hypertensive treatment (AHT) and alcohol usage were self-reported in a web-based questionnaire. Diabetes was defined as either self-reported diabetes diagnosis, use of anti-diabetic medication or fasting plasma glucose levels ≥7.0 mmol/L at two separate visits at the research facility.

#### Laboratory assays

Blood samples were drawn after an overnight fast and analyzed for plasma glucose, creatinine and cystatin C at the Department of Clinical Chemistry, Malmö, which is part of a national standardization and quality control system. Plasma glucose was measured using the HemoCue Glucose System (HemoCue, Ängelholm, Sweden). Plasma creatinine was measured using an enzymatic colorimetric assay with an IDMS-traceable calibrator on the Hitachi Modular P analysis system (Roche, Switzerland). Plasma levels of cystatin C were determined by an automated particle-based immunoassay using Hitachi Modular P analysis system and reagents from DAKO (Dako A/S, Denmark). Relative estimated glomerular filtration rate (eGFR) was calculated as a mean of eGFR derived from creatinine and eGFR derived from cystatin C and reported as mL/min/1.73 m^2^.

#### Proteomic profiling

Proximity Extension Assay (PEA) technique was applied to analyze plasma levels of proteins (OLINK Bioscience, Uppsala, Sweden) in a sub-sample of the population (N=444; consecutive subjects from 6^th^ of March 2013 until 17^th^ of June 2015 with complete data on all examinations). Four OLINK panels were analyzed: (1) Inflammation Panel (<u>inflammation panel specifics</u>), (2) Metabolism Panel (<u>metabolism panel specifics</u>), (3) CVD II panel (<u>CVDII panel specifics</u>) and (4) CVD III panel (<u>CVDIII panel specifics</u>), all comprising 92 proteins each within different domains (N = 92×4 = 368). Proteins with ≥15% samples below limit of detection were excluded (N=20 for the Metabolism panel; N=27 for the Inflammation panel; N=7 for the CVDII panel; and N=4 for the CVDIII panel; **Supplemental Table S1**). Additionally, 11 proteins were overlapping between panels thus, only one of each marker was included in linear regression analyses (**Supplemental Table S2**), resulting in 299 proteins that were taken forward to analyses. Validation data and coefficients of variance for all panels is available on the OLINK homepage (http://www.olink.com).

### Animal Study

The investigations using research animals conform to the EU Directive 2010/63/EU for animal experiments and were conducted in accordance with European Animal Protection laws. All protocols were approved by the institutional ethical committee of Lund University (Dnr. 5.8.18/12637/2017). Commercially available male and female wild-type (WT) C57BL/6N mice were obtained from Taconic (Ejby, Denmark) and housed under standard 12h:12h light-dark cycle with access to food and water *ad libitum*. Mice with a body weight (BW) ≥ 25g were housed in groups of four to five mice per cage. Experimental groups were designed in a way to minimize stress for the animals and to guarantee maximal information using the lowest group size possible using a power calculation with Type I error α= 0.05 and Power of 1-β > 0.8 (80%) based on previous studies.^15^ Hypertension was induced using angiotensin II (AngII)-releasing osmotic pumps (Alzet-2006, AgnTho’s, Sweden). In brief, animals were anaesthetized with isoflurane (IsoFlo^®^ vet 100%, Sweden; 2.5% at 1.5L/min in room air) for subcutaneous pump implantation containing AngII (infusion rate 20ng/kg/min over four weeks). BP was measured bi-weekly in conscious mice using non-invasive tail-cuff plethysmography (CODA, EMKA, France), starting one week before pump implantation after a training period of 7 days. Weekly blood draws from the vena saphena were performed starting prior to pump implantation (=baseline) at seven days intervals and blood was collected in EDTA-coated tubes (Sarstedt, Germany). At termination, mice were anaesthetized (isoflurane 2.5% at 1.5L/min in room air) before euthanasia through cervical dislocation. Mesenteric arteries were dissected and immediately processed for RNA isolation using the Trizol method as per manufacturer’s instructions and subsequent quantitative real-time PCR.

### S1P plasma quantitation

S1P was extracted by mixing 10 µl of plasma with 90 µl of ice-cold methanol containing 22.2 nM S1P-D7 (Avanti Polar Lipids / Merck, Darmstadt, Germany) as internal standard. After incubation on ice for 30 min precipitate was removed by centrifugation (20,000 g for 10 min at 4 °C). The supernatant was analyzed by LC-MS/MS on a 6495 QQQ instrument (Agilent Technologies, Sweden) essentially as described.^38^ Extracts were separated on a 2.1 x 50 mm Acquity UPLC Peptide HSS T3 C18 column (Waters, Sweden) at a flow rate of 0.5 ml/min using eluents (A) water / 0.1 % formic acid / 1 mM ammonium formate, and (B) methanol / 0.1 % formic acid / 1 mM ammonium formate, with a gradient of 20 % A and 80 % B to 100 % B over 2 min, followed by 100 % B for 6 min. By multiple reaction monitoring, MS/MS transitions of *m/z* 380 to 264 (with 380 to 82 as qualifier) for S1P, and 387 to 271 (with 387 to 82 as qualifier) for S1P-D7, were measured. A calibration curve consisting of 7 concentrations in the range of 0.1 to 2.4 µM S1P in 4 % fatty acid-free BSA was generated in triplicates and measured during each session. Additionally, 3 plasma samples were used as quality control samples and measured during each session to ensure reproducibility.

### Ex vivo and in vitro testing

Endothelial cells of murine (bEND.3; ATCC^®^ CRL-2299^™^) and human (HMEC-1; ATCC^®^ CRL-3243^™^) origin were cultured in DMEM containing 10% FBS and 1% Penicillin-Streptomycin and MCDB131 supplemented with 10% FBS, 1% Penicillin-Streptomycin, 1% Glutamine (2mM), 1% NEAA, 1% sodium pyruvate, 0.1% Amphotericin B and 0.1% hEGF (10ng/ml), respectively. Cell cultures were maintained at 37°C with 5 % CO_2_ and split 1:4 at a seeding density of 10^6^ cells. Cells were incubated with 1 µM S1P or vehicle (4% fatty acid-free BSA) for 24 hrs prior to processing for RNA isolation using the Trizol method as per manufacturer’s instructions and subsequent quantitative real-time PCR. Mesenteric arteries isolated from male WT C57Bl/6N mice were cultured in DMEM (Gibco Life Technology, Sweden) containing 10% FBS (Gibco Life Technology, Sweden) and 1% Penicillin-Streptomycin (Sigma-Aldrich, Sweden) for 24 hrs in the presence of 1 µM S1P (Cayman Chemicals, BioNordika, Sweden) or vehicle (4% fatty acid-free BSA) prior to processing for RNA isolation and subsequent quantitative real-time PCR. Standard biochemical procedures were utilized for experiments involving reverse transcription polymerase chain reaction, quantitative PCR, and ELISA. Methodological details and primer sequences are provided in the data supplement.

### Statistical methods

#### Human study

Comparisons between BP groups were carried out using one-way ANOVA for continuous variables, and χ2 tests for binary variables. For linear regression analyses of associations between S1P and BP, S1P was z-transformed. For associations between S1P and systolic BP and diastolic BP, linear regression analyses were carried out unadjusted, adjusted for age and sex (*Model 1*), and further adjusted for BMI, eGFR, diabetes status, AHT, smoking status and alcohol usage (*Model 2*). Interaction analyses between age, eGFR, BMI, sex, and S1P were carried out in linear regression analyses using a moderator variable. For associations between S1P and systolic BP ≥120 mmHg; ≥130 mmHg; and ≥140 mmHg, logistic regressions were carried out unadjusted, adjusted for age and sex (*Model 1*), and further adjusted for BMI, eGFR, diabetes status, AHT, smoking status and alcohol usage (*Model 2*). For both linear and logistic regressions, P<0.05 was considered statistically significant.

For associations between proteins from the OLINK-panels and S1P, NPX (log2) data were used and S1P was log2-transformed accordingly as recommended for OLINK data (<u>Measuring protein biomarkers with OLINK</u>). Linear regression analyses were carried out unadjusted and adjusted for age and sex. In order to adjust for multiple testing, a Bonferroni-corrected P-value of 0.05/299 (P<1.67×10^-4^) was applied. Pearson’s correlation analyses were carried out with two-tailed significance testing and computation of exact correlation coefficients (Pearson’s r). With correlation matrices being calculated for four different panels, multiple comparison adjustment was performed using Bonferroni-corrected P-values for each correlation matrix individually; (P ≤7.81×10^-4^ = 0.05/64 for the inflammation panel, P ≤ 7.04×10^-4^ = 0.05/71 for the metabolism panel, P ≤ 5.95×10^-4^ = 0.05/84 for the CVDII panel and P≤ 5.75×10^-4^ = 0.05/87 for the CVDIII panel). The panel-wise correction for multiple comparison avoids a (possible incorrect) rejection of significant correlations when correction is instead based on strict P-values from the large amount of overall 299 markers. For methodological correctness, we always compare to the Bonferroni-corrected significance of all 299 markers used in the linear regression. Both, Pearson’s correlation and linear regression analyses were carried out using SPSS 26.0 (IBM).

#### Animal study

All data are expressed as mean ± SEM, where N is the number of animals. For assessment of differences in BP and plasma S1P levels over time, two-way repeated measure ANOVA followed by Tukey post hoc testing was performed. For comparison of multiple independent groups, parametric one-way ANOVA test was used, followed by Tukey post hoc test with exact P value computation. For comparison of two groups a two-tailed unpaired t-test was utilized. Differences were considered significant at error probabilities of P≤0.05. All statistical analyses were carried out using GraphPad software (Version 8.4.2).

#### Ex vivo and in vitro studies

All data are expressed as mean ± SEM, where N is the number of animals or independent *in vitro* experiments. For comparison of two groups a two-tailed unpaired t-test was utilized. Differences were considered significant at error probabilities of P ≤ 0.05. All statistical analyses were carried out using GraphPad software (Version 8.4.2).

#### Visualization of correlation data

The matrix of pairwise correlations was used to construct a network graph with nodes given by the individual markers and edges drawn between nodes if their corresponding markers show a correlation of absolute value r ≥ 0.3. Edge width was weighted by correlation in absolute values. The network was grouped into clusters using Gephi 0.9.2,^39^ which implements clustering by modularity taking edge weight into consideration.^40^ Resolution was chosen around the default value of 1.0 so that a maximal modularity score is obtained.

A separate t-SNE ^41^ analysis on the correlation data considers each variable as a data point with the correlation to all other variables as its features, defining a high-dimensional representation of the variables. We utilized the scikit-learn 0.23.1-implementation of t-SNE in Python (3.8.3) to learn a two-dimensional representation suitable for visualization, which reflects the relation of similar correlations encoded as proximity in the high-dimensional data.

#### Search strategy for biomarker associations with CVD and inflammatory disease

PubMed searches for relevant studies published within the past five years, using the following combination of Medical Subject Heading (MeSH) terms and text words, with no language limitations: “biomarker name”[MeSH Terms] OR “biomarker name”[All Fields] OR “biomarker name”[All Fields]) AND “CVD”[All Fields] for associations with CVD, and “biomarker name”[MeSH Terms] OR “biomarker name”[All Fields] OR “biomarker name”[All Fields]) AND “inflammatory disease”[All Fields] for associations with inflammatory disease. All observational and clinical studies as well as journal articles were included in the search.

## Results

### S1P plasma levels associate with systolic BP

S1P plasma levels were quantified in MOS-G2 participants with recorded systolic and diastolic BP information. Characteristics of the study population are presented in **Table 1**. There was a steady significant increase in S1P plasma levels as systolic BP values increased within cut offs defined as <120 mmHg; ≥120 mmHg; ≥130 mmHg, and ≥140 mmHg (P=0.024; **Table 1**). Subjects with systolic BP ≥140 mmHg were older and presented with significantly higher S1P plasma concentrations, higher BMI, higher diastolic BP, more frequent anti-hypertensive treatment and lower eGFR compared to subjects with BP <120 mmHg. Further, one-way ANOVA analysis revealed no statistically significant differences in plasma levels of S1P between the sexes (P=0.163).

**Table 1.**
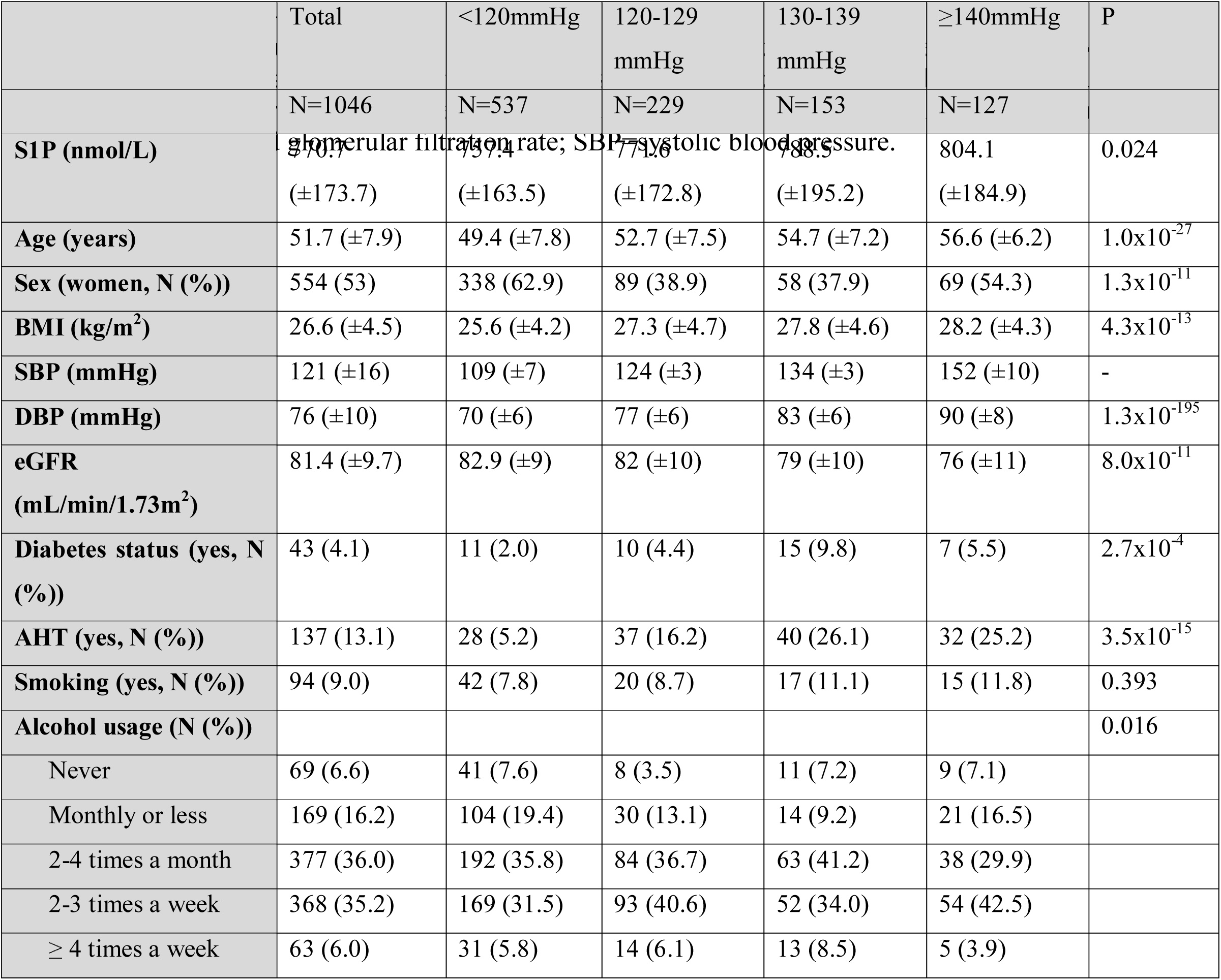
Characteristics of the study population.^1^

To explore associations between S1P and BP, linear regression analyses were performed revealing that each one standard deviation (1SD) increment of S1P was associated with increasing systolic BP but not diastolic BP when adjusted for age, sex, BMI, eGFR, diabetes status, anti-hypertensive treatment, smoking and alcohol usage (**Supplemental Table S3**). The relationships between S1P and BP were not mediated by age (P=0.975), sex (P=0.495), eGFR (P=0.980), or BMI (P=0.834) as determined by interaction analyses. Further, logistic regression analyses revealed that each 1SD increment of S1P was associated with systolic BP ≥120 mmHg, ≥130 mmHg and ≥140mmHg in adjusted models, with similar odds ratios (**Table 2**).

**Table 2.** S1P and systolic blood pressure associations.^2^

|  | ≥120 mmHg |  | ≥130 mmHg |  | ≥140 mmHg |  |
| --- | --- | --- | --- | --- | --- | --- |
|  | OR (CI95%) | P | OR (CI95%) | P | OR (CI95%) | P |
| **Unadjusted** |  |  |  |  |  |  |
| S1P | 1.17 (1.04-1.32) | 0.011 | 1.21 (1.06-1.38) | 0.005 | 1.23 (1.03-1.46) | 0.021 |
| **Model 1** |  |  |  |  |  |  |
| S1P | 1.14 (1.01-1.30) | 0.047 | 1.19 (1.03-1.37) | 0.016 | 1.21 (1.01-1.46) | 0.037 |
| Age | 1.10 (1.08-1.12) | $1.0 \times 10^{-21}$ | 1.10 (1.08-1.13) | $1.7 \times 10^{-19}$ | 1.11 (1.08-1.15) | $5.4 \times 10^{-13}$ |
| Sex | 0.41 (0.31-0.53) | $1.9 \times 10^{-11}$ | 0.66 (0.49-0.88) | 0.004 | 1.12 (0.76-1.65) | 0.563 |
| **Model 2** |  |  |  |  |  |  |
| S1P | 1.15 (1.01-1.32) | 0.046 | 1.21 (1.04-1.40) | 0.012 | 1.23 (1.02-1.48) | 0.034 |
| Age | 1.09 (1.06-1.11) | $9.0 \times 10^{-14}$ | 1.08 (1.06-1.11) | $6.7 \times 10^{-11}$ | 1.10 (1.06-1.13) | $6.2 \times 10^{-8}$ |
| Sex | 0.48 (0.36-0.64) | $2.6 \times 10^{-7}$ | 0.77 (0.57-1.05) | 0.098 | 1.20 (0.80-1.81) | 0.387 |
| BMI | 1.09 (1.06-1.13) | $1.0 \times 10^{-7}$ | 1.07 (1.04-1.11) | $6.3 \times 10^{-6}$ | 1.07 (1.03-1.12) | 0.002 |
| eGFR | 1.01 (0.99-1.02) | 0.643 | 0.99 (0.97-1.01) | 0.202 | 0.98 (0.96-1.00) | 0.048 |
| Diabetes status | 1.00 (0.46-2.20) | 0.993 | 0.86 (0.43-1.72) | 0.676 | 1.71 (0.69-4.23) | 0.249 |
| AHT | 2.89 (1.80-4.63) | $1.1 \times 10^{-5}$ | 2.12 (1.41-3.20) | $3.1 \times 10^{-4}$ | 1.47 (0.89-2.43) | 0.130 |
| Smoking | 1.25 (0.77-2.02) | 0.377 | 1.27 (0.77-2.11) | 0.348 | 1.14 (0.60-2.16) | 0.686 |
| Alcohol |  |  |  |  |  |  |
| Never | 1.09 (0.50-2.40) | 0.828 | 1.48 (0.63-3.44) | 0.366 | 2.11 (0.62-7.17) | 0.232 |
| Monthly or less | 0.98 (0.51-1.91) | 0.961 | 0.94 (0.45-1.97) | 0.877 | 2.24 (0.75-6.72) | 0.149 |
| 2-4 times a month | 1.43 (0.79-1.91) | 0.238 | 1.30 (0.68-2.47) | 0.433 | 1.74 (0.63-4.83) | 0.286 |
| 2-3 times a week | 1.53 (0.84-2.76) | 0.164 | 1.27 (0.67-2.41) | 0.466 | 2.54 (0.94-6.92) | 0.067 |
| ≥ 4 times a week | 0.92 (0.42-2.02) | 0.828 | 0.79 (0.41-1.50) | 0.466 | 0.57 (0.21-1.59) | 0.286 |

<sup>2</sup> Values are odds ratios (OR) and confidence intervals (CI95%) and significance values (P) for logistic regression analyses. AHT=anti-hypertensive treatment. BMI=body mass index; DBP=diastolic blood pressure; eGFR=estimated glomerular filtration rate; SBP=systolic blood pressure.

In order to verify a possible positive correlation between plasma S1P and systolic BP, we longitudinally assessed plasma S1P concentrations in a murine model of slowly developing hypertension (induced by a low-pressor dose of 20ng/kg/min AngII over the course of four weeks). In this model, BP steadily increased and established significance at four weeks after AngII pump implantation (**Figure 1A**). Similar to systolic BP, plasma S1P concentrations were significantly elevated compared to baseline after four weeks of AngII perfusion (**Figure 1B**). Thus, S1P plasma levels presented with a positive linear relationship to systolic BP in our murine model (r=0.7132, R^2^=0.5086; P<0.0001; **Figure 1C**).

**Figure 1.**
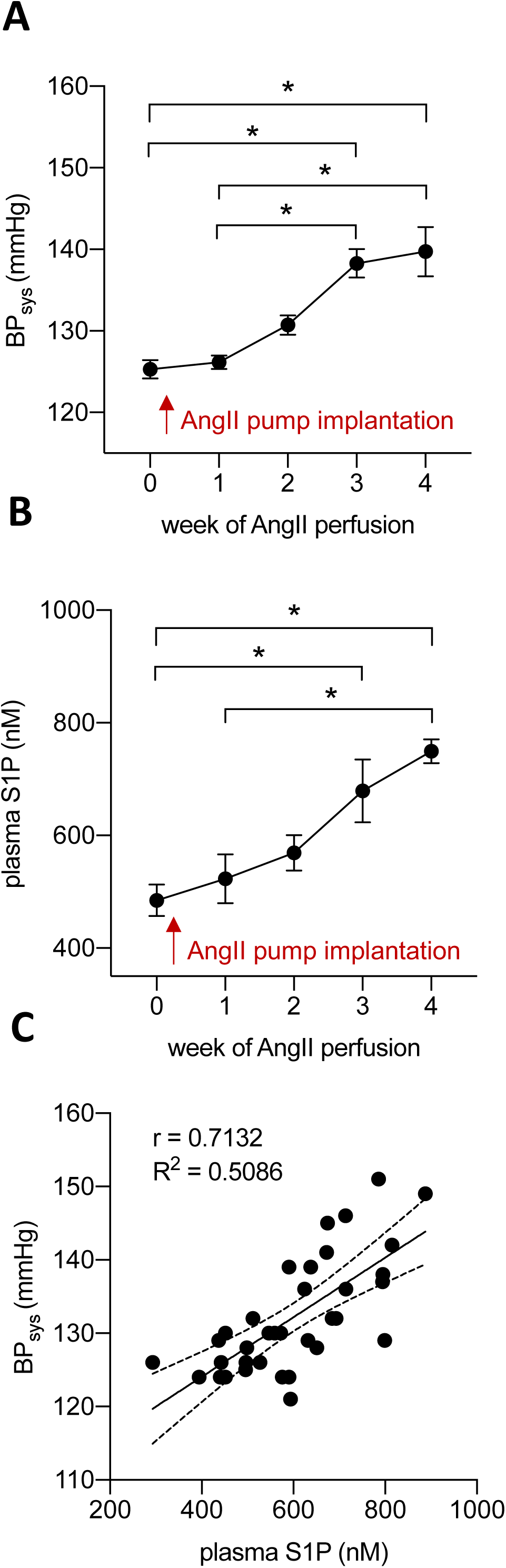
S1P plasma concentrations increase with systolic blood pressure in a mouse model of AngII-induced hypertension. **|A|** Longitudinal assessment of systolic BP (BP_sys_ in mmHg) in responses to a low pressor dose of AngII (20ng/kg/min) in WT mice over the course of four weeks. **|B|** Longitudinal plasma S1P quantification in WT mice developing hypertension over the course of four weeks. **|C|** Linear regression analysis of systolic BP (BP_sys_ in mmHg) and plasma S1P concentrations of WT mice developing hypertension in response to a low pressor dose of AngII (20ng/kg/min). In **A** and **B**, N=7; * denotes P≤0.05 compared to baseline after two-way repeated measure ANOVA followed by Dunnett’s post hoc testing. In **C**, N=7 per group and time point; linear regression analysis of association between plasma S1P and systolic BP levels (BP_sys_ in mmHg) with a calculated goodness of fit measurement (R^2^), Pearson’s r and exact P-value computation (P<0.0001).

### Proteomic profiling reveals significant associations between S1P plasma levels and inflammation, metabolism and CVD markers

As hypertension is a major modifiable risk factor for CVD,^12, 13, 15, 28^ relates to metabolic disease,^25–27^ and its pathophysiology strongly links to immune system activation and inflammation,^19,20–22^ we tested if S1P plasma levels associate to biomarkers of CVD, metabolism and inflammation in a subset of our study population (N=444). We categorized all markers with values above detection limit based on reported associations to cardiovascular or inflammatory diseases. **Figure 2** illustrates an overview of biomarkers detected in each OLINK panel, incorporating all significant S1P correlations after single comparison (grey dots) and multiple comparison testing (black dots). Pearson correlation analysis revealed 23 significant associations between plasma S1P and inflammation panel markers (panel-specific Bonferroni-correction), 21 of which were validated with Bonferroni-correction for testing all 299 markers (**Figure 2A** and **Supplemental Figure 2A**). Altogether 23 metabolism panel markers significantly correlated with plasma S1P (significant after both for panel specific Bonferroni-correction and for testing all 299 markers **Figure 2B** and **Supplemental Figure 2B**). CVDII and CVDIII panels presented with 22 and 24 proteins that correlated significantly with S1P levels (Bonferroni-corrected for individual panels), of which 21 and 16 remained significant after Bonferroni-correction for testing all 299 markers (**Figure 2C/D** and **Supplemental Figure S2C/D**). Overall, Pearson correlations revealed a total of 185 correlations with S1P levels across all panels, of which 92 and 81 remained significant after Bonferroni-correction for the number of panel-specific tests and all 299 tests, respectively (**Supplemental Figure S2)**. Correction for age and sex in the linear regression analyses resulted in the same number of significant associations for the individual biomarker panels. A full list of all unadjusted and age- and sex-adjusted associations in linear regression analyses between S1P and 299 proteins from all four panels (Inflammation, Metabolism, CVDII and CVDIII) is presented in **Supplemental Table S4**.

**Figure 2.**
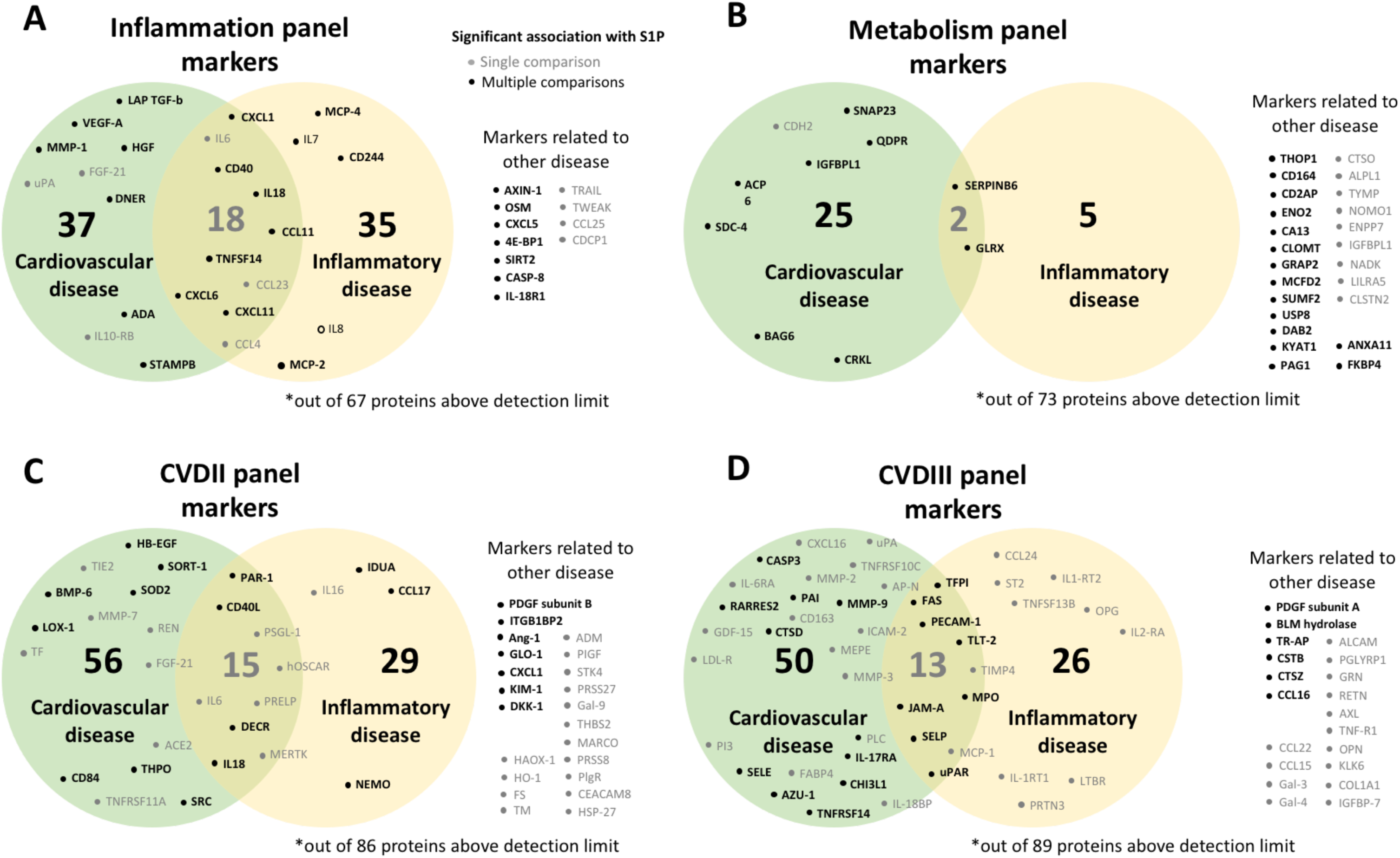
S1P significantly associates with various biomarkers in inflammation, metabolism and CVD OLINK panels. Overview of significant S1P correlations and associations amongst 299 proteins from |**A**| inflammation, |**B**| metabolism, |**C**| CDVII and |**D**| CDVIII OLINK panels. The numbers represent all panel markers with previously reported links to cardiovascular (green) or inflammatory disease (yellow), including overlapping markers in each panel. Significant S1P associations to these markers after Pearson’s correlation and linear regression analyses are presented for two-tailed test (grey dots), and for multiple comparisons with Bonferroni-correction of P-values (black dots). N=444

When applying a cut-off defined as r ≥ 0.3 or r ≤ - 0.3 to determine the strongest significant Pearson’s correlations^42, 43^ with plasma S1P in all four panels, a total of 29 markers were extracted that all revealed significant associations with S1P in linear regression analyses. 16 out of 29 strongly correlated markers belong to CVDII and CVDIII panels, 9 to the metabolism panel and 4 to the inflammation panel. **Figure 3** illustrates individual correlation networks of all four OLINK panels. Modularity clustering was applied to all networks as an algorithmic approach to detect communities of markers with strong pairwise correlations within a community and less frequent inter-correlations to markers of other communities. For the inflammation panel, marker clusters can be linked to T-cell homeostasis, immune cell homeostasis and chemotaxis (*cluster A*), neutrophil chemotaxis and angiogenesis (*cluster B*), or regulation of immune responses and association to CVD (*cluster C*), respectively (**Figure 3A**). S1P associations belong to *cluster B* and link to T-cell metabolism and apoptosis. Similarly, modularity clustering of metabolism panel markers detected 3 groups, separating markers associated to cell adhesion (*cluster A*), apoptosis and cellular stress response (*cluster B*), or cellular metabolism (*cluster C*). S1P associations link to markers representing *cluster B* (**Figure 3B**). CVDII panel clusters represent features linking to inflammation and metabolism associated to atherosclerosis, plaque development and cardiovascular events (*cluster A*), angiogenesis (*cluster B*), vascular dysfunction and inflammation (*cluster C*), the latter including all S1P associations of this panel (**Figure 3C**). For the CVDIII panel, marker clusters link to vascular dysfunction, remodeling and inflammation (*cluster A*), endothelial activation and inflammation (*cluster B*) or development of atherosclerosis and cardiovascular events (*cluster C*), respectively (**Figure 3D**). Strongest S1P associations belong to *cluster B*.

**Figure 3.**
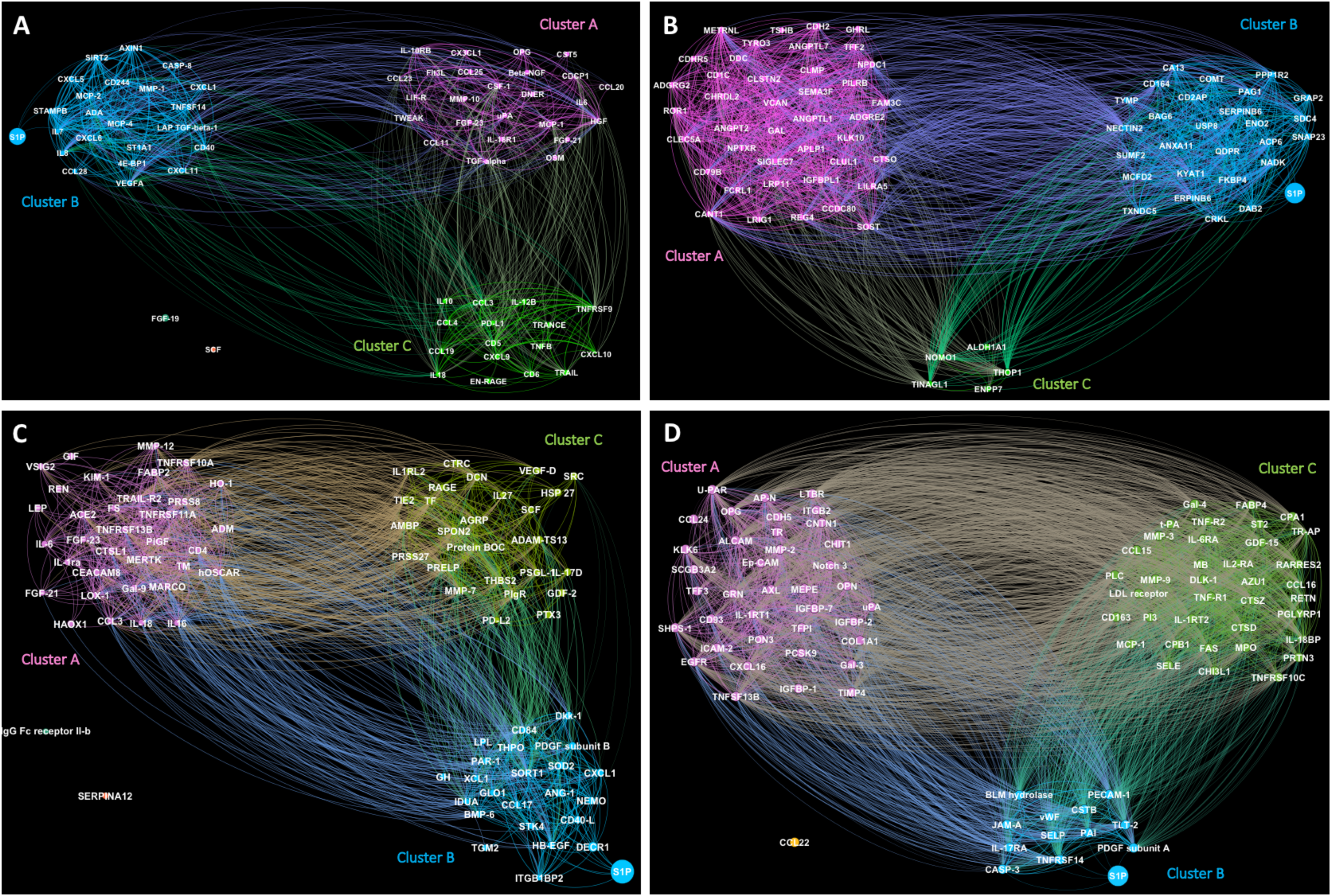
Correlation networks showing significant S1P correlations with biomarkers in the inflammation, metabolism and CVD OLINK panels. Modularity clustering applied to correlation networks for |**A**| inflammation panel (modularity = 0.271), |**B**| metabolism panel (modularity = 0.273), |**C**| CVDII panel (modularity = 0.189) and |**D**| CVDIII panel (modularity = 0.03) where each node represents a marker and edges between nodes a correlation factor of r ≥ 0.3 or r ≤ -0.3. Coloring according to cluster and edge width is weighted by correlation.

Modularity clustering applied to a network constructed from correlation factors of markers of all panels combined, separates markers into five distinct clusters, which can be linked to different processes involved in inflammation and CVD previously reported by clinical and pre-clinical studies (**Figure 4**). All significant S1P correlations are highlighted in red and associate to clusters 2, 3 and 4, representing markers linked to vascular inflammation, cell adhesion, immune cell metabolism and chemotaxis as well as atherosclerosis and cardiovascular events. To support the significance and stability of the found clusters, an application of a t-distributed stochastic neighbor embedding (t-SNE) visualizes the high dimensional correlation data in two dimensions, revealing an underlying data structure that can be compared to the clusters (**Supplemental Figure 3**). While the positioning of markers within a modularity cluster is arbitrary, the t-SNE locations of markers are meaningful where proximity is explained by similarity in correlations with other markers. The t-SNE arranges the markers into 9 visual groups that were linked to different processes involved in inflammation and CVD. When comparing tSNE groups with the modularity clusters, 81-92% marker overlap was observed between 5 tSNE groups (groups 1-3, 7, 9; **Supplemental Figure S3**) and clusters 1 to 5 (**Figure S4**). All S1P correlations are associated to groups 7 and 8, resembling marker groups linked to vascular inflammation, immune cell homeostasis and cell adhesion or endothelial inflammation and thrombosis, respectively. Marker overlap with cluster 3 (92%), where most markers can be linked to vascular inflammation, immune cell homeostasis and cell adhesion, largely confirms the meaningfulness of the modularity clusters. Correlation data used for all visualizations are presented as correlation matrices (**Supplemental Figure S4**).

**Figure 4.**
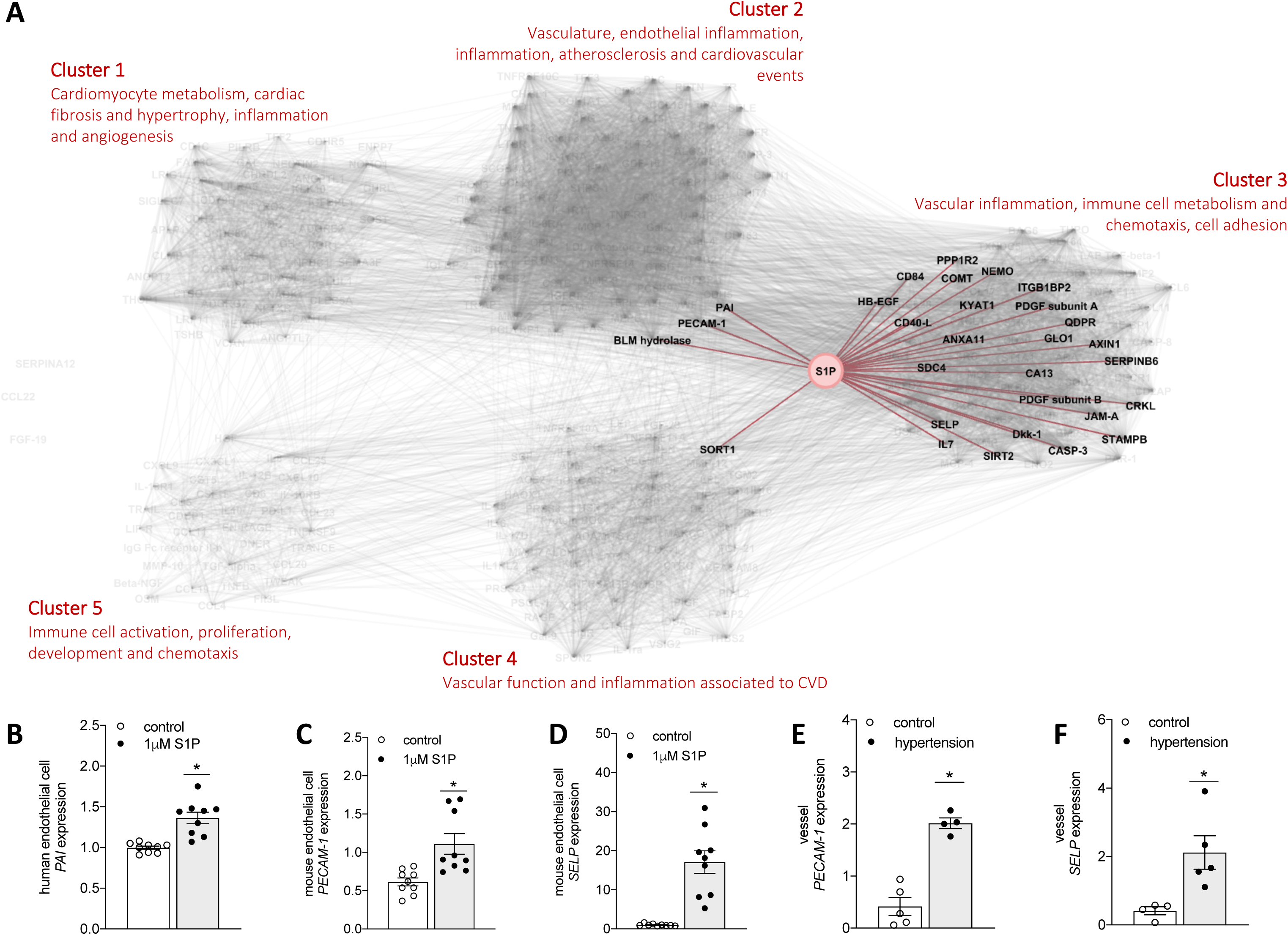
S1P increases various markers of inflammation and vascular dysfunction. **|A|** Modularity clustering applied to a network constructed from correlation factors of markers (r ≥ 0.3 or r ≤ -0.3) from inflammation, metabolism, CVDII and CVDIII OLINK panels combined. Significant S1P correlations highlighted in red. Clusters were categorized based on previously reported marker involvement in inflammation and CVD. **|B|** Augmentation of endothelial activation marker mRNA expression in human endothelial cells in response to 1 µM S1P (6hrs). **|C|** and **|D|** Augmentation of endothelial activation marker mRNA expression in murine endothelial cells in response to 1 µM S1P (12hrs). **|E|** and **|F|** Augmentation of endothelial activation marker mRNA expression in mesenteric arteries isolated from normotensive and hypertensive mice. In **A**, N=444. In **B-D**, N=3 per group in triplicates; * denotes P≤0.05 after single unpaired comparisons. In **E-F**, N=5 per group; * denotes P≤0.05 after single unpaired comparisons.

To experimentally test the effect of S1P on marker expression profiles, we performed *in vitro* and *ex vivo* experiments utilizing endothelial cells of murine and human origin that were treated with 1 µM S1P. As illustrated in **Figure 4B-D**, exposure to S1P significantly increased the expression of markers characteristic for angiogenesis and endothelial activation such as PAI (plasminogen activation inhibitor-1; **Figure 4B**), PECAM-1 (Platelet endothelial cell adhesion molecule 1; **Figure 4C**) and SELP (P-selectin; **Figure 4D**) in cultured human or mouse endothelial cells. All markers presented strongest positive associations with plasma S1P in our study cohort (r=0.409, β .409 for PAI-1; r=0.389, β=0.378 for PECAM-1 and r=0.383, β=0.384 for SELP). Remarkably, mesenteric arteries isolated from hypertensive mice presented with augmented PECAM-1 and SELP mRNA expression levels when compared to normotensive controls (**Figure 4E** and **Figure 4F**), strongly suggesting a link between plasma S1P and hypertension-associated vascular dysfunction and inflammation as systolic BP significantly correlated with plasma S1P levels in this model (see **Figure 1C**). Although cross-sectional associations do not allow conclusions on causality, several different markers increased expression after exposure to high S1P concentrations, including small resistance artery Ang-1 (angiopoietin 1), endothelial cell Casp3 (caspase 3), endothelial cell IL18 and monocytic cell CD40 (**Supplemental Figure S5**).

### Proteomic profiling discloses sex-specific associated effects of S1P for a subset of markers

As sex-dependent differences have been discussed for various S1P responses,^44^ we investigated the possibility of sex-specific S1P associations of all protein markers tested in our study. In single comparisons, we identified 66 significant sex dependent S1P correlations, out of which 32 showed stronger associations with male sex and 34 with female sex (**Supplemental Table S5**). After panel-specific correction for multiple comparisons, the majority of sex-specific differences were dictated by female sex (3 of 4 in the inflammation panel, 3 of 3 in the CVDII panel and 1 of 2 in the metabolism panel), while only one male-specific association remained significant after correction for multiple comparisons in the CVDIII panel. Eight out of ten sex-specific associations were confirmed with strict Bonferroni-correction for all 299 markers (**Table 3**). Amongst them IL18, which exerts apparent vascular and immune responses during hypertension and associates to adverse cardiovascular events, presented with a significantly stronger correlation in females (r = 0.278 and β = 0.538; P = 1.80×10^-5^) compared to males (r = 0.157 and β = 0.283; P = 0.021). Investigating the relation between IL18 and S1P in a controlled experimental setting revealed a similar sex-specific difference as evident by higher IL18 plasma levels in female hypertensive mice compared to their male counterparts (**Figure 5A**). Testing a linear relationship between IL18 and plasma S1P disclosed an extremely strong association in female mice (r = 0.9260, R^2^ = 0.8574; P < 0.0001) but not in male mice (r = 0.2175, R^2^ = 0.0473; P = 0.436) in our model (**Figure 5B**). Similar to observations in our human cohort, plasma S1P responses to BP increases did not differ sex-specifically (**Figure 5C**). In neither mice nor humans did IL18 associate to BP in either sex (male mice: r = 0.257, R^2^ = 0.066, P = 0.354; female mice r = -0.222, R^2^ = 0.491, P = 0.427; men: r = 0.070, P = 0.308 and β = 1.27; P = 0.308; women: r = 0.076, P = 0.257 and β = 2.90, P = 0.257).

**Figure 5.**
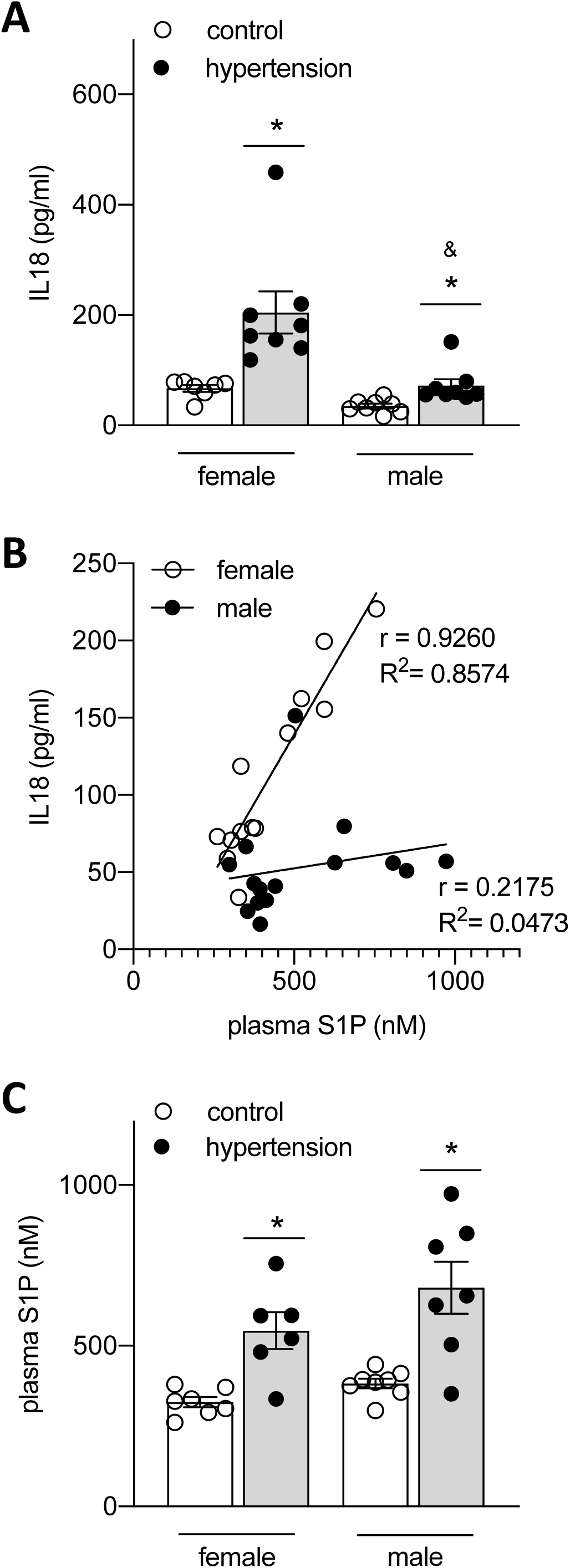
Plasma S1P-IL18 associations differ sex-dependently. **|A|** Plasma IL18 concentrations in normotensive and hypertensive female and male WT mice. **|B|** Linear regression showing associations between plasma S1P and IL18 in normotensive and hypertensive female and male WT mice. **|C|** Plasma S1P concentrations in normotensive and hypertensive female and male WT mice. In **A** and **C**, N=7-8 per group; * denotes P≤0.05 compared to same sex control and & denotes P≤0.05 compared to female hypertension group after one-way ANOVA followed by Tukey’s post hoc testing. In **B**, N=6-8 per group; Pearson correlation with goodness of fit and r computation; significance from zero was calculated for female (P<0.0001) but not male mice (P=0.4361).

**Table 3.** List of proteins from four proteomic panels that show significant sex-dependent associations with plasma S1P.^3^

| **Inflammation panel** | **Male (N=216)** |  |  | **Female (N=228)** |  |  |
| --- | --- | --- | --- | --- | --- | --- |
|  | **Pearson's r** | **β** | **P** | **Pearson's r** | **β** | **P** |
| **MMP-1** | 0.258 | 0.911 | 1.28x10<sup>-4</sup> | 0.202 | 0.663 | 0.002 |
| **CD40** | 0.200 | 0.345 | 0.003 | 0.254 | 0.382 | 1.02x10<sup>-4</sup> |
| **TWEAK** | 0.039 | 0.050 | 0.571 | 0.246 | 0.283 | 1.78x10<sup>-4</sup> |
| **CD244** | 0.160 | 0.235 | 0.019 | 0.245 | 0.308 | 1.84x10<sup>-4</sup> |
| **Metabolism panel** |  |  |  |  |  |  |
|  | **Pearson's r** | **β** | **P** | **Pearson's r** | **β** | **P** |
| **ENO2** | 0.250 | 0.601 | 1.97x10<sup>-4</sup> | 0.316 | 0.677 | 7.97x10<sup>-7</sup> |
| **SUMF2** | 0.284 | 0.654 | 2.10x10<sup>-5</sup> | 0.196 | 0.435 | 0.003 |
| **CVDII panel** |  |  |  |  |  |  |
|  | **Pearson's r** | **β** | **P** | **Pearson's r** | **β** | **P** |
| **LOX-1** | 0.194 | 0.305 | 0.004 | 0.250 | 0.423 | 1.21x10<sup>-4</sup> |
| **IL18** | 0.157 | 0.283 | 0.021 | 0.278 | 0.538 | 1.80x10<sup>-5</sup> |
| **GLO1** | 0.241 | 0.619 | 3.41x10<sup>-4</sup> | 0.389 | 0.929 | 8.93x10<sup>-10</sup> |
| **CVDIII panel** |  |  |  |  |  |  |
|  | **Pearson's r** | **β** | **P** | **Pearson's r** | **β** | **P** |
| **CTSD** | 0.276 | 0.567 | 4.0x10<sup>-5</sup> | 0.189 | 0.271 | 0.009 |

<sup>3</sup> The association-determining sex highlighted in orange. Multiple comparison adjustment was performed using Bonferroni-corrected P-values of 0.05/299. For the inflammation panel, two additional significant sex-specific association with plasma S1P are found significant according to panel-specific Bonferroni correction of 0.05/64 ( $P \leq 7.81 \times 10^{-4}$ ) that are highlighted in yellow. N=444.

## Discussion

For the first time, we show significant association between plasma S1P and systolic BP levels in a large human cohort study (N=1046) and validate these findings in a longitudinally conducted pre-clinical murine hypertension study where S1P plasma levels positively correlate with systolic BP. Additionally, our data provide first evidence of significant associations between plasma S1P and multiple cardiovascular, inflammation and metabolism biomarkers assessed by proteomic profiling of 444 MOS participants. Some of these markers present with marked sex-specific interactions. Taken together, our translational findings strongly suggest a link between S1P plasma concentrations and systolic BP and encourage further prospective studies that investigate S1P’s potential as therapeutic target or risk marker in hypertensive disease.

The bioactive phosphosphingolipid S1P plays a critical role in both the vascular and the immune system and has proven involvement in experimental hypertension.^12, 15, 45, 46^ Specifically, one of the enzyme generating S1P, sphingosine kinase 2 (SphK2), presented as a key player in mediating plasma S1P responses to AngII, immune cell egress from secondary lymphoid tissue and vascular dysfunction and thereby, contributing to the development of overt hypertension.^15^ Thus far, human-based studies investigating S1P associations with BP were lacking. Our study is the first to reveal associations between increasing S1P plasma levels and systolic BP cut-offs of <120mmHg, ≥120mmHg, ≥130mmHg, and ≥140mmHg. Concurrently, the effect size of S1P association with systolic BP in humans is comparable to those of well-established hypertension-associated biomarkers such as renin, soluble interleukin 1 receptor-like 1 (ST2) and IL6. Together, these promising results call for further prospective studies to investigate S1P’s potential suitability as predictive marker for identifying subjects at high risk of incident hypertension in the population. As the first study showing significant associations between plasma S1P and systolic BP in a large population cohort, it is reassuring that the mean S1P plasma levels measured in our human study population validate those recently published for a study group comprising 174 healthy participants with a median age of 45.5 years,^47^ which further substantiates our findings. Similar to our study, neither S1P plasma nor serum levels were affected by sex, age or BMI.^47, 48^

Previously, S1P was assigned with marker capacity for coronary artery disease.^49^ The authors speculated whether S1P might be a marker for inflammatory processes associated with coronary disease or whether sphingolipids are ischemic markers. Here, we provide first evidence of significant associations between plasma S1P and multiple CVD, inflammation and metabolism markers in a relatively young study population with very few cardiovascular incidents, suggesting that S1P might associate to pathogenesis (e.g., inflammatory processes) rather than endpoints. Modularity clustering that detects communities of markers with strong pairwise correlations within a community arranged the strongest S1P associations into marker communities linked to vascular inflammation, immune cell homeostasis, cell adhesion, endothelial inflammation and thrombosis. PECAM-1 as multi-functional vascular cell adhesion molecule,^50^ presented with positive associations to S1P in humans. A tSNE visualization, where locations of markers are meaningful as proximity is explained by similarity in correlations with other markers, grouped PECAM-1 in close proximity to other makers related to endothelial inflammation, thrombosis, atherosclerosis and cardiovascular events, confirming S1P-PECAM-1 involvement in endothelial damage and deregulation of inflammatory responses at the blood-endothelial interface.^51^ In light of this, we observe augmentation of PECAM-1 expression in response to excess exogenous S1P *in vitro*, *in vivo* and *ex vivo*, suggesting a pivotal relationship that supports previous studies demonstrating PECAM-1 as a downstream target for S1P signaling in human endothelial cells.^52^ Together with our findings that show elevated resistance artery PECAM-1 expression and increased plasma S1P in hypertensive mice, these data are supportive of potential contributions of S1P-PECAM-1 deregulations in hypertensive disease. Similarly, SELP that plays an essential role in the recruitment of leukocytes during inflammation^53^ presented with strong S1P associations in our human cohort and was upregulated in human and murine endothelial cells following exogenous S1P treatment. In our visualization approaches, SELP clusters together with markers of vascular inflammation and cell adhesion and in close proximity to PECAM-1, respectively. This is particularly intriguing since soluble SELP levels are elevated among healthy women at risk for future vascular events^54^ thus, encouraging the consideration of S1P as therapeutic target or risk marker for hypertensive disease.

A potential sex-specificity for S1P plasma concentrations is controversially discussed with equally many studies suggesting sex-dependent differences as those reporting similar S1P plasma levels between the sexes.^44, 47, 48, 55–58^ A previous study investigating S1P levels, sex and pre/postmenopausal status in 108 healthy participants demonstrated a significant association between plasma S1P and estrogen, with higher S1P plasma concentration in pre-menopausal than post-menopausal women.^44^ Evaluating a much smaller study population, the authors assessed plasma S1P levels in individuals in an age range between 16 to 55 years as compared to our study population with a narrow age range, 51.7 (±7.9) years, and presumably a predominance of post-menopausal women. Nonetheless, the herein observed sex-independence of S1P concentrations, specifically in respect to disease, requires verification in even larger cohorts.

In light of known sex-specific differences in BP control,^59^ experimental studies highlighted the critical role of inflammation (i.e. regulatory T-cells and T helper 17 cells) in mediating such differences.^60^ Despite the relative sex-independence of plasma S1P in our study, some of the herein identified S1P associations with proteomic markers presented with sex-specific interactions. Amongst them IL18, which has been suggested as independent predictor of cardiovascular events in subjects with metabolic syndrome^61^ and chronic kidney disease,^62^ is involved in destabilization of atherosclerotic plaques^63^ and was linked to essential hypertension.^64^ The herein observed sex-specific differences in S1P-IL18 associations are interesting as they are independent of BP (i.e. correlation and linear regression analyses revealed no significant associations between IL18 and BP in either sex). In respect to BP, only one experimental study thus far reported significantly higher IL18 mRNA expression in the renal cortex of female spontaneously hypertensive rats compared to age-matched males^65^ despite 10-15 mmHg higher BP in male compared to female rats.^66^ This is particularly interesting since BP in men significantly differs from women in our study cohort (125 mmHg (±14) and 119 (±17) for men and women respectively; P=7.5×10^-10^). Similar female-driven increases in the renal cortex of spontaneously hypertensive rats were observed for CD40 expression,^65^ which positively associates with S1P plasma levels in women but not men in our study. Thus, our findings show that BP-independent sex-specific S1P associations to different inflammation, metabolism and CVD markers exist, suggestive of sex-specific differences in immune and vascular responses during hypertension or its development with potential significance for cardiovascular health that might only be unveiled when directly comparing to S1P plasma levels.

### Perspective

For the first time, we show a clear relationship between increments in plasma S1P and increasing systolic BP in a large human population cohort. Together with the observed associations between plasma S1P and multiple CVD, inflammation and metabolism biomarkers, this is suggestive of S1P’s biomarker potential. Our findings strongly encourage further prospective studies to investigate S1P’s capacity as predictive marker for hypertensive disease.

### Strengths and limitations

By studying a general population and adjusting for risk factors for hypertension, we demonstrated that S1P plasma level are associated with increased systolic BP in humans. Further, we substantiated these findings in an animal model of hypertension. However, since BP regulation is multifactorial, drawing conclusions about associations should be done with caution. The present study shares limitations common to all cross-sectional studies as no conclusion of causality can be drawn. Moreover, the MOS study was carried out in subjects of mainly Swedish descent, and the conclusions may not be generalizable to all populations or ethnicities. Also, since the MOS study is ongoing and participants still fairly young, we were unable to investigate possible association of S1P with incident disease (e.g., incident hypertension or CVD).

The use of mass spectrometry to determine S1P concentrations may be seen as limitation due to the specific tissue preparation requirements and the limited availability of this technological platform. Thus far, however, no feasible alternative for processing large sample sizes with such accuracy is available.

Proteomic profiling of four different OLINK panels (inflammation, metabolism, CVDII and CVDIII) was performed using inter-plate controls for markers within the individual panels but no standardization was performed to compare between the different panels. By analyzing markers that appeared in more than one panel, a high degree of similarity (i.e. correlation of the same marker between two panels) was observed for inflammation, metabolism and CVDII panels (r ∼ 0.8). In a few cases of the CVDIII panel, however, lower correlation coefficients were observed (r ∼ 0.45 for two extreme cases). We identified a technical variation associated to the OLINK data as reason for low correlation between duplicate markers, which has been reported before^67^ and which became substantial for some markers with a biological variance below 0.4 NPX. Thus, correlations to markers outside the panels, in particular S1P, should not be expected to markedly surpass correlation factors of duplicate markers and some associations may remain undetected due to the commonly used cut-off r > 0.3. Additionally, we observed a small bias on correlation factors within a panel. This induces a higher chance for markers within one panel to fall into the same cluster when combining all four panels. To mitigate the role of the technical error in the cluster formations, we performed additional individual panel clustering for data interpretation.

## Disclosure

None.

## Data availability

The data underlying this article are available in the article and in its online supplementary material. Study population-specific data that support the findings of this study are available upon reasonable request from the Steering Committee of Malmö Diet and Cancer study by contacting its chair, Professor Olle Melander, but restrictions apply to the availability of these data, which were used under license for the current study, and so are not publicly available due to ethical and legal restrictions related to the Swedish Biobanks in Medical Care Act (2002:297) and the Personal Data Act (1998:204).

## Author contribution

Conception [AM], design of the work [AM], data acquisition [FM, LV], data analysis [AJ, HP, FM, LV], data interpretation [AM], manuscript drafting [AJ, HP, MM, AM], manuscript revision [AJ, FM, LV, HP, MM, MOM, PMN], funding acquisition [AM, MM, MOM, PMN].

## Data Availability

The data underlying this article are available in the article and in its online supplementary material. Study population-specific data that support the findings of this study are available upon reasonable request from the Steering Committee of Malmö Diet and Cancer study by contacting its chair, Professor Olle Melander, but restrictions apply to the availability of these data, which were used under license for the current study, and so are not publicly available due to ethical and legal restrictions related to the Swedish Biobanks in Medical Care Act (2002:297) and the Personal Data Act (1998:204).

## Novelty and Significance

1. What Is New?

- First study to assess plasma S1P levels in a large family-based study cohort (N=1046)
- Proteomic profiling that revealed associations between plasma S1P and multiple cardiovascular, inflammation and metabolism biomarkers
2. What Is Relevant?

- S1P levels increase with systolic BP
- BP but not plasma S1P differs sex-specifically
- Sex-specific S1P associations with several biomarkers, including IL18 exist but are mostly BP-independent
3. **Summary:** For the first time, it is shown that plasma S1P levels increase with systolic BP in a large relatively young and healthy population cohort where it also associates with several cardiovascular, inflammation and metabolism biomarkers.

## References

1. Forouzanfar MH, Liu P, Roth GA, Ng M, Biryukov S, Marczak L, Alexander L, Estep K, Hassen Abate K, Akinyemiju TF, Ali R, Alvis-Guzman N, Azzopardi P, Banerjee A, Barnighausen T, Basu A, Bekele T, Bennett DA, Biadgilign S, Catala-Lopez F, Feigin VL, Fernandes JC, Fischer F, Gebru AA, Gona P, Gupta R, Hankey GJ, Jonas JB, Judd SE, Khang YH, Khosravi A, Kim YJ, Kimokoti RW, Kokubo Y, Kolte D, Lopez A, Lotufo PA, Malekzadeh R, Melaku YA, Mensah GA, Misganaw A, Mokdad AH, Moran AE, Nawaz H, Neal B, Ngalesoni FN, Ohkubo T, Pourmalek F, Rafay A, Rai RK, Rojas-Rueda D, Sampson UK, Santos IS, Sawhney M, Schutte AE, Sepanlou SG, Shifa GT, Shiue I, Tedla BA, Thrift AG, Tonelli M, Truelsen T, Tsilimparis N, Ukwaja KN, Uthman OA, Vasankari T, Venketasubramanian N, Vlassov VV, Vos T, Westerman R, Yan LL, Yano Y, Yonemoto N, Zaki ME, Murray CJ. Global burden of hypertension and systolic blood pressure of at least 110 to 115 mm hg, 1990-2015. JAMA. 2017;317:165–182

2. Lewington S, Clarke R, Qizilbash N, Peto R, Collins R, Prospective Studies C. Age-specific relevance of usual blood pressure to vascular mortality: A meta-analysis of individual data for one million adults in 61 prospective studies. Lancet. 2002;360:1903–1913

3. Group SR, Wright JT, Jr., Williamson JD, Whelton PK, Snyder JK, Sink KM, Rocco MV, Reboussin DM, Rahman M, Oparil S, Lewis CE, Kimmel PL, Johnson KC, Goff DC, Jr., Fine LJ, Cutler JA, Cushman WC, Cheung AK, Ambrosius WT. A randomized trial of intensive versus standard blood-pressure control. N Engl J Med. 2015;373:2103–2116

4. Williams B, Mancia G, Spiering W, Agabiti Rosei E, Azizi M, Burnier M, Clement DL, Coca A, de Simone G, Dominiczak A, Kahan T, Mahfoud F, Redon J, Ruilope L, Zanchetti A, Kerins M, Kjeldsen SE, Kreutz R, Laurent S, Lip GYH, McManus R, Narkiewicz K, Ruschitzka F, Schmieder RE, Shlyakhto E, Tsioufis C, Aboyans V, Desormais I, Group ESCSD. 2018 esc/esh guidelines for the management of arterial hypertension. Eur Heart J. 2018;39:3021–3104

5. Whelton PK, Carey RM, Aronow WS, Casey DE, Jr., Collins KJ, Dennison Himmelfarb C, DePalma SM, Gidding S, Jamerson KA, Jones DW, MacLaughlin EJ, Muntner P, Ovbiagele B, Smith SC, Jr., Spencer CC, Stafford RS, Taler SJ, Thomas RJ, Williams KA, Sr., Williamson JD, Wright JT, Jr. 2017 acc/aha/aapa/abc/acpm/ags/apha/ash/aspc/nma/pcna guideline for the prevention, detection, evaluation, and management of high blood pressure in adults: A report of the american college of cardiology/american heart association task force on clinical practice guidelines. J Am Coll Cardiol. 2018;71:e127–e248

6. Muntner P, Carey RM, Gidding S, Jones DW, Taler SJ, Wright JT, Jr., Whelton PK. Potential us population impact of the 2017 acc/aha high blood pressure guideline. Circulation. 2018;137:109–118

7. Byrd JB. Personalized medicine and treatment approaches in hypertension: Current perspectives. Integr Blood Press C. 2016;9:59–66

8. Pokharel Y, Sun W, de Lemos JA, Taffet GE, Virani SS, Ndumele CE, Mosley TH, Hoogeveen RC, Coresh J, Wright JD, Heiss G, Boerwinkle EA, Bozkurt B, Solomon SD, Ballantyne CM, Nambi V. High-sensitivity troponin t and cardiovascular events in systolic blood pressure categories: Atherosclerosis risk in communities study. Hypertension. 2015;65:78–84

9. Uddin SMI, Mirbolouk M, Kianoush S, Orimoloye OA, Dardari Z, Whelton SP, Miedema MD, Nasir K, Rumberger JA, Shaw LJ, Berman DS, Budoff MJ, McEvoy JW, Matsushita K, Blaha MJ, Graham G. Role of coronary artery calcium for stratifying cardiovascular risk in adults with hypertension. Hypertension. 2019;73:983–989

10. Patel KV, Pandey A, de Lemos JA. Conceptual framework for addressing residual atherosclerotic cardiovascular disease risk in the era of precision medicine. Circulation. 2018;137:2551–2553

11. Pandey A, Patel KV, Vongpatanasin W, Ayers C, Berry JD, Mentz RJ, Blaha MJ, McEvoy JW, Muntner P, Vaduganathan M, Correa A, Butler J, Shimbo D, Nambi V, deFilippi C, Seliger SL, Ballantyne CM, Selvin E, de Lemos JA, Joshi PH. Incorporation of biomarkers into risk assessment for allocation of antihypertensive medication according to the 2017 acc/aha high blood pressure guideline: A pooled cohort analysis. Circulation. 2019;140:2076–2088

12. Cantalupo A, Gargiulo A, Dautaj E, Liu C, Zhang Y, Hla T, Di Lorenzo A. S1pr1 (sphingosine-1-phosphate receptor 1) signaling regulates blood flow and pressure. Hypertension. 2017;70:426–434

13. Don-Doncow N, Zhang Y, Matuskova H, Meissner A. The emerging alliance of sphingosine-1-phosphate signalling and immune cells: From basic mechanisms to implications in hypertension. Br J Pharmacol. 2019;176:1989–2001

14. Hoefer J, Azam MA, Kroetsch JT, Leong-Poi H, Momen MA, Voigtlaender-Bolz J, Scherer EQ, Meissner A, Bolz SS, Husain M. Sphingosine-1-phosphate-dependent activation of p38 mapk maintains elevated peripheral resistance in heart failure through increased myogenic vasoconstriction. Circ Res. 2010;107:923–933

15. Meissner A, Miro F, Jimenez-Altayo F, Jurado A, Vila E, Planas AM. Sphingosine-1-phosphate signalling-a key player in the pathogenesis of angiotensin ii-induced hypertension. Cardiovasc Res. 2017;113:123–133

16. Rivera J, Proia RL, Olivera A. The alliance of sphingosine-1-phosphate and its receptors in immunity. Nature reviews. Immunology. 2008;8:753–763

17. Yagi K, Lidington D, Wan H, Fares JC, Meissner A, Sumiyoshi M, Ai J, Foltz WD, Nedospasov SA, Offermanns S, Nagahiro S, Macdonald RL, Bolz SS. Therapeutically targeting tumor necrosis factor-alpha/sphingosine-1-phosphate signaling corrects myogenic reactivity in subarachnoid hemorrhage. Stroke. 2015;46:2260–2270

18. Yang J, Noyan-Ashraf MH, Meissner A, Voigtlaender-Bolz J, Kroetsch JT, Foltz W, Jaffray D, Kapoor A, Momen A, Heximer SP, Zhang H, van Eede M, Henkelman RM, Matthews SG, Lidington D, Husain M, Bolz SS. Proximal cerebral arteries develop myogenic responsiveness in heart failure via tumor necrosis factor-alpha-dependent activation of sphingosine-1-phosphate signaling. Circulation. 2012;126:196–206

19. Matloubian M, Lo CG, Cinamon G, Lesneski MJ, Xu Y, Brinkmann V, Allende ML, Proia RL, Cyster JG. Lymphocyte egress from thymus and peripheral lymphoid organs is dependent on s1p receptor 1. Nature. 2004;427:355–360

20. Eken A, Duhen R, Singh AK, Fry M, Buckner JH, Kita M, Bettelli E, Oukka M. S1p1 deletion differentially affects th17 and regulatory t cells. Sci Rep. 2017;7:12905

21. Garris CS, Wu L, Acharya S, Arac A, Blaho VA, Huang Y, Moon BS, Axtell RC, Ho PP, Steinberg GK, Lewis DB, Sobel RA, Han DK, Steinman L, Snyder MP, Hla T, Han MH. Defective sphingosine 1-phosphate receptor 1 (s1p1) phosphorylation exacerbates th17-mediated autoimmune neuroinflammation. Nature immunology. 2013;14:1166–1172

22. Liao JJ, Huang MC, Goetzl EJ. Cutting edge: Alternative signaling of th17 cell development by sphingosine 1-phosphate. Journal of immunology. 2007;178:5425–5428

23. Wilkerson BA, Argraves KM. The role of sphingosine-1-phosphate in endothelial barrier function. Biochim Biophys Acta. 2014;1841:1403–1412

24. Chen T, Lin R, Jin S, Chen R, Xue H, Ye H, Huang Z. The sphingosine-1-phosphate/sphingosine-1-phosphate receptor 2 axis in intestinal epithelial cells regulates intestinal barrier function during intestinal epithelial cells-cd4+t-cell interactions. Cell Physiol Biochem. 2018;48:1188–1200

25. Fox TE, Bewley MC, Unrath KA, Pedersen MM, Anderson RE, Jung DY, Jefferson LS, Kim JK, Bronson SK, Flanagan JM, Kester M. Circulating sphingolipid biomarkers in models of type 1 diabetes. J Lipid Res. 2011;52:509–517

26. Frej C, Andersson A, Larsson B, Guo LJ, Norstrom E, Happonen KE, Dahlback B. Quantification of sphingosine 1-phosphate by validated lc-ms/ms method revealing strong correlation with apolipoprotein m in plasma but not in serum due to platelet activation during blood coagulation. Anal Bioanal Chem. 2015;407:8533–8542

27. Fayyaz S, Henkel J, Japtok L, Kramer S, Damm G, Seehofer D, Puschel GP, Kleuser B. Involvement of sphingosine 1-phosphate in palmitate-induced insulin resistance of hepatocytes via the s1p2 receptor subtype. Diabetologia. 2014;57:373–382

28. Spijkers LJ, van den Akker RF, Janssen BJ, Debets JJ, De Mey JG, Stroes ES, van den Born BJ, Wijesinghe DS, Chalfant CE, MacAleese L, Eijkel GB, Heeren RM, Alewijnse AE, Peters SL. Hypertension is associated with marked alterations in sphingolipid biology: A potential role for ceramide. PloS one. 2011;6:e21817

29. Xue Y, Jiang W, Ma Q, Wang X, Jia P, Li Q, Chen S, Song B, Wang Y, Zhang J, Liu J, Yang G, Lin Y, Liu J, Wei L, Dong C, Li H, Xie Z, Bai L, Ma A. U-shaped association between plasma sphingosine-1-phosphate levels and mortality in patients with chronic systolic heart failure: A prospective cohort study. Lipids Health Dis. 2020;19:125

30. Polzin A, Piayda K, Keul P, Dannenberg L, Mohring A, Graler M, Zeus T, Kelm M, Levkau B. Plasma sphingosine-1-phosphate concentrations are associated with systolic heart failure in patients with ischemic heart disease. J Mol Cell Cardiol. 2017;110:35–37

31. Sattler K, Lehmann I, Graler M, Brocker-Preuss M, Erbel R, Heusch G, Levkau B. Hdl-bound sphingosine 1-phosphate (s1p) predicts the severity of coronary artery atherosclerosis. Cell Physiol Biochem. 2014;34:172–184

32. Chen J, Tang H, Sysol JR, Moreno-Vinasco L, Shioura KM, Chen T, Gorshkova I, Wang L, Huang LS, Usatyuk PV, Sammani S, Zhou G, Raj JU, Garcia JG, Berdyshev E, Yuan JX, Natarajan V, Machado RF. The sphingosine kinase 1/sphingosine-1-phosphate pathway in pulmonary arterial hypertension. American journal of respiratory and critical care medicine. 2014;190:1032–1043

33. Zhao YD, Chu L, Lin K, Granton E, Yin L, Peng J, Hsin M, Wu L, Yu A, Waddell T, Keshavjee S, Granton J, de Perrot M. A biochemical approach to understand the pathogenesis of advanced pulmonary arterial hypertension: Metabolomic profiles of arginine, sphingosine-1-phosphate, and heme of human lung. PloS one. 2015;10:e0134958

34. Gairhe S, Joshi SR, Bastola MM, McLendon JM, Oka M, Fagan KA, McMurtry IF. Sphingosine-1-phosphate is involved in the occlusive arteriopathy of pulmonary arterial hypertension. Pulmonary circulation. 2016;6:369–380

35. Tabeling C, Yu H, Wang L, Ranke H, Goldenberg NM, Zabini D, Noe E, Krauszman A, Gutbier B, Yin J, Schaefer M, Arenz C, Hocke AC, Suttorp N, Proia RL, Witzenrath M, Kuebler WM. Cftr and sphingolipids mediate hypoxic pulmonary vasoconstriction. Proceedings of the National Academy of Sciences of the United States of America. 2015;112:E1614–1623

36. Brunkwall L, Jonsson D, Ericson U, Hellstrand S, Kennback C, Ostling G, Jujic A, Melander O, Engstrom G, Nilsson J, Ohlsson B, Klinge B, Orho-Melander M, Persson M, Nilsson PM. The malmo offspring study (mos): Design, methods and first results. Eur J Epidemiol. 2020

37. Rosvall M, Janzon L, Berglund G, Engström G, Hedblad B. Incident coronary events and case fatality in relation to common carotid intima-media thickness. Journal of Internal Medicine. 2005;257:430–437

38. Chipeaux C, de Person M, Burguet N, Billette de Villemeur T, Rose C, Belmatoug N, Heron S, Le Van Kim C, Franco MC, 2017 #34}, Moussa F. Optimization of ultra-high pressure liquid chromatography - tandem mass spectrometry determination in plasma and red blood cells of four sphingolipids and their evaluation as biomarker candidates of gaucher’s disease. J Chromatogr A. 2017;1525:116–125

39. Bastian M. HS, Jacomy M. Gephi: An open source software for exploring and manipulating networks. International AAAI Conference on Weblogs and Social Media. 2009

40. Blondel VD GG, Lambiotte R, Lefebvre E. Fast unfolding of communities in large networks. Journal of Statistical Mechanics: Theory and Experiment. 2008;10

41. GE vdMLaH. Visualizing high-dimensional data using t-sne. . Journal of Machine Learning Research 2008:2579–2605

42. Mukaka MM. Statistics corner: A guide to appropriate use of correlation coefficient in medical research. Malawi Med J. 2012;24:69–71

43. Schober P, Boer C, Schwarte LA. Correlation coefficients: Appropriate use and interpretation. Anesth Analg. 2018;126:1763–1768

44. Guo S, Yu Y, Zhang N, Cui Y, Zhai L, Li H, Zhang Y, Li F, Kan Y, Qin S. Higher level of plasma bioactive molecule sphingosine 1-phosphate in women is associated with estrogen. Biochim Biophys Acta. 2014;1841:836–846

45. Swendeman SL, Xiong Y, Cantalupo A, Yuan H, Burg N, Hisano Y, Cartier A, Liu CH, Engelbrecht E, Blaho V, Zhang Y, Yanagida K, Galvani S, Obinata H, Salmon JE, Sanchez T, Di Lorenzo A, Hla T. An engineered s1p chaperone attenuates hypertension and ischemic injury. Sci Signal. 2017;10

46. Cantalupo A, Zhang Y, Kothiya M, Galvani S, Obinata H, Bucci M, Giordano FJ, Jiang XC, Hla T, Di Lorenzo A. Nogo-b regulates endothelial sphingolipid homeostasis to control vascular function and blood pressure. Nat Med. 2015;21:1028–1037

47. Daum G, Winkler M, Moritz E, Muller T, Geffken M, von Lucadou M, Haddad M, Peine S, Boger RH, Larena-Avellaneda A, Debus ES, Graler M, Schwedhelm E. Determinants of serum- and plasma sphingosine-1-phosphate concentrations in a healthy study group. TH Open. 2020;4:e12–e19

48. Moritz E, Wegner D, Gross S, Bahls M, Dorr M, Felix SB, Ittermann T, Oswald S, Nauck M, Friedrich N, Boger RH, Daum G, Schwedhelm E, Rauch BH. Reference intervals for serum sphingosine-1-phosphate in the population-based study of health in pomerania. Clin Chim Acta. 2017;468:25–31

49. Deutschman DH, Carstens JS, Klepper RL, Smith WS, Page MT, Young TR, Gleason LA, Nakajima N, Sabbadini RA. Predicting obstructive coronary artery disease with serum sphingosine-1-phosphate. Am Heart J. 2003;146:62–68

50. Lertkiatmongkol P, Liao D, Mei H, Hu Y, Newman PJ. Endothelial functions of platelet/endothelial cell adhesion molecule-1 (cd31). Curr Opin Hematol. 2016;23:253–259

51. Privratsky JR, Newman DK, Newman PJ. Pecam-1: Conflicts of interest in inflammation. Life Sci. 2010;87:69–82

52. Huang YT, Chen SU, Chou CH, Lee H. Sphingosine 1-phosphate induces platelet/endothelial cell adhesion molecule-1 phosphorylation in human endothelial cells through csrc and fyn. Cell Signal. 2008;20:1521–1527

53. Robinson SD, Frenette PS, Rayburn H, Cummiskey M, Ullman-Culleré M, Wagner DD, Hynes RO. Multiple, targeted deficiencies in selectins reveal a predominant role for p-selectin in leukocyte recruitment. Proceedings of the National Academy of Sciences. 1999;96:11452–11457

54. Ridker PM, Buring JE, Rifai N. Soluble p-selectin and the risk of future cardiovascular events. Circulation. 2001;103:491–495

55. Yafasova A, Mandrup CM, Egelund J, Nyberg M, Stallknecht B, Hellsten Y, Nielsen LB, Christoffersen C. Effect of menopause and exercise training on plasma apolipoprotein m and sphingosine-1-phosphate. J Appl Physiol (1985). 2019;126:214–220

56. Chung MY, Park SY, Chung JO, Cho DH, Chung DJ. Plasma sphingosine 1-phosphate concentrations and cardiovascular autonomic neuropathy in individuals with type 2 diabetes. Scientific reports. 2020;10:12768

57. Winkler MS, Nierhaus A, Holzmann M, Mudersbach E, Bauer A, Robbe L, Zahrte C, Geffken M, Peine S, Schwedhelm E, Daum G, Kluge S, Zoellner C. Decreased serum concentrations of sphingosine-1-phosphate in sepsis. Crit Care. 2015;19:372

58. Ohkawa R, Nakamura K, Okubo S, Hosogaya S, Ozaki Y, Tozuka M, Osima N, Yokota H, Ikeda H, Yatomi Y. Plasma sphingosine-1-phosphate measurement in healthy subjects: Close correlation with red blood cell parameters. Ann Clin Biochem. 2008;45:356–363

59. Maranon R, Reckelhoff JF. Sex and gender differences in control of blood pressure. Clin Sci (Lond). 2013;125:311–318

60. Tipton AJ, Baban B, Sullivan JC. Female spontaneously hypertensive rats have a compensatory increase in renal regulatory t cells in response to elevations in blood pressure. Hypertension. 2014;64:557–564

61. Troseid M, Seljeflot I, Hjerkinn EM, Arnesen H. Interleukin-18 is a strong predictor of cardiovascular events in elderly men with the metabolic syndrome: Synergistic effect of inflammation and hyperglycemia. Diabetes Care. 2009;32:486–492

62. Formanowicz D, Wanic-Kossowska M, Pawliczak E, Radom M, Formanowicz P. Usefulness of serum interleukin-18 in predicting cardiovascular mortality in patients with chronic kidney disease--systems and clinical approach. Scientific reports. 2015;5:18332

63. Mallat Z, Corbaz A, Scoazec A, Besnard S, Leseche G, Chvatchko Y, Tedgui A. Expression of interleukin-18 in human atherosclerotic plaques and relation to plaque instability. Circulation. 2001;104:1598–1603

64. S OZ, Ulucam ZM. Association between interleukin-18 level and left ventricular mass index in hypertensive patients. Korean Circ J. 2017;47:238–244

65. Tipton AJ, Sullivan JC. Sex differences in t cells in hypertension. Clin Ther. 2014;36:1882–1900

66. Sullivan JC, Semprun-Prieto L, Boesen EI, Pollock DM, Pollock JS. Sex and sex hormones influence the development of albuminuria and renal macrophage infiltration in spontaneously hypertensive rats. Am J Physiol Regul Integr Comp Physiol. 2007;293:R1573–1579

67. Yeh CY, Adusumilli R, Kullolli M, Mallick P, John EM, Pitteri SJ. Assessing biological and technological variability in protein levels measured in pre-diagnostic plasma samples of women with breast cancer. Biomark Res. 2017;5:30

