## supplemental material for "Plasma S1P links to hypertension and biomarkers of inflammation and cardiovascular disease – findings from a translational investigation"

### Methods

***Ex vivo and in vitro testing:*** Mesenteric arteries isolated from male WT C57Bl/6N mice were cultured in DMEM (Gibco Life Technology, Sweden) containing 10% FBS (Gibco Life Technology, Sweden) and 1% Penicillin-Streptomycin (Sigma- Aldrich, Sweden) for 24 hrs in the presence of 1µM S1P (Cayman Chemicals, BioNordika, Sweden) or vehicle (4% fatty acid-free BSA). Endothelial cells of murine (bEND.3; ATCC® CRL-2299™) and human (HMEC-1; ATCC® CRL-3243™) origin were cultured in DMEM containing 10% FBS and 1% Penicillin-Streptomycin and MCDB131 supplemented with 10% FBS, 1% Penicillin-Streptomycin, 1% Glutamine (2mM), 1% NEAA, 1% sodium pyruvate, 0.1% Amphotericin B and 0.1% hEGF (10ng/ml), respectively. Human monocytic cells (THP-1; ATCC® TIB-202™) were cultured in DMEM containing 10% FBS and 1% Penicillin-Streptomycin. Cell cultures were maintained at 37°C with 5 % $CO_2$ and split 1:4 at a seeding density of $10^6$ cells. Cells were incubated with 1 µM S1P or vehicle (4% fatty acid-free BSA) for 24 hrs.

***Quantitative real-time PCR (qPCR):*** RNA was isolated from cultured cells or isolated mesenteric arteries (pooled from one mouse) using a Trizol® method. As per instructions using a “High Capacity Reverse Transcriptase” kit (AB Bioscience, Sweden), 1µg of total RNA was reverse transcribed with random hexamer primers. The resulting cDNA was diluted to a final volume of 200µl and subsequently used as a template for qPCR reactions. qPCR was performed in triplicates using *Power* SYBR® Green PCR Master Mix (Life Technology, Sweden) according to the manufacturer’s instructions. Each reaction comprised 2ng cDNA, 3ul master mix and 0.2uM final concentration of each primer. Cycling and detection were carried out using a CFX Connect™ Real-Time PCR Detection System and data quantified using Sequence CFX Manager™ Software (Biorad, Sweden). qPCR was performed for a total of 40 cycles (95°C 15 sec, 60°C 60 sec) followed by a dissociation stage. All data were normalized to species specific housekeeping genes L14 (mouse) and GPI (human) and quantification was carried out via the absolute method using standard curves generated from pooled cDNA representative of each sample to be analyzed.

| Target | Primer sequence (human) |
| --- | --- |
| **GPI** | F: 5'-AGG CTG CTG CCA CAT AAG GT-3’<br>R: 5'-CCA AGG CTC CAA GCA TGA AT-3' |
| **PAI** | F: 5'-CGCAAGGCACCTCTGAGAAC-3’<br>R: 5'-ACCTGCTGAAACACCCTCAC-3' |
| **IL18** | F: 5'-GATAGCCAGCCTAGAGGTATGG-3’<br>R: 5'-CCTTGATGTTATCAGGAGGATTCA-3' |

***ELISA:*** IL18 protein levels in plasma were determined using IL18 mouse ELISA kit (Nordic Biosite, Sweden) as per manufacturer’s instruction.

***Fluorescence activated cell sorting – in vitro experiments:*** Cells were collected and washed with PBS and incubated with Fc-block before samples were incubated with primary antibodies for 30 minutes at 4$^o$C. After centrifugation, the supernatant was decanted, washed, and re-suspended in FACS buffer (PBS + 2% FBS + 2mM EDTA; pH 7.4). Data acquisition was carried out in a BD ACCURI C6 Plus cytometer (BD Biosciences). Data analysis was performed with FlowJo software (version 10, TreeStar Inc., Ashland, OR, USA). Cells were plotted on forward versus side scatter and single cells were gated on FSC-A versus FSC-H linearity.

| Target | Primer sequence (mouse) |
| --- | --- |
| **L14** | F: 5'-GGCTTTAGTGGATGGACCCT-3’<br>R: 5'-ATTGATATCCGCCTTCTCCC-3' |
| **SELP** | F: 5’-AAGATGCCTGGCTACTGGACAC-3’<br>R: 5’-CAAGAGGCTGAACGCAGGTCAT-3’ |
| **PECAM-1** | F: 5'-GCAAATACCCACAGTTCCTCCC-3’<br>R: 5'-GGATGGTGAAGTTGGCTACAGG-3' |
| **Casp-3** | F: 5'-TGGTGATGAAGGGGTCATTTATG-3’<br>R: 5'-TTCGGCTTTCCAGTCAGACTC-3' |

**Supplemental Figure 1 – Overview human study cohort (MOS).**

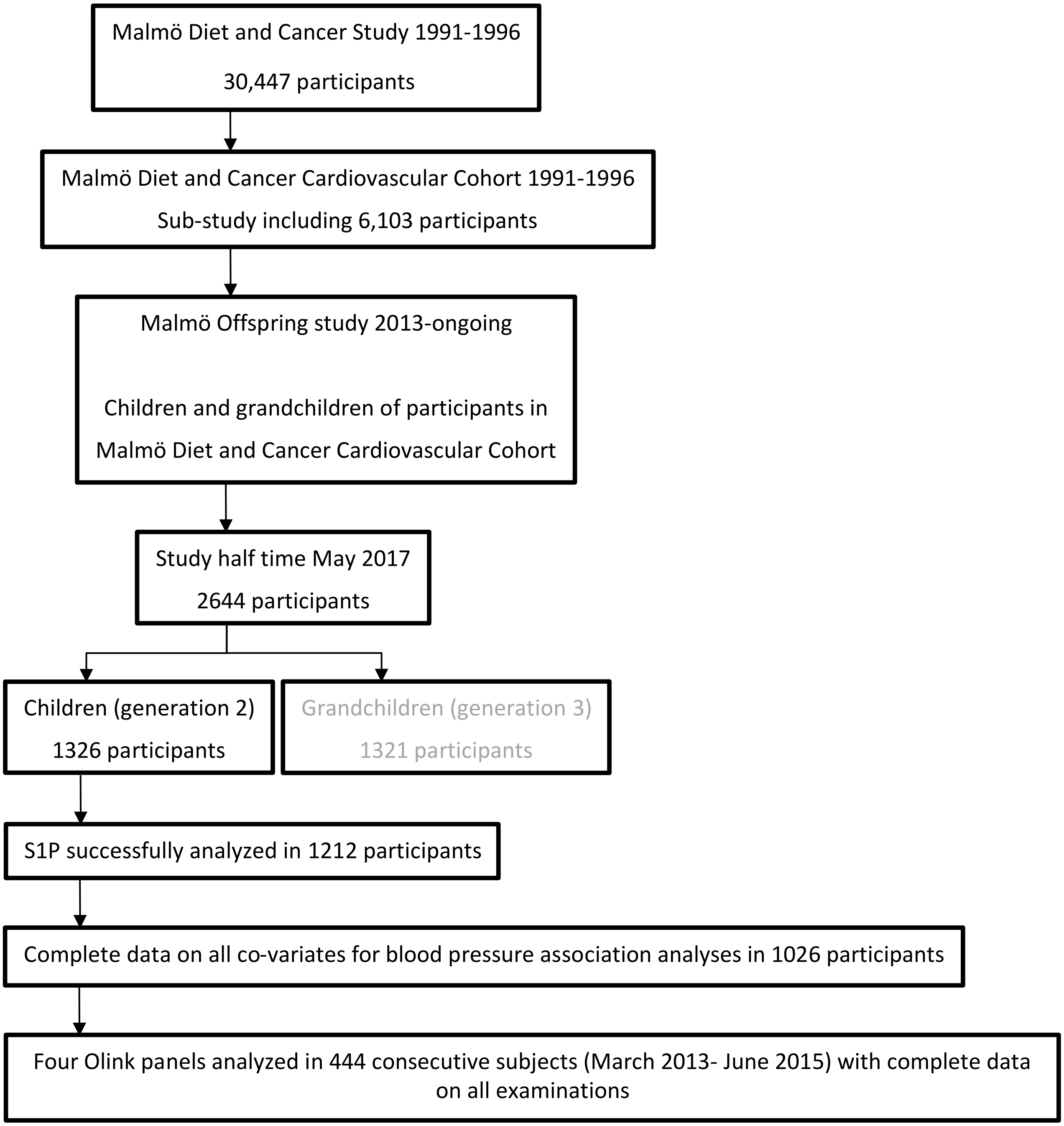

**Supplemental Figure 2 – S1P significantly associates with various inflammation, metabolism and CVD markers.** Overview of S1P correlations and associations with 299 proteins from |**A**| inflammation, |**B**| metabolism, |**C**| CDVII and |**D**| CDVIII OLINK panels. Pearson's correlations illustrate ranges from positive correlations in red to negative correlations in blue with reported correlation coefficients. Significance was tested in a two-tailed test where * denotes P≤0.05 and ** denotes P≤0.01. & denotes significance with panel-specific Bonferroni-correction of P-values (P≤7.81x10$^{-4}$ = 0.05/64 for the inflammation panel, P≤7.04x10$^{-4}$ = 0.05/71 for the metabolism panel, P≤5.95x10$^{-4}$ = 0.05/84 for the CVDII panel and P≤5.75x10$^{-4}$ = 0.05/87 for the CVDIII panel). β-values are given for all significant S1P associations with Bonferroni correction for all 299 markers. N=444.

**A**

| | inflammation panel (Pearson's r) | SIGNIF; 2-TAILED | SIGNIF; Bonferroni corrected | Linear Regression (β-value) | Linear Regression (P-value) |
|---|---|---|---|---|---|
| | S1P (nM) | S1P (nM) | S1P (nM) | S1P (nM) | S1P (nM) |
| S1P (nM) | 1,000 | | | | |
| IL8 | 0,137 | ** | | | |
| VEGFA | 0,259 | ** | & | 0.259 | 3,18x10$^{-8}$ |
| CDCP1 | 0,140 | ** | | | |
| CD244 | 0,196 | ** | & | 0,196 | 3,32x10$^{-5}$ |
| IL7 | 0,364 | ** | & | 0,364 | 2,37x10$^{-15}$ |
| OPG | 0,078 | | | | |
| LAP TGF-beta-1 | 0,269 | ** | & | 0,269 | 8,75x10$^{-9}$ |
| uPA | 0,094 | ** | | | |
| IL6 | 0,145 | ** | | | |
| MCP-1 | 0,073 | | | | |
| CXCL11 | 0,228 | ** | & | 0,228 | 1,18x10$^{-6}$ |
| AXIN1 | 0,331 | ** | & | 0,331 | 8,34x10$^{-13}$ |
| TRAIL | 0,135 | ** | | | |
| CXCL9 | -0,005 | | | | |
| CST5 | -0,040 | | | | |
| OSM | 0,202 | ** | & | 0,202 | 1,84x10$^{-5}$ |
| CXCL1 | 0,176 | ** | & | | |
| CCL4 | 0,122 | * | | | |
| CD6 | 0,046 | | | | |
| SCF | -0,047 | | | | |
| IL18 | 0,225 | ** | & | 0,231 | 1,36x10$^{-6}$ |
| TGF-alpha | 0,058 | | | | |
| MCP-4 | 0,228 | ** | & | 0,228 | 1,15x10$^{-6}$ |
| CCL11 | 0,122 | ** | | | |
| TNFSF14 | 0,279 | ** | & | 0,279 | 2,18x10$^{-9}$ |
| FGF-23 | 0,043 | | | | |
| MMP-1 | 0,223 | ** | & | 0,223 | 2,11x10$^{-6}$ |
| LIF-R | 0,086 | | | | |
| FGF-21 | 0,143 | ** | | | |
| CCL19 | 0,003 | | | | |
| IL-10RB | 0,110 | * | | | |
| IL-18R1 | 0,185 | ** | & | 0.185 | 8,59x10$^{-5}$ |
| PD-L1 | 0,076 | | | | |
| Beta-NGF | 0,063 | | | | |
| CXCL5 | 0,262 | ** | & | 0,262 | 2,12x10$^{-8}$ |
| TRANCE | 0,059 | | | | |
| HGF | 0,279 | ** | & | 0,279 | 2,31x10$^{-9}$ |
| IL-12B | -0,078 | | | | |
| MMP-10 | 0,055 | | | | |
| IL10 | 0,087 | | | | |
| CCL23 | 0,029 | | | | |
| CD5 | -0,015 | | | | |
| CCL3 | 0,079 | | | | |
| Flt3L | 0,047 | | | | |
| CXCL6 | 0,279 | ** | & | 0,279 | 2,33x10$^{-9}$ |
| CXCL10 | 0,033 | | | | |
| 4E-BP1 | 0,262 | ** | & | 0,262 | 2,23x10$^{-8}$ |
| SIRT2 | 0,370 | ** | & | 0,37 | 7,09x10$^{-16}$ |
| CCL28 | 0,103 | * | | | |
| DNER | 0,046 | | | | |
| EN-RAGE | 0,087 | | | | |
| CD40 | 0,224 | ** | & | 0,224 | 1,86x10$^{-6}$ |
| FGF-19 | 0,055 | | | | |
| MCP-2 | 0,178 | ** | & | | |
| CASP-8 | 0,210 | ** | & | 0,21 | 8,45x10$^{-6}$ |
| CCL25 | 0,097 | * | | | |
| CX3CL1 | -0,044 | | | | |
| TNFRSF9 | 0,037 | | | | |
| TWEAK | 0,125 | ** | | | |
| CCL20 | 0,053 | | | | |
| ST1A1 | 0,085 | | | | |
| STAMPB | 0,361 | ** | & | 0,361 | 3,95x10$^{-15}$ |
| ADA | 0,266 | ** | & | 0,266 | 1,30x10$^{-8}$ |
| TNFB | -0,041 | | | | |
| CSF-1 | 0,036 | | | | |

**. Correlation is significant at the 0.01 level (2-tailed).
*. Correlation is significant at the 0.05 level (2-tailed).
&. Correlation is significant after multiple comparisons.

**B**

| | metabolism panel (Pearson's r) | SIGNIF; 2-TAILED | SIGNIF; Bonferroni corrected | Linear Regression (β-value) | Linear Regression (P-value) |
|---|---|---|---|---|---|
| | S1P (nM) | S1P (nM) | S1P (nM) | S1P (nM) | S1P (nM) |
| S1P (nM) | 1,000 | | | | |
| CLMP | 0,000 | | | | |
| LRIG1 | 0,024 | | | | |
| NPTXR | 0,046 | | | | |
| THOP1 | 0,216 | ** | & | 0,216 | 3,75x10$^{-6}$ |
| CTSO | 0,134 | ** | | | |
| FCRL1 | -0,008 | | | | |
| CD164 | 0,192 | ** | & | 0,192 | 3,89x10$^{-5}$ |
| DDC | 0,072 | | | | |
| ACP6 | 0,163 | ** | | | |
| TFF2 | -0,045 | | | | |
| ANGPT2 | 0,037 | | | | |
| CD2AP | 0,259 | ** | & | 0,259 | 2,38x10$^{-8}$ |
| ANGPTL7 | 0,029 | | | | |
| CLEC5A | 0,019 | | | | |
| TINAGL1 | -0,005 | | | | |
| ENO2 | 0,289 | ** | & | 0,289 | 4,07x10$^{-10}$ |
| NADK | 0,129 | ** | | | |
| GHRL | -0,029 | | | | |
| SERPINB6 | 0,316 | ** | & | 0,316 | 6,41x10$^{-12}$ |
| CDHR5 | 0,038 | | | | |
| CCDC80 | 0,088 | | | | |
| CA13 | 0,325 | ** | & | 0,325 | 1,59x10$^{-12}$ |
| SEMA3F | 0,004 | | | | |
| KLK10 | -0,067 | | | | |
| PILRB | 0,014 | | | | |
| ANGPTL1 | 0,018 | | | | |
| APLP1 | -0,121 | ** | | | |
| ADGRG2 | -0,006 | | | | |
| TYMP | 0,077 | | | | |
| GRAP2 | 0,296 | ** | & | 0,296 | 1,33x10$^{-10}$ |
| LILRA5 | 0,144 | ** | | | |
| ALDH1A1 | -0,004 | | | | |
| CD79B | 0,024 | | | | |
| ANXA11 | 0,313 | ** | & | 0,313 | 1,48x10$^{-11}$ |
| SIGLEC7 | 0,083 | | | | |
| QDPR | 0,324 | ** | & | 0,324 | 1,78x10$^{-12}$ |
| SNAP23 | 0,299 | ** | & | 0,299 | 9,46x10$^{-11}$ |
| CLSTN2 | 0,096 | * | | | |
| COMT | 0,325 | ** | & | 0,325 | 1,56x10$^{-12}$ |
| CLUL1 | -0,019 | | | | |
| CHRDL2 | 0,085 | | | | |
| NOMO1 | 0,153 | ** | | | |
| SOST | 0,035 | | | | |
| FAM3C | 0,025 | | | | |
| TXNDC5 | 0,279 | ** | & | 0,79 | 1,97x10$^{-9}$ |
| PPP1R2 | 0,324 | ** | & | 0,324 | 1,70x10$^{-12}$ |
| LRP11 | 0,015 | | | | |
| ADGRE2 | 0,048 | | | | |
| ENPP7 | 0,175 | ** | | | |
| MCFD2 | 0,272 | ** | & | 0,272 | 4,10x10$^{-9}$ |
| REG4 | 0,001 | | | | |
| SUMF2 | 0,254 | ** | & | 0,254 | 4,64x10$^{-8}$ |
| CANT1 | 0,082 | | | | |
| CD1C | -0,009 | | | | |
| GAL | 0,022 | | | | |
| CDH2 | 0,134 | ** | | | |
| TYRO3 | 0,072 | | | | |
| CRKL | 0,336 | ** | & | 0,336 | 2,49x10$^{-13}$ |
| IGFBPL1 | 0,101 | * | | | |
| VCAN | 0,003 | | | | |
| TSHB | -0,082 | | | | |
| BAG6 | 0,278 | ** | & | 0,278 | 2,03x10$^{-9}$ |
| NECTIN2 | 0,124 | ** | | | |
| USP8 | 0,232 | ** | & | 0,232 | 2,22x10$^{-6}$ |
| FKBP4 | 0,285 | ** | & | 0,285 | 5,87x10$^{-9}$ |
| SDC4 | 0,364 | ** | & | 0,364 | 1,44x10$^{-15}$ |
| PAG1 | 0,215 | ** | & | 0,15 | 4,10x10$^{-6}$ |
| KYAT1 | 0,312 | ** | & | 0,312 | 1,26x10$^{-11}$ |
| DAB2 | 0,296 | ** | & | 0,296 | 1,54x10$^{-10}$ |
| NPDC1 | 0,031 | | | | |
| METRNL | 0,009 | | | | |
| ROR1 | -0,040 | | | | |

**. Correlation is significant at the 0.01 level (2-tailed).
*. Correlation is significant at the 0.05 level (2-tailed).
&. Correlation is significant after multiple comparisons.

**C**

| | CVDII panel (Pearson's r) | SIGNIF; 2-TAILED | SIGNIF; Bonferroni corrected | Linear Regression (β-value) | Linear Regression (P-value) |
|---|---|---|---|---|---|
| | S1P (nM) | S1P (nM) | S1P (nM) | S1P (nM) | S1P (nM) |
| S1P (nM) | 1,000 | | | | |
| BMP-6 | 0,200 | ** | & | 0,2 | 1,99x10$^{-5}$ |
| ANG-1 | 0,282 | ** | & | 0,282 | 6,45x10$^{-10}$ |
| ADM | 0,123 | ** | | | |
| CD40-L | 0,340 | ** | & | 0,34 | 1,45x10$^{-13}$ |
| PIGF | 0,107 | * | | | |
| ADAM-TS13 | 0,036 | | | | |
| Protein BOC | 0,053 | | | | |
| SRC | -0,267 | ** | & | -0,267 | 8,99x10$^{-9}$ |
| IL-1ra | 0,087 | | | | |
| IL-6 | 0,163 | ** | | | |
| TNFRSF10A | 0,029 | | | | |
| STK4 | 0,170 | ** | | | |
| IDUA | 0,275 | ** | & | 0,275 | 3,44x10$^{-9}$ |
| TNFRSF11A | 0,093 | * | | | |
| PAR-1 | 0,295 | ** | & | 0,295 | 1,93x10$^{-10}$ |
| TRAIL-R2 | 0,061 | | | | |
| PRSS27 | 0,103 | * | | | |
| TIE2 | 0,120 | ** | | | |
| TF | 0,030 | ** | | | |
| IL1RL2 | 0,081 | | | | |
| PDGF subunit B | 0,345 | ** | & | 0,345 | 6,02x10$^{-14}$ |
| IL27 | 0,012 | | | | |
| IL-17D | 0,020 | | | | |
| CXCL1 | 0,206 | ** | & | 0,206 | 1,09x10$^{-5}$ |
| LOX-1 | 0,238 | ** | & | 0,238 | 3,42x10$^{-7}$ |
| Gal-9 | 0,086 | * | | | |
| GIF | 0,040 | | | | |
| SCF | -0,028 | | | | |
| IL-18 | 0,246 | ** | & | 0,246 | 1,38x10$^{-7}$ |
| FGF-21 | 0,148 | ** | | | |
| PIgR | 0,086 | * | | | |
| RAGE | -0,022 | | | | |
| SOD2 | 0,275 | ** | & | 0,275 | 3,38x10$^{-9}$ |
| CTRC | -0,014 | | | | |
| FGF-23 | 0,076 | | | | |
| SPON2 | 0,081 | | | | |
| GH | -0,039 | | | | |
| FS | 0,152 | ** | | | |
| GLO1 | 0,318 | ** | & | 0,318 | 5,50x10$^{-12}$ |
| CD84 | 0,345 | ** | & | 0,345 | 6,29x10$^{-13}$ |
| SERPINA12 | -0,054 | | | | |
| REN | 0,107 | * | | | |
| DECR1 | 0,255 | ** | & | 0,255 | 4,27x10$^{-8}$ |
| MERTK | 0,098 | * | | | |
| KIM-1 | 0,176 | ** | & | | |
| THBS2 | 0,086 | * | | | |
| TM | 0,145 | ** | | | |
| VSIG2 | 0,052 | | | | |
| AMBP | 0,049 | | | | |
| PRELP | 0,089 | * | | | |
| HO-1 | 0,101 | * | | | |
| XCL1 | 0,058 | | | | |
| IL16 | 0,148 | ** | | | |
| SORT1 | 0,316 | ** | & | 0,316 | 7,37x10$^{-12}$ |
| CEACAM8 | 0,165 | ** | | | |
| PTX3 | -0,054 | | | | |
| PSGL-1 | 0,091 | * | | | |
| CCL17 | 0,245 | ** | & | 0,245 | 1,54x10$^{-7}$ |
| CCL3 | 0,080 | | | | |
| MMP-7 | 0,107 | * | | | |
| IgG Fc receptor II-b | 0,088 | | | | |
| ITGB1BP2 | 0,422 | ** | & | 0,422 | 1,14x10$^{-20}$ |
| DCN | -0,003 | | | | |
| Dkk-1 | 0,354 | ** | & | 0,354 | 1,20x10$^{-14}$ |
| LPL | 0,000 | | | | |
| PRSS8 | 0,114 | ** | | | |
| AGRP | 0,033 | | | | |
| HB-EGF | 0,366 | ** | & | 0,366 | 1,25x10$^{-15}$ |
| GDF-2 | 0,001 | | | | |
| FABP2 | 0,062 | | | | |
| THPO | 0,273 | ** | & | 0,273 | 4,75x10$^{-9}$ |
| MARCO | 0,152 | ** | | | |
| MMP-12 | -0,025 | | | | |
| ACE2 | 0,147 | ** | | | |
| PD-L2 | -0,040 | | | | |
| CTSL1 | 0,097 | | | | |
| hOSCAR | 0,087 | * | | | |
| TNFRSF13B | 0,036 | | | | |
| TGM2 | 0,080 | | | | |
| LEP | 0,040 | | | | |
| HSP 27 | -0,136 | ** | | | |
| CD4 | 0,018 | | | | |
| NEMO | 0,352 | ** | & | 0,352 | 1,57x10$^{-14}$ |
| VEGF-D | 0,038 | | | | |
| HAOX1 | 0,101 | * | | | |

**. Correlation is significant at the 0.01 level (2-tailed).
*. Correlation is significant at the 0.05 level (2-tailed).
&. Correlation is significant after multiple comparisons.

**D**

| | CVDIII panel (Pearson's r) | SIGNIF; 2-TAILED | SIGNIF; Bonferroni corrected | Linear Regression (β-value) | Linear Regression (P-value) |
|---|---|---|---|---|---|
| | S1P (nM) | S1P (nM) | S1P (nM) | S1P (nM) | S1P (nM) |
| S1P (nM) | 1,000 | | | | |
| TNFRSF14 | 0,264 | ** | & | 0,264 | 1,26x10$^{-8}$ |
| LDL receptor | 0,132 | ** | | | |
| ITGB2 | 0,025 | | | | |
| IL-17RA | 0,166 | ** | & | | |
| TNF-R2 | 0,086 | | | | |
| MMP-9 | 0,231 | ** | & | 0,231 | 6,72x10$^{-7}$ |
| IL2-RA | 0,086 | * | | | |
| OPG | 0,110 | * | | | |
| ALCAM | 0,097 | * | | | |
| TFF3 | -0,029 | | | | |
| SELP | 0,384 | ** | & | 0,384 | 2,32x10$^{-17}$ |
| CSTB | 0,247 | ** | & | 0,247 | 1,01x10$^{-7}$ |
| MCP-1 | 0,112 | * | | | |
| CD163 | 0,155 | ** | | | |
| Gal-3 | 0,114 | * | | | |
| GRN | 0,141 | ** | | | |
| MEPE | 0,114 | * | | | |
| BLM hydrolase | 0,313 | ** | & | 0,313 | 9,90x10$^{-12}$ |
| PLC | 0,114 | * | | | |
| LTBR | 0,124 | ** | | | |
| Notch 3 | 0,057 | | | | |
| TIMP4 | 0,122 | * | | | |
| CNTN1 | 0,052 | | | | |
| CDH5 | 0,060 | | | | |
| TLT-2 | 0,181 | ** | & | 0,181 | 1,10x10$^{-4}$ |
| FABP4 | 0,095 | * | | | |
| TFPI | 0,168 | ** | & | | |
| PAI | 0,409 | ** | & | 0,409 | 1,15x10$^{-19}$ |
| CCL24 | 0,095 | * | | | |
| TR | 0,015 | | | | |
| TNFRSF10C | 0,136 | ** | | | |
| GDF-15 | 0,149 | ** | | | |
| SELE | 0,234 | ** | & | 0,234 | 5,02x10$^{-7}$ |
| AZU1 | 0,217 | ** | & | 0,217 | 3,13x10$^{-6}$ |
| DLK-1 | -0,005 | | | | |
| MPO | 0,174 | ** | & | | |
| CXCL16 | 0,094 | * | | | |
| IL-6RA | 0,147 | ** | | | |
| RETN | 0,116 | * | | | |
| IGFBP-1 | -0,024 | | | | |
| CHIT1 | 0,048 | | | | |
| TR-AP | 0,211 | ** | & | 0,211 | 5,94x10$^{-6}$ |
| CCL22 | 0,119 | ** | | | |
| PI3 | 0,126 | ** | | | |
| Ep-CAM | 0,002 | | | | |
| AP-N | 0,125 | ** | | | |
| AXL | 0,089 | * | | | |
| IL-1RT1 | 0,104 | * | | | |
| MMP-2 | 0,101 | * | | | |
| FAS | 0,172 | ** | & | | |
| MB | 0,038 | | | | |
| TNFSF13B | 0,142 | ** | | | |
| PRTN3 | 0,138 | ** | | | |
| PCSK9 | -0,010 | | | | |
| U-PAR | 0,170 | ** | & | | |
| OPN | 0,165 | ** | | | |
| CTSD | 0,248 | ** | & | 0,248 | 1,03x10$^{-7}$ |
| PGLYRP1 | 0,156 | ** | | | |
| CPA1 | 0,039 | | | | |
| JAM-A | 0,380 | ** | & | 0,38 | 5,64x10$^{-17}$ |
| Gal-4 | 0,112 | * | | | |
| IL-1RT2 | 0,172 | ** | | | |
| SHPS-1 | 0,045 | | | | |
| CCL15 | 0,123 | ** | | | |
| CASP-3 | 0,403 | ** | & | 0,403 | 4,54x10$^{-19}$ |
| uPA | 0,143 | ** | | | |
| CPB1 | 0,051 | | | | |
| CHI3L1 | 0,173 | ** | & | | |
| ST2 | 0,157 | * | | | |
| t-PA | 0,162 | | | | |
| SCGB3A2 | 0,061 | | | | |
| EGFR | 0,104 | * | | | |
| IGFBP-7 | 0,130 | ** | | | |
| CD93 | 0,073 | | | | |
| IL-18BP | 0,096 | * | | | |
| COL1A1 | 0,123 | ** | | | |
| PON3 | 0,027 | | | | |
| CTSZ | 0,181 | ** | & | | |
| MMP-3 | 0,139 | ** | | | |
| RARRES2 | 0,188 | ** | & | 0,188 | 5,69x10$^{-5}$ |
| ICAM-2 | 0,103 | * | | | |
| KLK6 | 0,097 | * | | | |
| PDGF subunit A | 0,392 | ** | & | 0,392 | 4,94x10$^{-18}$ |
| TNF-R1 | 0,105 | * | | | |
| IGFBP-2 | 0,013 | | | | |
| vWF | 0,024 | | | | |
| PECAM-1 | 0,389 | ** | & | 0,378 | 8,57x10$^{-18}$ |
| CCL16 | 0,169 | ** | & | | |

**. Correlation is significant at the 0.01 level (2-tailed).
*. Correlation is significant at the 0.05 level (2-tailed).
&. Correlation is significant after multiple comparisons.

**Supplemental Figure 3 – t-distributed stochastic neighbor embedding (t-SNE) of correlation data of all OLINK panels combined.** t-SNE locations of markers are meaningful where proximity is explained by similarity in correlations with other markers. The t-SNE arranges the markers into 9 visual groups that were associated with processes related to inflammation and CVD. N=444

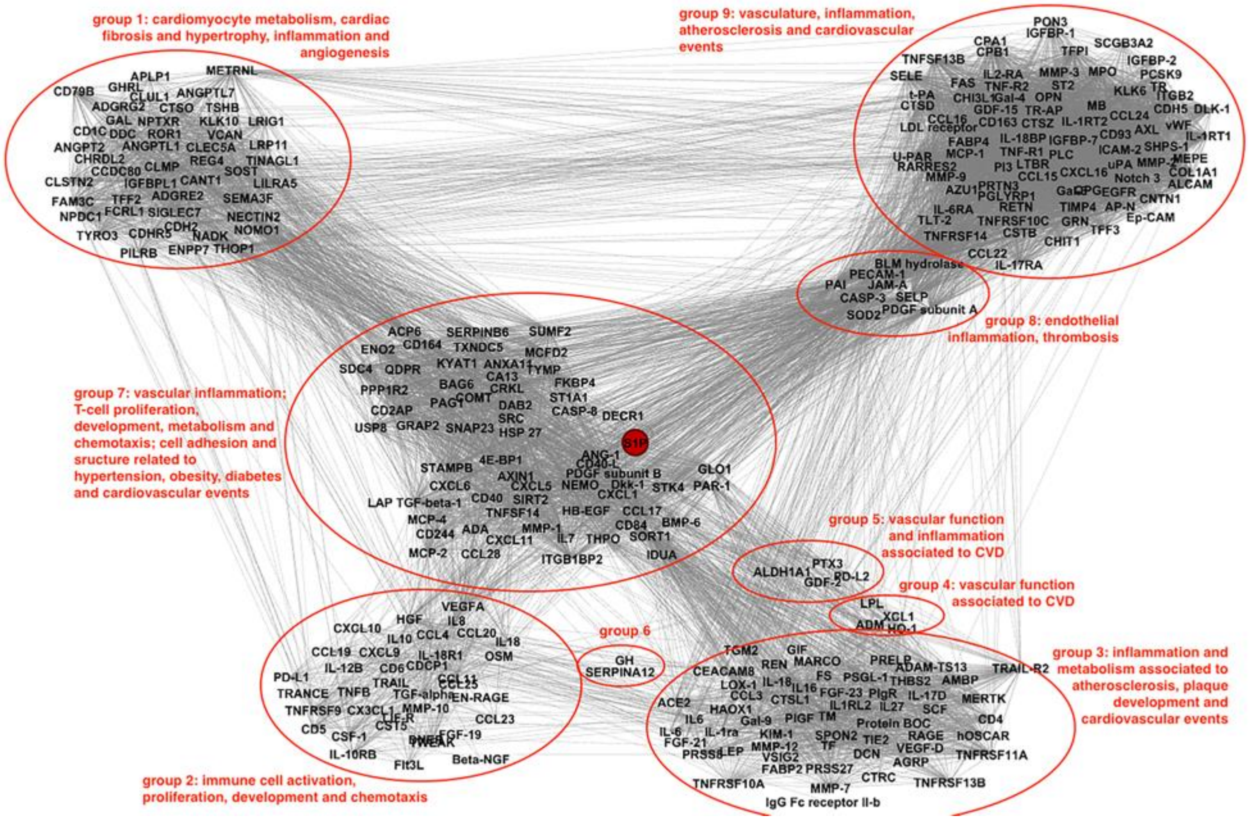

**Supplemental Figure 4 – Correlation matrices** for |**A**| inflammation OLINK panel (64 markers above detection limit), |**B**| metabolism OLINK panel (71 markers above detection limit), |**C**| CVDII OLINK panel (84 markers above detection limit) and |**D**| CVDIII OLINK panel (87 markers above detection limit). N=444

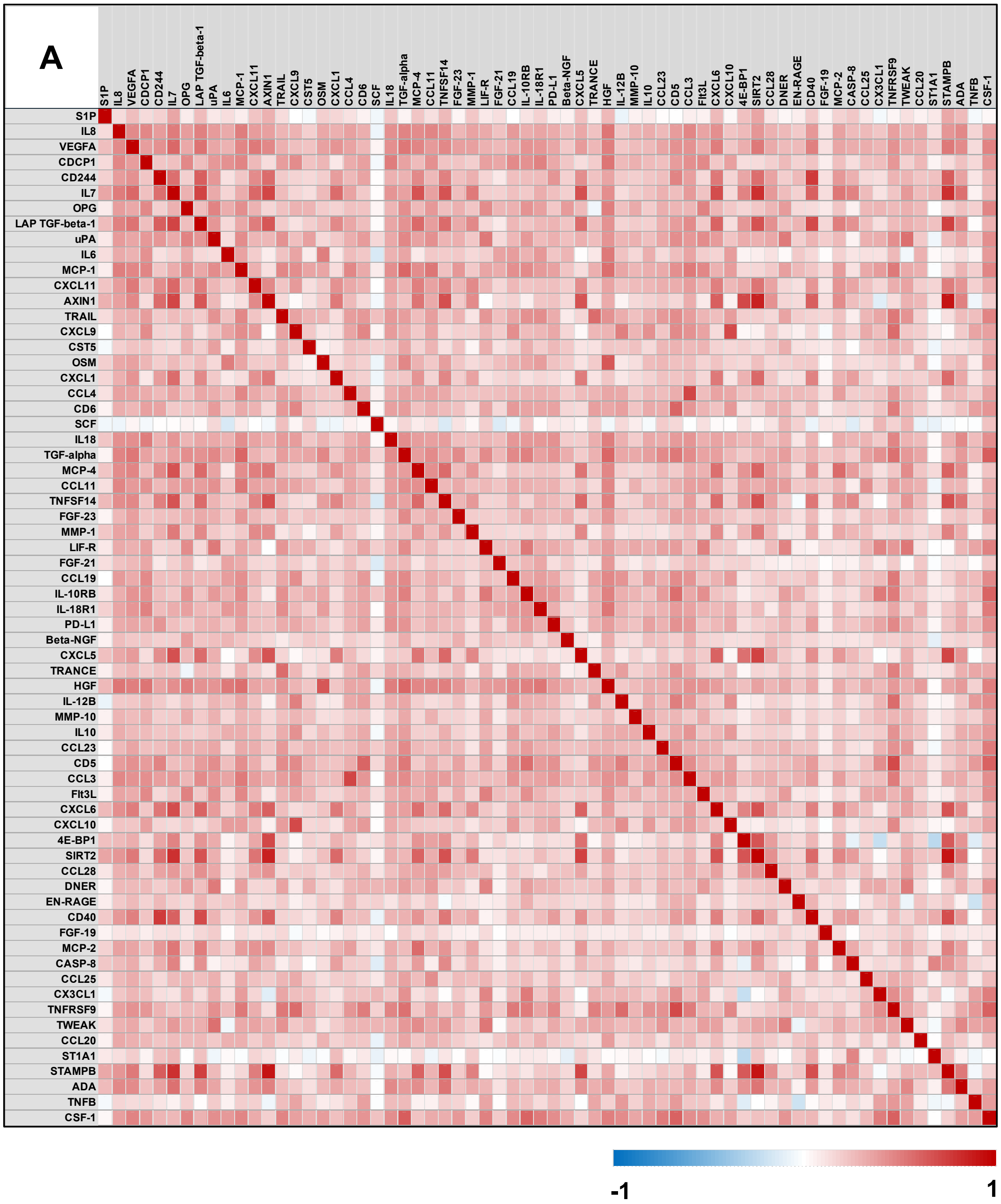

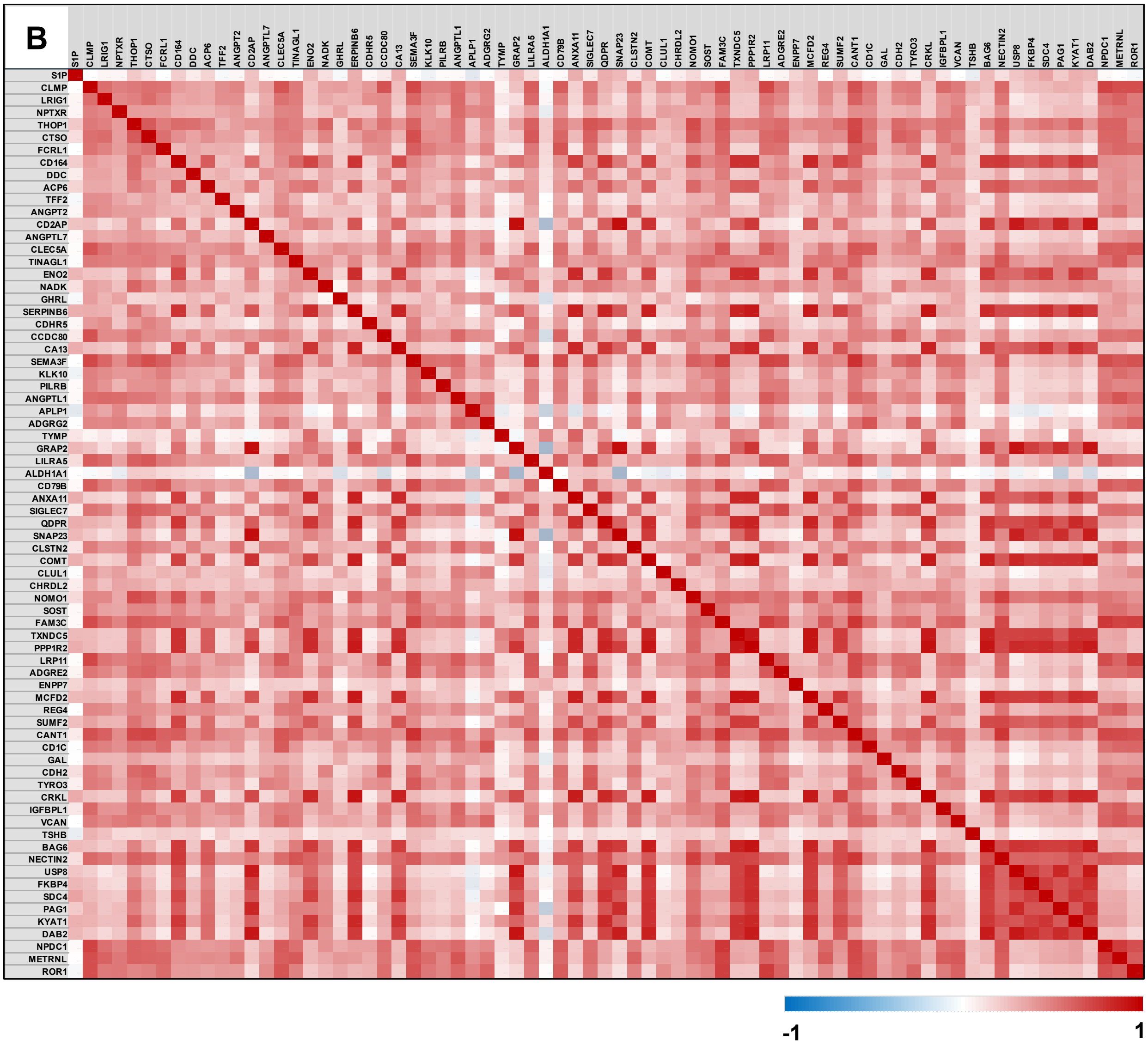

B
S1P
CLMP
LRIG1
NPTXR
THOP1
CTSO
FCRL1
CD164
DDC
ACP6
TFF2
ANGPT2
CD2AP
ANGPTL7
CLEC5A
TINAGL1
ENO2
NADK
GHRL
SERPINB6
CDHR5
CCDC80
CA13
SEMA3F
KLK10
PILRB
ANGPTL1
APLP1
ADGRG2
TYMP
GRAP2
LILRA5
ALDH1A1
CD79B
ANXA11
SIGLEC7
QDPR
SNAP23
CLSTN2
COMT
CLUL1
CHRDL2
NOMO1
SOST
FAM3C
TXNDC5
PPP1R2
LRP11
ADGRE2
ENPP7
MCFD2
REG4
SUMF2
CANT1
CD1C
GAL
CDH2
TYRO3
CRKL
IGFBPL1
VCAN
TSHB
BAG6
NECTIN2
USP8
FKBP4
SDC4
PAG1
KYAT1
DAB2
NPDC1
METRNL
ROR1
-1
1

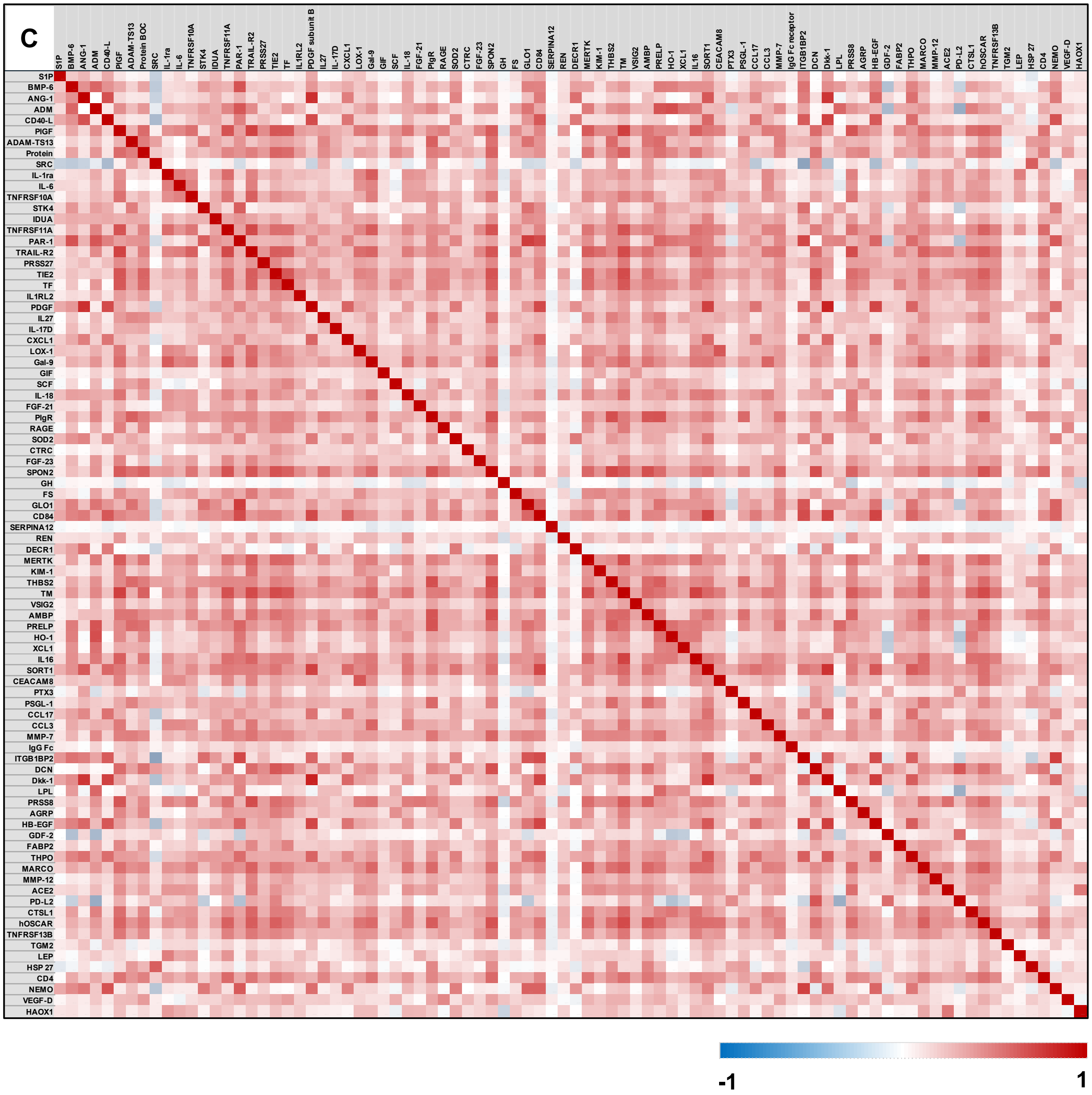
C
S1P
BMP-6
ANG-1
ADM
CD40-L
PIGF
ADAM-TS13
Protein BOC
SRC
IL-1ra
IL-6
TNFRSF10A
STK4
IDUA
TNFRSF11A
PAR-1
TRAIL-R2
PRSS27
TIE2
TF
IL1RL2
PDGF subunit B
IL27
IL-17D
CXCL1
LOX-1
Gal-9
GIF
SCF
IL-18
FGF-21
PIgR
RAGE
SOD2
CTRC
FGF-23
SPON2
GH
FS
GLO1
CD84
SERPINA12
REN
DECR1
MERTK
KIM-1
THBS2
TM
VSIG2
AMBP
PRELP
HO-1
XCL1
IL16
SORT1
CEACAM8
PTX3
PSGL-1
CCL17
CCL3
MMP-7
IgG Fc receptor
ITGB1BP2
DCN
Dkk-1
LPL
PRSS8
AGRP
HB-EGF
GDF-2
FABP2
THPO
MARCO
MMP-12
ACE2
PD-L2
CTSL1
hOSCAR
TNFRSF13B
TGM2
LEP
HSP 27
CD4
NEMO
VEGF-D
HAOX1
-1
1

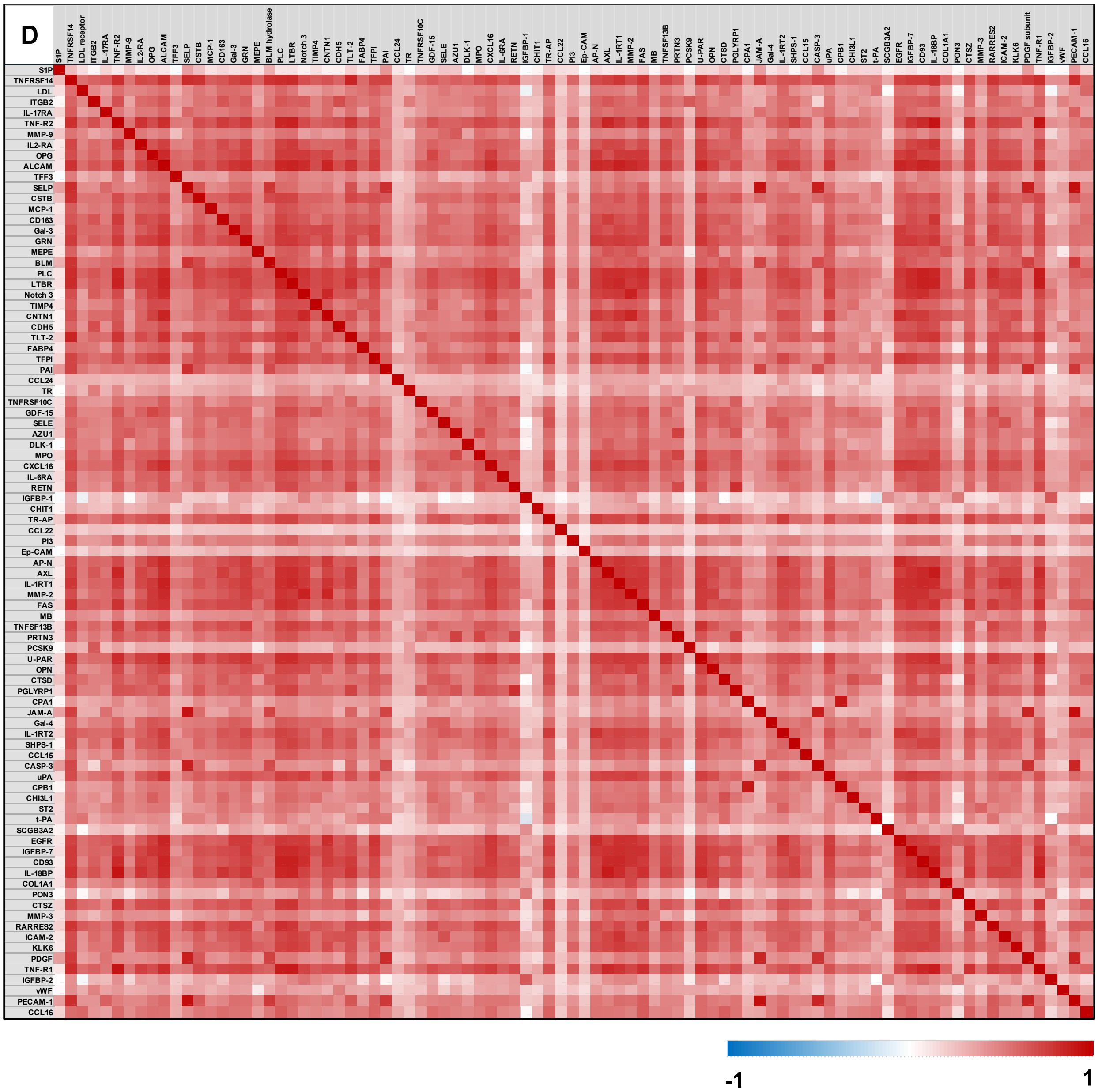
D
S1P
TNFRSF14
LDL receptor
ITGB2
IL-17RA
TNF-R2
MMP-9
IL2-RA
OPG
ALCAM
TFF3
SELP
CSTB
MCP-1
CD163
Gal-3
GRN
MEPE
BLM hydrolase
PLC
LTBR
Notch 3
TIMP4
CNTN1
CDH5
TLT-2
FABP4
TFPI
PAI
CCL24
TR
TNFRSF10C
GDF-15
SELE
AZU1
DLK-1
MPO
CXCL16
IL-6RA
RETN
IGFBP-1
CHIT1
TR-AP
CCL22
PI3
Ep-CAM
AP-N
AXL
IL-1RT1
MMP-2
FAS
MB
TNFSF13B
PRTN3
PCSK9
U-PAR
OPN
CTSD
PGLYRP1
CPA1
JAM-A
Gal-4
IL-1RT2
SHPS-1
CCL15
CASP-3
uPA
CPB1
CHI3L1
ST2
t-PA
SCGB3A2
EGFR
IGFBP-7
CD93
IL-18BP
COL1A1
PON3
CTSZ
MMP-3
RARRES2
ICAM-2
KLK6
PDGF subunit
TNF-R1
IGFBP-2
vWF
PECAM-1
CCL16
S1P
TNFRSF14
LDL
ITGB2
IL-17RA
TNF-R2
MMP-9
IL2-RA
OPG
ALCAM
TFF3
SELP
CSTB
MCP-1
CD163
Gal-3
GRN
MEPE
BLM
PLC
LTBR
Notch 3
TIMP4
CNTN1
CDH5
TLT-2
FABP4
TFPI
PAI
CCL24
TR
TNFRSF10C
GDF-15
SELE
AZU1
DLK-1
MPO
CXCL16
IL-6RA
RETN
IGFBP-1
CHIT1
TR-AP
CCL22
PI3
Ep-CAM
AP-N
AXL
IL-1RT1
MMP-2
FAS
MB
TNFSF13B
PRTN3
PCSK9
U-PAR
OPN
CTSD
PGLYRP1
CPA1
JAM-A
Gal-4
IL-1RT2
SHPS-1
CCL15
CASP-3
uPA
CPB1
CHI3L1
ST2
t-PA
SCGB3A2
EGFR
IGFBP-7
CD93
IL-18BP
COL1A1
PON3
CTSZ
MMP-3
RARRES2
ICAM-2
KLK6
PDGF
TNF-R1
IGFBP-2
vWF
PECAM-1
CCL16
-1
1

**Supplemental Figure 5 – S1P induces the expression of different markers of inflammation and vascular dysfunction.** **|A|** Augmentation of the pro-angiogenic marker angiopoietin-1 (Ang-1) mRNA expression in response to 1 µM S1P (24hrs) in small resistance arteries isolated from WT mice (N=5 per group). **|B|** Augmentation of caspase 3 (CASP3) mRNA expression in murine endothelial cells in response to 1 µM S1P (6hrs; N=3 per group in triplicates). **|C|** Augmentation of interleukin 18 (IL18) mRNA expression in human endothelial cells in response to 1 µM S1P (12hrs; N=3 per group in triplicates). **|D|** Increase of CD40 protein surface expression in response to 1 µM S1P (24hrs) on human monocytic cells (THP-1; N=3 per group). Representative dot plots illustrating shift in fluorescent signal for CD40 (x-axis). * *denotes* $P \leq 0.05$ *compared to respective control after unpaired t-test.*

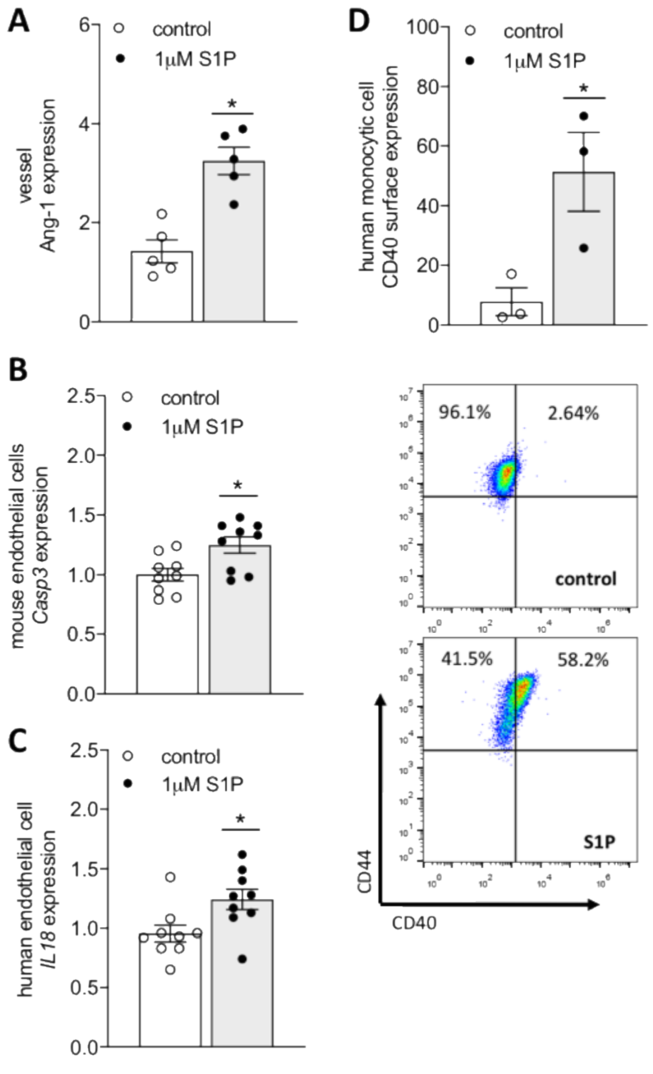

**Supplemental Table S1 - Proteins with ≥15% samples below limit of detection in OLINK panels that were excluded from the analyses.**

| # | Inflammation panel | Metabolism panel | CVDII panel | CVDIII panel |
|---|---|---|---|---|
| 1 | MCP-3 | AHCY | SLAMF7 | EPHB4 |
| 2 | GDNF | S100P | IL4RA | SPON 1 |
| 3 | IL-17C | GLRX | PAPPA | PSPD |
| 4 | IL17A | SERPINB8 | GT | Nt-proBNP |
| 5 | IL20RA | DIABLO | BNP | |
| 6 | IL2RB | ANXA4 | CA5A | |
| 7 | IL1alpha | ITGB7 | PARP1 | |
| 8 | TGFalpha | APEX1 | | |
| 9 | TSLP | ENTPD5 | | |
| 10 | IL2 | HDGF | | |
| 11 | IL10RA | CTSH | | |
| 12 | FGF5 | NQO2 | | |
| 13 | IL15RA | DPP7 | | |
| 14 | IL22RA1 | SSC4D | | |
| 15 | IL24 | RTN4R | | |
| 14 | IL13 | FBP1 | | |
| 17 | ARTN | ARG1 | | |
| 18 | TNF | MEP1B | | |
| 19 | IL20 | NTproBNP | | |
| 20 | IL33 | RNASE3 | | |
| 21 | IFNgamma | | | |

| | |
|---|---|
| 22 | IL4 |
| 23 | LIF |
| 24 | NRTN |
| 25 | NT3 |
| 26 | IL5 |

**Supplemental Table S2 - List of overlapping proteins between the different OLINK panels.**

| Target Protein | CVDII | CVDIII | Inflammation | Metabolism |
|---|---|---|---|---|
| Interleukin-6 (IL6) | x | | x | |
| Stem cell factor (SCF) | x | | x | |
| C-C motif chemokine 3 (CCL3) | x | | x | |
| C-X-C motif chemokine 1 (CXCL1) | x | | x | |
| Interleukin-18 (IL-18) | x | | x | |
| Fibroblast growth factor 21 (FGF-21) | x | | x | |
| Fibroblast growth factor 23 (FGF-23) | x | | x | |
| N-terminal prohormone brain natriuretic peptide (NT-proBNP) | | x | | x |
| Monocyte chemotactic protein 1 (MCP-1) | | x | x | |
| Osteoprotegerin (OPG) | | x | x | |
| Urokinase-type plasminogen activator (uPA) | | x | x | |

**Supplemental Table S3 - Associations between S1P and systolic blood pressure and diastolic blood pressure.** Values are β-coefficients and significance values (P) for linear regression analyses. AHT=anti-hypertensive treatment. BMI=body mass index; DBP=diastolic blood pressure; eGFR=estimated glomerular filtration rate; SBP=systolic blood pressure.

| **Linear regression analyses** | | | | |
|---|---|---|---|---|
| | **SBP** | | **DBP** | |
| | **β** | **P** | **β** | **P** |
| **Unadjusted** | | | | |
| S1P | 1.467 | 0.003 | 0.483 | 0.098 |
| **Model 1** | | | | |
| S1P | 1.103 | 0.013 | 0.346 | 0.217 |
| Age | 0.714 | $4.38x10^{-34}$ | 0.327 | $2.33x10^{-19}$ |
| Sex | -5.772 | $1.91x10^{-10}$ | -1.258 | 0.026 |
| **Model 2** | | | | |
| S1P | 1.057 | 0.015 | 0.263 | 0.334 |
| Age | 0.604 | $7.43x10^{-21}$ | 0.264 | $3.73x10^{-11}$ |
| Sex | -4.387 | $1.55x10^{-6}$ | -0.323 | 0.569 |
| BMI | 0.662 | $1.77x10^{-10}$ | 0.531 | $3.74x10^{-16}$ |
| eGFR | -0.045 | 0.373 | -0.043 | 0.170 |
| Diabetes status | 1.595 | 0.484 | 3.668 | 0.010 |
| AHT | 5.955 | $1.52x10^{-5}$ | 2.770 | 0.001 |
| Smoking | 1.826 | 0.234 | 2.038 | 0.034 |
| Alcohol | | | | |
| Never | 0.866 | 0.641 | -0.696 | 0.548 |

| | | | | |
|---|---|---|---|---|
| Monthly or less | -2.540 | 0.061 | -0.974 | 0.249 |
| 2-4 times a month | -0.529 | 0.609 | 0.483 | 0.455 |
| 2-3 times a week | -0.866 | 0.641 | 0.698 | 0.548 |
| ≥ 4 times a week | -3.400 | 0.076 | -1.349 | 0.260 |

**Supplemental Table S4 – Associations of all proteins from four proteomic panels (inflammation, metabolism, CVDII and CVDIII) and S1P** (proteins that meet age and sex adjusted Bonferroni-corrected significant associations with S1P (0.05/299; P≤1.67x10$^{-4}$) in bold type).

INFLAMMATION PANEL

| Model | | Unstandardized Coefficients | | Standardized Coefficients | p |
|---|---|---|---|---|---|
| | | β | Std. Error | Beta | |
| **IL8** | | | | | |
| Unadjusted | S1P | 0.269 | 0.092 | 0.137 | 0.004 |
| Age and sex adjusted | S1P | 0.247 | 0.093 | 0.126 | 0.008 |
| | Age | 0.007 | 0.004 | 0.099 | 0.036 |
| | Sex | -0.060 | 0.056 | -0.051 | 0.280 |
| **VEGFA** | | | | | |
| **Unadjusted** | **S1P** | **0.377** | **0.067** | **0.259** | **3.18x10-8** |
| **Age and sex adjusted** | **S1P** | **0.380** | **0.067** | **0.261** | **2.89x10-8** |
| | **Age** | **0.004** | **0.003** | **0.069** | **0.131** |
| | **Sex** | **0.044** | **0.040** | **0.051** | **0.272** |
| **CDCP1** | | | | | |
| Unadjusted | S1P | 0.253 | 0.085 | 0.140 | 0.003 |
| Age and sex adjusted | S1P | 0.191 | 0.079 | 0.106 | 0.016 |
| | Age | 0.025 | 0.003 | 0.361 | 0.000 |
| | Sex | -0.149 | 0.048 | -0.138 | 0.002 |
| **CD244** | | | | | |
| **Unadjusted** | **S1P** | **0.266** | **0.063** | **0.196** | **3.32x10-5** |
| **Age and sex adjusted** | **S1P** | **0.277** | **0.064** | **0.204** | **1.77x10-5** |
| | **Age** | **-0.003** | **0.002** | **-0.056** | **0.233** |
| | **Sex** | **0.039** | **0.038** | **0.048** | **0.306** |
| **IL7** | | | | | |
| **Unadjusted** | **S1P** | **0.779** | **0.095** | **0.364** | **2.37x10-15** |
| **Age and sex adjusted** | **S1P** | **0.799** | **0.095** | **0.374** | **7.56x10-16** |
| | **Age** | **0.000** | **0.004** | **-0.006** | **0.896** |
| | **Sex** | **0.102** | **0.057** | **0.079** | **0.076** |
| **LAPTGF-beta-1** | | | | | |
| **Unadjusted** | **S1P** | **0.538** | **0.092** | **0.269** | **8.75x10-9** |
| **Age and sex adjusted** | **S1P** | **0.546** | **0.093** | **0.272** | **7.75x10-9** |

| | | | | | |
|---|---|---|---|---|---|
| | **Age** | **-0.003** | **0.004** | **-0.034** | **0.455** |
| | **Sex** | **0.020** | **0.056** | **0.017** | **0.717** |
| **uPA** | | | | | |
| Unadjusted | S1P | 0.104 | 0.052 | 0.094 | 0.047 |
| Age and sex adjusted | S1P | 0.111 | 0.052 | 0.101 | 0.034 |
| | Age | 0.004 | 0.002 | 0.085 | 0.072 |
| | Sex | 0.063 | 0.031 | 0.096 | 0.045 |
| **MCP1** | | | | | |
| Unadjusted | S1P | 0.096 | 0.062 | 0.073 | 0.123 |
| Age and sex adjusted | S1P | 0.073 | 0.062 | 0.056 | 0.239 |
| | Age | 0.008 | 0.002 | 0.166 | 0.000 |
| | Sex | -0.063 | 0.037 | -0.079 | 0.092 |
| **CXCL11** | | | | | |
| **Unadjusted** | **S1P** | **0.694** | **0.141** | **0.228** | **1.18x10-6** |
| **Age and sex adjusted** | **S1P** | **0.752** | **0.139** | **0.248** | **9.92x10-8** |
| | **Age** | **0.009** | **0.005** | **0.078** | **0.088** |
| | **Sex** | **0.359** | **0.083** | **0.197** | **0.000** |
| **AXIN1** | | | | | |
| **Unadjusted** | **S1P** | **1.064** | **0.144** | **0.331** | **8.34x10-13** |
| **Age and sex adjusted** | **S1P** | **1.057** | **0.146** | **0.329** | **1.87x10-12** |
| | **Age** | **-0.004** | **0.006** | **-0.028** | **0.532** |
| | **Sex** | **-0.060** | **0.087** | **-0.031** | **0.491** |
| **TRAIL** | | | | | |
| Unadjusted | S1P | 0.165 | 0.057 | 0.135 | 0.004 |
| Age and sex adjusted | S1P | 0.152 | 0.058 | 0.124 | 0.009 |
| | Age | 0.004 | 0.002 | 0.078 | 0.097 |
| | Sex | -0.042 | 0.035 | -0.057 | 0.231 |
| **CXCL9** | | | | | |
| Unadjusted | S1P | -0.013 | 0.120 | -0.005 | 0.912 |
| Age and sex adjusted | S1P | -0.034 | 0.117 | -0.013 | 0.771 |
| | Age | 0.025 | 0.005 | 0.259 | 0.000 |
| | Sex | 0.059 | 0.070 | 0.039 | 0.401 |
| **CST5** | | | | | |
| Unadjusted | S1P | -0.061 | 0.072 | -0.040 | 0.399 |
| Age and sex adjusted | S1P | -0.079 | 0.072 | -0.052 | 0.269 |
| | Age | 0.010 | 0.003 | 0.177 | 0.000 |
| | Sex | -0.026 | 0.043 | -0.029 | 0.541 |
| **OSM** | | | | | |

| | | | | | |
|---|---|---|---|---|---|
| **Unadjusted** | **S1P** | **0.470** | **0.108** | **0.202** | **1.84x10-5** |
| **Age and sex adjusted** | **S1P** | **0.448** | **0.109** | **0.193** | **4.71x10-5** |
| | **Age** | **-0.003** | **0.004** | **-0.032** | **0.488** |
| | **Sex** | **-0.128** | **0.065** | **-0.092** | **0.050** |
| **CCL4** | | | | | |
| Unadjusted | S1P | 0.279 | 0.108 | 0.122 | 0.010 |
| Age and sex adjusted | S1P | 0.251 | 0.108 | 0.110 | 0.021 |
| | Age | 0.003 | 0.004 | 0.036 | 0.451 |
| | Sex | -0.119 | 0.065 | -0.087 | 0.068 |
| **CD6** | | | | | |
| Unadjusted | S1P | 0.078 | 0.079 | 0.046 | 0.329 |
| Age and sex adjusted | S1P | 0.072 | 0.080 | 0.043 | 0.369 |
| | Age | 0.008 | 0.003 | 0.120 | 0.011 |
| | Sex | 0.021 | 0.048 | 0.020 | 0.668 |
| **SCF** | | | | | |
| Unadjusted | S1P | -0.067 | 0.068 | -0.047 | 0.326 |
| Age and sex adjusted | S1P | -0.065 | 0.069 | -0.046 | 0.341 |
| | Age | 0.003 | 0.003 | 0.058 | 0.226 |
| | Sex | 0.028 | 0.041 | 0.033 | 0.495 |
| **TGF-alpha** | | | | | |
| Unadjusted | S1P | 0.058 | 0.047 | 0.058 | 0.219 |
| Age and sex adjusted | S1P | 0.064 | 0.047 | 0.065 | 0.175 |
| | Age | -0.001 | 0.002 | -0.015 | 0.757 |
| | Sex | 0.030 | 0.028 | 0.050 | 0.299 |
| **MCP-4** | | | | | |
| **Unadjusted** | **S1P** | **0.449** | **0.091** | **0.228** | **1.15x10-6** |
| **Age and sex adjusted** | **S1P** | **0.461** | **0.091** | **0.235** | **6.82x10-7** |
| | **Age** | **0.005** | **0.004** | **0.061** | **0.191** |
| | **Sex** | **0.092** | **0.055** | **0.078** | **0.094** |
| **CCL11** | | | | | |
| Unadjusted | S1P | 0.191 | 0.074 | 0.122 | 0.010 |
| Age and sex adjusted | S1P | 0.156 | 0.073 | 0.099 | 0.033 |
| | Age | 0.013 | 0.003 | 0.213 | 0.000 |
| | Sex | -0.095 | 0.044 | -0.101 | 0.030 |
| **TNFSF14** | | | | | |
| **Unadjusted** | **S1P** | **0.623** | **0.102** | **0.279** | **2.18x10-9** |
| **Age and sex adjusted** | **S1P** | **0.642** | **0.103** | **0.288** | **9.62x10-10** |
| | **Age** | **5.475x10-5** | **0.004** | **0.001** | **0.989** |

| | | | | | |
|---|---|---|---|---|---|
| | **Sex** | **0.099** | **0.062** | **0.074** | **0.109** |
| **FGF-23** | | | | | |
| Unadjusted | S1P | 0.064 | 0.070 | 0.043 | 0.362 |
| Age and sex adjusted | S1P | 0.072 | 0.070 | 0.049 | 0.308 |
| | Age | 0.003 | 0.003 | 0.047 | 0.321 |
| | Sex | 0.059 | 0.042 | 0.066 | 0.166 |
| **MMP-1** | | | | | |
| **Unadjusted** | **S1P** | **0.755** | **0.157** | **0.223** | **2.11x10-6** |
| **Age and sex adjusted** | **S1P** | **0.766** | **0.158** | **0.226** | **1.62x10-6** |
| | **Age** | **0.013** | **0.006** | **0.095** | **0.039** |
| | **Sex** | **0.139** | **0.095** | **0.068** | **0.142** |
| **LIF-R** | | | | | |
| Unadjusted | S1P | 0.078 | 0.043 | 0.086 | 0.069 |
| Age and sex adjusted | S1P | 0.083 | 0.043 | 0.092 | 0.054 |
| | Age | 0.003 | 0.002 | 0.089 | 0.061 |
| | Sex | 0.045 | 0.026 | 0.084 | 0.078 |
| **FGF-21** | | | | | |
| Unadjusted | S1P | 0.689 | 0.227 | 0.143 | 0.003 |
| Age and sex adjusted | S1P | 0.606 | 0.226 | 0.126 | 0.007 |
| | Age | 0.034 | 0.009 | 0.180 | 0.000 |
| | Sex | -0.194 | 0.135 | -0.067 | 0.153 |
| **CCL19** | | | | | |
| Unadjusted | S1P | 0.007 | 0.112 | 0.003 | 0.949 |
| Age and sex adjusted | S1P | -0.015 | 0.113 | -0.006 | 0.894 |
| | Age | 0.006 | 0.004 | 0.070 | 0.140 |
| | Sex | -0.072 | 0.068 | -0.051 | 0.288 |
| **IL-10RB** | | | | | |
| Unadjusted | S1P | 0.132 | 0.057 | 0.110 | 0.020 |
| Age and sex adjusted | S1P | 0.122 | 0.057 | 0.101 | 0.034 |
| | Age | 0.004 | 0.002 | 0.096 | 0.043 |
| | Sex | -0.025 | 0.034 | -0.035 | 0.463 |
| **IL-18R1** | | | | | |
| **Unadjusted** | **S1P** | **0.277** | **0.070** | **0.185** | **8.59x10-5** |
| **Age and sex adjusted** | **S1P** | **0.257** | **0.070** | **0.172** | **2.84x10-4** |
| | **Age** | **0.005** | **0.003** | **0.085** | **0.070** |
| | **Sex** | **-0.072** | **0.042** | **-0.080** | **0.088** |
| **PD-L1** | | | | | |
| Unadjusted | S1P | 0.116 | 0.072 | 0.076 | 0.109 |

| | | | | | |
|---|---|---|---|---|---|
| Age and sex adjusted | S1P | 0.073 | 0.071 | 0.048 | 0.303 |
| | Age | 0.000 | 0.003 | 0.006 | 0.901 |
| | Sex | -0.216 | 0.043 | -0.236 | 0.000 |
| **Beta-NGF** | | | | | |
| Unadjusted | S1P | 0.077 | 0.057 | 0.063 | 0.183 |
| Age and sex adjusted | S1P | 0.073 | 0.058 | 0.060 | 0.209 |
| | Age | 0.002 | 0.002 | 0.040 | 0.403 |
| | Sex | -0.006 | 0.035 | -0.009 | 0.854 |
| **CXCL5** | | | | | |
| **Unadjusted** | **S1P** | **0.767** | **0.134** | **0.262** | **2.12x10-8** |
| **Age and sex adjusted** | **S1P** | **0.853** | **0.131** | **0.291** | **1.96x10-10** |
| | **Age** | **0.002** | **0.005** | **0.018** | **0.681** |
| | **Sex** | **0.454** | **0.079** | **0.258** | **0.000** |
| **TRANCE** | | | | | |
| Unadjusted | S1P | 0.121 | 0.098 | 0.059 | 0.215 |
| Age and sex adjusted | S1P | 0.115 | 0.098 | 0.056 | 0.243 |
| | Age | -0.007 | 0.004 | -0.091 | 0.055 |
| | Sex | -0.081 | 0.059 | -0.066 | 0.170 |
| **HGF** | | | | | |
| **Unadjusted** | **S1P** | **0.405** | **0.066** | **0.279** | **2.31x10-9** |
| **Age and sex adjusted** | **S1P** | **0.377** | **0.066** | **0.259** | **1.98x10-8** |
| | **Age** | **0.009** | **0.003** | **0.156** | **0.001** |
| | **Sex** | **-0.087** | **0.040** | **-0.100** | **0.028** |
| **IL-12B** | | | | | |
| Unadjusted | S1P | -0.150 | 0.091 | -0.078 | 0.100 |
| Age and sex adjusted | S1P | -0.129 | 0.092 | -0.067 | 0.160 |
| | Age | 0.003 | 0.004 | 0.037 | 0.436 |
| | Sex | 0.129 | 0.055 | 0.112 | 0.019 |
| **MMP-10** | | | | | |
| Unadjusted | S1P | 0.117 | 0.102 | 0.055 | 0.249 |
| Age and sex adjusted | S1P | 0.119 | 0.102 | 0.056 | 0.244 |
| | Age | 0.006 | 0.004 | 0.069 | 0.147 |
| | Sex | 0.047 | 0.061 | 0.037 | 0.440 |
| **IL10** | | | | | |
| Unadjusted | S1P | 0.147 | 0.082 | 0.087 | 0.073 |
| Age and sex adjusted | S1P | 0.138 | 0.082 | 0.081 | 0.094 |
| | Age | -0.004 | 0.003 | -0.055 | 0.258 |
| | Sex | -0.072 | 0.050 | -0.070 | 0.150 |

| | | | | | |
|---|---|---|---|---|---|
| **CCL23** | | | | | |
| Unadjusted | S1P | 0.047 | 0.078 | 0.029 | 0.543 |
| Age and sex adjusted | S1P | 0.048 | 0.078 | 0.030 | 0.536 |
| | Age | 0.006 | 0.003 | 0.092 | 0.054 |
| | Sex | 0.043 | 0.047 | 0.044 | 0.359 |
| **CD5** | | | | | |
| Unadjusted | S1P | -0.017 | 0.056 | -0.015 | 0.755 |
| Age and sex adjusted | S1P | -0.009 | 0.056 | -0.008 | 0.871 |
| | Age | 0.002 | 0.002 | 0.046 | 0.334 |
| | Sex | 0.056 | 0.034 | 0.080 | 0.097 |
| **CCL3** | | | | | |
| Unadjusted | S1P | 0.150 | 0.090 | 0.079 | 0.098 |
| Age and sex adjusted | S1P | 0.112 | 0.090 | 0.059 | 0.216 |
| | Age | 0.008 | 0.003 | 0.111 | 0.019 |
| | Sex | -0.141 | 0.054 | -0.123 | 0.009 |
| **Flt3L** | | | | | |
| Unadjusted | S1P | 0.064 | 0.065 | 0.047 | 0.325 |
| Age and sex adjusted | S1P | 0.064 | 0.063 | 0.047 | 0.311 |
| | Age | 0.013 | 0.002 | 0.252 | 0.000 |
| | Sex | 0.088 | 0.038 | 0.106 | 0.022 |
| **CXCL6** | | | | | |
| **Unadjusted** | **S1P** | **0.726** | **0.119** | **0.279** | **2.33x10-9** |
| **Age and sex adjusted** | **S1P** | **0.752** | **0.120** | **0.288** | **8.78x10-10** |
| | **Age** | **-0.002** | **0.005** | **-0.022** | **0.628** |
| | **Sex** | **0.116** | **0.072** | **0.074** | **0.108** |
| **CXCL10** | | | | | |
| Unadjusted | S1P | 0.096 | 0.140 | 0.033 | 0.493 |
| Age and sex adjusted | S1P | 0.065 | 0.141 | 0.022 | 0.646 |
| | Age | 0.011 | 0.005 | 0.092 | 0.053 |
| | Sex | -0.092 | 0.084 | -0.052 | 0.276 |
| **4EBP1** | | | | | |
| **Unadjusted** | **S1P** | **1.049** | **0.184** | **0.262** | **2.23x10-8** |
| **Age and sex adjusted** | **S1P** | **1.035** | **0.186** | **0.258** | **4.28x10-8** |
| | **Age** | **0.011** | **0.007** | **0.072** | **0.119** |
| | **Sex** | **0.002** | **0.111** | **0.001** | **0.988** |
| **SIRT2** | | | | | |
| **Unadjusted** | **S1P** | **1.086** | **0.130** | **0.370** | **7.09x10-16** |
| **Age and sex adjusted** | **S1P** | **1.138** | **0.129** | **0.388** | **3.51x10-17** |

| | | | | | |
|---|---|---|---|---|---|
| | **Age** | **-0.003** | **0.005** | **-0.023** | **0.597** |
| | **Sex** | **0.246** | **0.078** | **0.139** | **0.002** |
| **CCL28** | | | | | |
| Unadjusted | S1P | 0.097 | 0.045 | 0.103 | 0.032 |
| Age and sex adjusted | S1P | 0.102 | 0.045 | 0.108 | 0.025 |
| | Age | 0.004 | 0.002 | 0.109 | 0.022 |
| | Sex | 0.042 | 0.027 | 0.075 | 0.120 |
| **DNER** | | | | | |
| Unadjusted | S1P | 0.042 | 0.044 | 0.046 | 0.335 |
| Age and sex adjusted | S1P | 0.032 | 0.044 | 0.035 | 0.462 |
| | Age | 0.000 | 0.002 | -0.009 | 0.854 |
| | Sex | -0.052 | 0.026 | -0.095 | 0.047 |
| **EN-RAGE** | | | | | |
| Unadjusted | S1P | 0.163 | 0.089 | 0.087 | 0.067 |
| Age and sex adjusted | S1P | 0.140 | 0.089 | 0.075 | 0.118 |
| | Age | 0.006 | 0.003 | 0.083 | 0.081 |
| | Sex | -0.086 | 0.053 | -0.077 | 0.109 |
| **CD40** | | | | | |
| **Unadjusted** | **S1P** | **0.359** | **0.074** | **0.224** | **1.86x10-6** |
| **Age and sex adjusted** | **S1P** | **0.364** | **0.075** | **0.227** | **1.69x10-6** |
| | **Age** | **0.000** | **0.003** | **0.005** | **0.909** |
| | **Sex** | **0.028** | **0.045** | **0.029** | **0.531** |
| **FGF-19** | | | | | |
| Unadjusted | S1P | 0.163 | 0.141 | 0.055 | 0.250 |
| Age and sex adjusted | S1P | 0.199 | 0.142 | 0.067 | 0.161 |
| | Age | -0.005 | 0.005 | -0.044 | 0.356 |
| | Sex | 0.154 | 0.085 | 0.086 | 0.071 |
| **MCP-2** | | | | | |
| Unadjusted | S1P | 0.420 | 0.111 | 0.178 | 1.69x10-4 |
| Age and sex adjusted | S1P | 0.415 | 0.112 | 0.175 | 2.38x10-4 |
| | Age | 0.001 | 0.004 | 0.013 | 0.790 |
| | Sex | -0.020 | 0.067 | -0.014 | 0.761 |
| **CASP-8** | | | | | |
| **Unadjusted** | **S1P** | **0.448** | **0.099** | **0.210** | **8.45x10-6** |
| **Age and sex adjusted** | **S1P** | **0.455** | **0.101** | **0.213** | **7.72x10-6** |
| | **Age** | **-0.003** | **0.004** | **-0.032** | **0.489** |
| | **Sex** | **0.016** | **0.060** | **0.013** | **0.788** |
| **CCL25** | | | | | |

| | | | | | |
|---|---|---|---|---|---|
| Unadjusted | S1P | 0.187 | 0.091 | 0.097 | 0.041 |
| Age and sex adjusted | S1P | 0.172 | 0.091 | 0.090 | 0.060 |
| | Age | 0.009 | 0.004 | 0.116 | 0.014 |
| | Sex | -0.016 | 0.055 | -0.014 | 0.765 |
| **CX3CL1** | | | | | |
| Unadjusted | S1P | -0.064 | 0.068 | -0.044 | 0.350 |
| Age and sex adjusted | S1P | -0.052 | 0.068 | -0.036 | 0.447 |
| | Age | 0.003 | 0.003 | 0.054 | 0.256 |
| | Sex | 0.079 | 0.041 | 0.091 | 0.056 |
| **TNFRSF9** | | | | | |
| Unadjusted | S1P | 0.052 | 0.068 | 0.037 | 0.443 |
| Age and sex adjusted | S1P | 0.033 | 0.068 | 0.023 | 0.623 |
| | Age | 0.006 | 0.003 | 0.115 | 0.015 |
| | Sex | -0.054 | 0.041 | -0.063 | 0.188 |
| **TWEAK** | | | | | |
| Unadjusted | S1P | 0.153 | 0.058 | 0.125 | 0.008 |
| Age and sex adjusted | S1P | 0.170 | 0.058 | 0.139 | 0.003 |
| | Age | 0.000 | 0.002 | 0.005 | 0.921 |
| | Sex | 0.089 | 0.035 | 0.121 | 0.011 |
| **CCL20** | | | | | |
| Unadjusted | S1P | 0.173 | 0.154 | 0.053 | 0.261 |
| Age and sex adjusted | S1P | 0.175 | 0.155 | 0.054 | 0.260 |
| | Age | -0.003 | 0.006 | -0.020 | 0.669 |
| | Sex | -0.005 | 0.093 | -0.003 | 0.954 |
| **ST1A1** | | | | | |
| Unadjusted | S1P | 0.227 | 0.127 | 0.085 | 0.073 |
| Age and sex adjusted | S1P | 0.255 | 0.127 | 0.096 | 0.046 |
| | Age | -0.004 | 0.005 | -0.034 | 0.468 |
| | Sex | 0.119 | 0.076 | 0.074 | 0.121 |
| **STAMPB** | | | | | |
| **Unadjusted** | **S1P** | **1.010** | **0.124** | **0.361** | **3.95x10-15** |
| **Age and sex adjusted** | **S1P** | **1.031** | **0.125** | **0.369** | **1.93x10-15** |
| | **Age** | **-0.003** | **0.005** | **-0.030** | **0.506** |
| | **Sex** | **0.088** | **0.075** | **0.052** | **0.242** |
| **ADA** | | | | | |
| **Unadjusted** | **S1P** | **0.371** | **0.064** | **0.266** | **1.30x10-8** |
| **Age and sex adjusted** | **S1P** | **0.372** | **0.065** | **0.267** | **1.55x10-8** |
| | **Age** | **-0.003** | **0.002** | **-0.056** | **0.223** |

| | | | | | |
|---|---|---|---|---|---|
| | **Sex** | **-0.013** | **0.039** | **-0.016** | **0.728** |
| **TNFB** | | | | | |
| Unadjusted | S1P | -0.062 | 0.071 | -0.041 | 0.384 |
| Age and sex adjusted | S1P | -0.033 | 0.071 | -0.022 | 0.647 |
| | Age | -0.002 | 0.003 | -0.029 | 0.534 |
| | Sex | 0.140 | 0.043 | 0.155 | 0.001 |
| **CSF-1** | | | | | |
| Unadjusted | S1P | 0.032 | 0.043 | 0.036 | 0.451 |
| Age and sex adjusted | S1P | 0.035 | 0.043 | 0.039 | 0.414 |
| | Age | 0.002 | 0.002 | 0.071 | 0.138 |
| | Sex | 0.031 | 0.026 | 0.057 | 0.235 |

METABOLISM PANEL

| Model | | Unstandardized Coefficients | | Standardized Coefficients | p |
|---|---|---|---|---|---|
| | | β | Std. error | Beta | |
| **CLMP** | | | | | |
| Unadjusted | S1P | 0.000 | 0.089 | 0.000 | 0.996 |
| Age and sex adjusted | S1P | 0.029 | 0.087 | 0.015 | 0.744 |
| | Age | 0.010 | 0.003 | 0.137 | 0.003 |
| | Sex | 0.203 | 0.052 | 0.181 | 0.000 |
| **LRIG1** | | | | | |
| Unadjusted | S1P | 0.063 | 0.072 | 0.042 | 0.378 |
| Age and sex adjusted | S1P | 0.067 | 0.072 | 0.044 | 0.352 |
| | Age | 0.001 | 0.003 | 0.015 | 0.745 |
| | Sex | 0.027 | 0.043 | 0.030 | 0.534 |
| **NPTXR** | | | | | |
| Unadjusted | S1P | 0.077 | 0.079 | 0.046 | 0.334 |
| Age and sex adjusted | S1P | 0.088 | 0.080 | 0.052 | 0.270 |
| | Age | 0.003 | 0.003 | 0.050 | 0.291 |
| | Sex | 0.078 | 0.048 | 0.077 | 0.104 |
| **THOP1** | | | | | |
| **Unadjusted** | **S1P** | **0.353** | **0.075** | **0.216** | **3.75x10-6** |
| **Age and sex adjusted** | **S1P** | **0.315** | **0.075** | **0.193** | **2.86x10-5** |
| | **Age** | **-0.001** | **0.003** | **-0.009** | **0.848** |
| | **Sex** | **-0.197** | **0.045** | **-0.202** | **0.000** |
| **CTSO** | | | | | |
| Unadjusted | S1P | 0.182 | 0.064 | 0.134 | 0.004 |
| Age and sex adjusted | S1P | 0.155 | 0.063 | 0.114 | 0.015 |
| | Age | 0.000 | 0.002 | -0.003 | 0.955 |
| | Sex | -0.142 | 0.038 | -0.174 | 0.000 |

| | | | | | |
|---|---|---|---|---|---|
| **FCRL1** | | | | | |
| Unadjusted | S1P | -0.015 | 0.089 | -0.008 | 0.863 |
| Age and sex adjusted | S1P | -0.015 | 0.090 | -0.008 | 0.871 |
| | Age | 0.000 | 0.003 | -0.005 | 0.915 |
| | Sex | 0.002 | 0.054 | 0.002 | 0.975 |
| **CD164** | | | | | |
| **Unadjusted** | **S1P** | **0.296** | **0.071** | **0.192** | **3.89x10-5** |
| **Age and sex adjusted** | **S1P** | **0.291** | **0.072** | **0.189** | **6.36x10-5** |
| | **Age** | **0.000** | **0.003** | **0.005** | **0.921** |
| | **Sex** | **-0.027** | **0.043** | **-0.029** | **0.535** |
| **DDC** | | | | | |
| Unadjusted | S1P | 0.148 | 0.097 | 0.072 | 0.127 |
| Age and sex adjusted | S1P | 0.110 | 0.097 | 0.054 | 0.256 |
| | Age | 0.005 | 0.004 | 0.060 | 0.198 |
| | Sex | -0.166 | 0.058 | -0.135 | 0.004 |
| **ACP6** | | | | | |
| Unadjusted | S1P | 0.348 | 0.099 | 0.163 | 4.95x10-4 |
| Age and sex adjusted | S1P | 0.343 | 0.100 | 0.161 | 6.62x10-4 |
| | Age | -0.001 | 0.004 | -0.007 | 0.878 |
| | Sex | -0.027 | 0.060 | -0.021 | 0.657 |
| **TFF2** | | | | | |
| Unadjusted | S1P | -0.106 | 0.111 | -0.045 | 0.340 |
| Age and sex adjusted | S1P | -0.098 | 0.107 | -0.042 | 0.361 |
| | Age | 0.022 | 0.004 | 0.247 | 0.000 |
| | Sex | 0.175 | 0.064 | 0.125 | 0.007 |
| **ANGPT2** | | | | | |
| Unadjusted | S1P | 0.052 | 0.067 | 0.037 | 0.433 |
| Age and sex adjusted | S1P | 0.068 | 0.067 | 0.048 | 0.315 |
| | Age | 0.001 | 0.003 | 0.017 | 0.717 |
| | Sex | 0.082 | 0.040 | 0.097 | 0.042 |
| **CD2AP** | | | | | |
| **Unadjusted** | **S1P** | **0.954** | **0.168** | **0.259** | **2.38x10-8** |
| **Age and sex adjusted** | **S1P** | **0.950** | **0.169** | **0.258** | **3.41x10-8** |
| | **Age** | **0.010** | **0.006** | **0.069** | **0.129** |
| | **Sex** | **0.039** | **0.101** | **0.018** | **0.698** |
| **ANGPTL7** | | | | | |
| Unadjusted | S1P | 0.031 | 0.052 | 0.029 | 0.556 |
| Age and sex adjusted | S1P | 0.007 | 0.052 | 0.007 | 0.894 |
| | Age | 0.003 | 0.002 | 0.061 | 0.209 |
| | Sex | -0.105 | 0.032 | -0.163 | 0.001 |
| **CLEC5A** | | | | | |

| | | | | | |
|---|---|---|---|---|---|
| Unadjusted | S1P | 0.024 | 0.060 | 0.019 | 0.694 |
| Age and sex adjusted | S1P | 0.039 | 0.060 | 0.031 | 0.517 |
| | Age | 0.003 | 0.002 | 0.060 | 0.198 |
| | Sex | 0.095 | 0.036 | 0.126 | 0.008 |
| **TINAGL1** | | | | | |
| Unadjusted | S1P | -0.007 | 0.065 | -0.005 | 0.909 |
| Age and sex adjusted | S1P | -0.008 | 0.065 | -0.006 | 0.899 |
| | Age | 0.003 | 0.003 | 0.058 | 0.221 |
| | Sex | 0.014 | 0.039 | 0.017 | 0.726 |
| **ENO2** | | | | | |
| **Unadjusted** | **S1P** | **0.654** | **0.102** | **0.289** | **4.07x10-10** |
| **Age and sex adjusted** | **S1P** | **0.642** | **0.103** | **0.284** | **1.12x10-9** |
| | **Age** | **-0.001** | **0.004** | **-0.017** | **0.706** |
| | **Sex** | **-0.069** | **0.062** | **-0.051** | **0.261** |
| **NADK** | | | | | |
| Unadjusted | S1P | 0.218 | 0.079 | 0.129 | 0.006 |
| Age and sex adjusted | S1P | 0.162 | 0.077 | 0.096 | 0.036 |
| | Age | 0.009 | 0.003 | 0.139 | 0.002 |
| | Sex | -0.230 | 0.046 | -0.227 | 0.000 |
| **GHRL** | | | | | |
| Unadjusted | S1P | -0.095 | 0.157 | -0.029 | 0.543 |
| Age and sex adjusted | S1P | 0.010 | 0.149 | 0.003 | 0.949 |
| | Age | 0.016 | 0.006 | 0.127 | 0.004 |
| | Sex | 0.636 | 0.089 | 0.320 | 0.000 |
| **SERPINB6** | | | | | |
| **Unadjusted** | **S1P** | **0.624** | **0.088** | **0.316** | **6.41x10-12** |
| **Age and sex adjusted** | **S1P** | **0.628** | **0.089** | **0.318** | **7.80x10-12** |
| | **Age** | **-0.002** | **0.003** | **-0.025** | **0.580** |
| | **Sex** | **0.007** | **0.053** | **0.006** | **0.898** |
| **CDHR5** | | | | | |
| Unadjusted | S1P | 0.055 | 0.068 | 0.038 | 0.419 |
| Age and sex adjusted | S1P | 0.051 | 0.068 | 0.035 | 0.458 |
| | Age | -0.005 | 0.003 | -0.083 | 0.078 |
| | Sex | -0.049 | 0.041 | -0.057 | 0.228 |
| **CCDC80** | | | | | |
| Unadjusted | S1P | 0.170 | 0.091 | 0.088 | 0.063 |
| Age and sex adjusted | S1P | 0.153 | 0.091 | 0.079 | 0.094 |
| | Age | 0.011 | 0.003 | 0.152 | 0.001 |
| | Sex | -0.018 | 0.054 | -0.016 | 0.739 |
| **CA13** | | | | | |
| **Unadjusted** | **S1P** | **0.927** | **0.128** | **0.325** | **1.59x10-12** |

| | | | | | |
|---|---|---|---|---|---|
| **Age and sex adjusted** | **S1P** | **0.935** | **0.129** | **0.327** | **1.71x10-12** |
| | **Age** | **-0.005** | **0.005** | **-0.043** | **0.336** |
| | **Sex** | **0.008** | **0.077** | **0.005** | **0.913** |
| **SEMA3F** | | | | | |
| Unadjusted | S1P | 0.005 | 0.055 | 0.004 | 0.930 |
| Age and sex adjusted | S1P | -0.013 | 0.055 | -0.011 | 0.813 |
| | Age | 0.002 | 0.002 | 0.042 | 0.377 |
| | Sex | -0.080 | 0.033 | -0.115 | 0.015 |
| **KLK10** | | | | | |
| Unadjusted | S1P | -0.101 | 0.072 | -0.067 | 0.164 |
| Age and sex adjusted | S1P | -0.106 | 0.073 | -0.070 | 0.149 |
| | Age | -0.001 | 0.003 | -0.017 | 0.726 |
| | Sex | -0.029 | 0.044 | -0.032 | 0.512 |
| **PILRB** | | | | | |
| Unadjusted | S1P | 0.032 | 0.104 | 0.014 | 0.761 |
| Age and sex adjusted | S1P | -0.002 | 0.104 | -0.001 | 0.988 |
| | Age | 0.006 | 0.004 | 0.067 | 0.154 |
| | Sex | -0.135 | 0.062 | -0.103 | 0.030 |
| **ANGPTL1** | | | | | |
| Unadjusted | S1P | 0.022 | 0.058 | 0.018 | 0.702 |
| Age and sex adjusted | S1P | 0.028 | 0.058 | 0.023 | 0.632 |
| | Age | 0.004 | 0.002 | 0.079 | 0.094 |
| | Sex | 0.052 | 0.035 | 0.071 | 0.133 |
| **APLP1** | | | | | |
| Unadjusted | S1P | -0.314 | 0.122 | -0.121 | 0.010 |
| Age and sex adjusted | S1P | -0.283 | 0.123 | -0.109 | 0.021 |
| | Age | 0.003 | 0.005 | 0.025 | 0.594 |
| | Sex | 0.175 | 0.073 | 0.112 | 0.017 |
| **ADGRG2** | | | | | |
| Unadjusted | S1P | -0.007 | 0.060 | -0.006 | 0.904 |
| Age and sex adjusted | S1P | 0.031 | 0.059 | 0.024 | 0.605 |
| | Age | -0.005 | 0.002 | -0.111 | 0.016 |
| | Sex | 0.160 | 0.035 | 0.211 | 0.000 |
| **TYMP** | | | | | |
| Unadjusted | S1P | 0.271 | 0.165 | 0.077 | 0.101 |
| Age and sex adjusted | S1P | 0.255 | 0.164 | 0.073 | 0.122 |
| | Age | -0.017 | 0.006 | -0.126 | 0.007 |
| | Sex | -0.185 | 0.098 | -0.088 | 0.060 |
| **GRAP2** | | | | | |
| **Unadjusted** | **S1P** | **1.427** | **0.217** | **0.296** | **1.33x10-10** |
| **Age and sex adjusted** | **S1P** | **1.408** | **0.218** | **0.292** | **2.90x10-10** |

| | | | | | |
|---|---|---|---|---|---|
| | **Age** | **0.016** | **0.008** | **0.087** | **0.053** |
| | **Sex** | **0.001** | **0.130** | **0.000** | **0.993** |
| **LILRA5** | | | | | |
| Unadjusted | S1P | 0.221 | 0.072 | 0.144 | 0.002 |
| Age and sex adjusted | S1P | 0.205 | 0.072 | 0.133 | 0.005 |
| | Age | 0.003 | 0.003 | 0.048 | 0.306 |
| | Sex | -0.065 | 0.043 | -0.071 | 0.133 |
| **ALDH1A1** | | | | | |
| Unadjusted | S1P | -0.008 | 0.113 | -0.004 | 0.942 |
| Age and sex adjusted | S1P | -0.045 | 0.113 | -0.019 | 0.692 |
| | Age | -0.003 | 0.004 | -0.036 | 0.464 |
| | Sex | -0.207 | 0.068 | -0.149 | 0.002 |
| **CD79B** | | | | | |
| Unadjusted | S1P | 0.033 | 0.063 | 0.024 | 0.605 |
| Age and sex adjusted | S1P | 0.012 | 0.063 | 0.009 | 0.845 |
| | Age | 0.000 | 0.002 | -0.006 | 0.904 |
| | Sex | -0.106 | 0.038 | -0.132 | 0.005 |
| **ANXA11** | | | | | |
| **Unadjusted** | **S1P** | **0.932** | **0.134** | **0.313** | **1.48x10-11** |
| **Age and sex adjusted** | **S1P** | **0.937** | **0.136** | **0.315** | **1.86x10-11** |
| | **Age** | **-0.005** | **0.005** | **-0.047** | **0.299** |
| | **Sex** | **-0.011** | **0.081** | **-0.006** | **0.896** |
| **SIGLEC7** | | | | | |
| Unadjusted | S1P | 0.114 | 0.065 | 0.083 | 0.080 |
| Age and sex adjusted | S1P | 0.069 | 0.063 | 0.050 | 0.275 |
| | Age | 0.008 | 0.002 | 0.150 | 0.001 |
| | Sex | -0.181 | 0.038 | -0.220 | 0.000 |
| **QDPR** | | | | | |
| **Unadjusted** | **S1P** | **0.621** | **0.086** | **0.324** | **1.78x10-12** |
| **Age and sex adjusted** | **S1P** | **0.615** | **0.086** | **0.320** | **4.41x10-12** |
| | **Age** | **-0.003** | **0.003** | **-0.038** | **0.394** |
| | **Sex** | **-0.051** | **0.052** | **-0.045** | **0.320** |
| **SNAP23** | | | | | |
| **Unadjusted** | **S1P** | **0.984** | **0.148** | **0.299** | **9.46x10-11** |
| **Age and sex adjusted** | **S1P** | **0.973** | **0.150** | **0.295** | **2.14x10-10** |
| | **Age** | **0.007** | **0.006** | **0.055** | **0.227** |
| | **Sex** | **-0.016** | **0.089** | **-0.008** | **0.862** |
| **CLSTN2** | | | | | |
| Unadjusted | S1P | 0.141 | 0.069 | 0.096 | 0.042 |
| Age and sex adjusted | S1P | 0.122 | 0.070 | 0.083 | 0.080 |
| | Age | 0.005 | 0.003 | 0.089 | 0.059 |

| | | | | | |
|---|---|---|---|---|---|
| | Sex | -0.066 | 0.042 | -0.075 | 0.112 |
| **COMT** | | | | | |
| **Unadjusted** | **S1P** | **0.901** | **0.124** | **0.325** | **1.56x10-12** |
| **Age and sex adjusted** | **S1P** | **0.898** | **0.125** | **0.324** | **2.87x10-12** |
| | **Age** | **0.004** | **0.005** | **0.039** | **0.385** |
| | **Sex** | **0.011** | **0.075** | **0.006** | **0.887** |
| **CLUL1** | | | | | |
| Unadjusted | S1P | -0.038 | 0.094 | -0.019 | 0.689 |
| Age and sex adjusted | S1P | 0.002 | 0.094 | 0.001 | 0.983 |
| | Age | 0.002 | 0.004 | 0.026 | 0.582 |
| | Sex | 0.215 | 0.056 | 0.181 | 0.000 |
| **CHRDL2** | | | | | |
| Unadjusted | S1P | 0.199 | 0.110 | 0.085 | 0.072 |
| Age and sex adjusted | S1P | 0.230 | 0.111 | 0.098 | 0.038 |
| | Age | -0.010 | 0.004 | -0.116 | 0.014 |
| | Sex | 0.093 | 0.066 | 0.066 | 0.159 |
| **NOMO1** | | | | | |
| Unadjusted | S1P | 0.229 | 0.070 | 0.153 | 0.001 |
| Age and sex adjusted | S1P | 0.190 | 0.069 | 0.127 | 0.006 |
| | Age | 0.002 | 0.003 | 0.029 | 0.531 |
| | Sex | -0.193 | 0.041 | -0.215 | 0.000 |
| **SOST** | | | | | |
| Unadjusted | S1P | 0.064 | 0.085 | 0.035 | 0.452 |
| Age and sex adjusted | S1P | 0.022 | 0.084 | 0.012 | 0.793 |
| | Age | 0.010 | 0.003 | 0.152 | 0.001 |
| | Sex | -0.149 | 0.050 | -0.139 | 0.003 |
| **FAM3C** | | | | | |
| Unadjusted | S1P | 0.041 | 0.076 | 0.025 | 0.592 |
| Age and sex adjusted | S1P | 0.005 | 0.075 | 0.003 | 0.945 |
| | Age | 0.007 | 0.003 | 0.115 | 0.014 |
| | Sex | -0.138 | 0.045 | -0.144 | 0.002 |
| **TXNDC5** | | | | | |
| **Unadjusted** | **S1P** | **0.700** | **0.114** | **0.279** | **1.97x10-9** |
| **Age and sex adjusted** | **S1P** | **0.699** | **0.115** | **0.278** | **2.99x10-9** |
| | **Age** | **0.001** | **0.004** | **0.012** | **0.785** |
| | **Sex** | **0.003** | **0.069** | **0.002** | **0.960** |
| **PPP1R2** | | | | | |
| **Unadjusted** | **S1P** | **0.902** | **0.124** | **0.324** | **1.70x10-12** |
| **Age and sex adjusted** | **S1P** | **0.895** | **0.125** | **0.322** | **3.91x10-12** |
| | **Age** | **0.000** | **0.005** | **-0.003** | **0.950** |
| | **Sex** | **-0.036** | **0.075** | **-0.022** | **0.629** |

| | | | | | |
|---|---|---|---|---|---|
| **LRP11** | | | | | |
| Unadjusted | S1P | 0.025 | 0.079 | 0.015 | 0.752 |
| Age and sex adjusted | S1P | 0.002 | 0.079 | 0.001 | 0.978 |
| | Age | 0.004 | 0.003 | 0.059 | 0.214 |
| | Sex | -0.094 | 0.047 | -0.094 | 0.048 |
| **ADGRE2** | | | | | |
| Unadjusted | S1P | 0.072 | 0.070 | 0.048 | 0.306 |
| Age and sex adjusted | S1P | 0.032 | 0.069 | 0.022 | 0.641 |
| | Age | 0.008 | 0.003 | 0.138 | 0.003 |
| | Sex | -0.155 | 0.041 | -0.174 | 0.000 |
| **ENPP7** | | | | | |
| Unadjusted | S1P | 0.529 | 0.140 | 0.175 | 1.84x10-4 |
| Age and sex adjusted | S1P | 0.479 | 0.140 | 0.158 | 0.001 |
| | Age | 0.010 | 0.005 | 0.088 | 0.058 |
| | Sex | -0.197 | 0.084 | -0.109 | 0.019 |
| **MCFD2** | | | | | |
| **Unadjusted** | **S1P** | **0.478** | **0.080** | **0.272** | **4.10x10-9** |
| **Age and sex adjusted** | **S1P** | **0.484** | **0.080** | **0.276** | **3.81x10-9** |
| | **Age** | **-0.001** | **0.003** | **-0.016** | **0.722** |
| | **Sex** | **0.023** | **0.048** | **0.022** | **0.631** |
| **REG4** | | | | | |
| Unadjusted | S1P | 0.003 | 0.102 | 0.001 | 0.978 |
| Age and sex adjusted | S1P | -0.023 | 0.101 | -0.011 | 0.822 |
| | Age | 0.013 | 0.004 | 0.152 | 0.001 |
| | Sex | -0.054 | 0.061 | -0.042 | 0.371 |
| **SUMF2** | | | | | |
| **Unadjusted** | **S1P** | **0.577** | **0.104** | **0.254** | **4.64x10-8** |
| **Age and sex adjusted** | **S1P** | **0.542** | **0.104** | **0.239** | **2.68x10-7** |
| | **Age** | **-0.001** | **0.004** | **-0.014** | **0.765** |
| | **Sex** | **-0.184** | **0.062** | **-0.136** | **0.003** |
| **CANT1** | | | | | |
| Unadjusted | S1P | 0.099 | 0.057 | 0.082 | 0.080 |
| Age and sex adjusted | S1P | 0.085 | 0.057 | 0.070 | 0.137 |
| | Age | 0.001 | 0.002 | 0.012 | 0.792 |
| | Sex | -0.070 | 0.034 | -0.098 | 0.039 |
| **CD1C** | | | | | |
| Unadjusted | S1P | -0.013 | 0.065 | -0.009 | 0.842 |
| Age and sex adjusted | S1P | 0.005 | 0.065 | 0.004 | 0.938 |
| | Age | -0.006 | 0.003 | -0.116 | 0.014 |
| | Sex | 0.055 | 0.039 | 0.067 | 0.159 |
| **GAL** | | | | | |

| | | | | | |
|---|---|---|---|---|---|
| Unadjusted | S1P | 0.067 | 0.142 | 0.022 | 0.638 |
| Age and sex adjusted | S1P | 0.036 | 0.140 | 0.012 | 0.800 |
| | Age | -0.019 | 0.005 | -0.159 | 0.001 |
| | Sex | -0.274 | 0.084 | -0.152 | 0.001 |
| **CDH2** | | | | | |
| Unadjusted | S1P | 0.231 | 0.082 | 0.134 | 0.005 |
| Age and sex adjusted | S1P | 0.168 | 0.078 | 0.097 | 0.032 |
| | Age | 0.012 | 0.003 | 0.177 | 0.000 |
| | Sex | -0.276 | 0.047 | -0.266 | 0.000 |
| **TYRO3** | | | | | |
| Unadjusted | S1P | 0.067 | 0.044 | 0.072 | 0.129 |
| Age and sex adjusted | S1P | 0.059 | 0.044 | 0.064 | 0.179 |
| | Age | -0.002 | 0.002 | -0.060 | 0.205 |
| | Sex | -0.046 | 0.026 | -0.082 | 0.086 |
| **CRKL** | | | | | |
| **Unadjusted** | **S1P** | **0.921** | **0.122** | **0.336** | **2.49x10-13** |
| **Age and sex adjusted** | **S1P** | **0.934** | **0.123** | **0.341** | **1.89x10-13** |
| | **Age** | **-0.006** | **0.005** | **-0.052** | **0.240** |
| | **Sex** | **0.033** | **0.074** | **0.020** | **0.654** |
| **IGFBPL1** | | | | | |
| Unadjusted | S1P | 0.138 | 0.064 | 0.101 | 0.032 |
| Age and sex adjusted | S1P | 0.125 | 0.064 | 0.092 | 0.050 |
| | Age | 0.009 | 0.002 | 0.179 | 0.000 |
| | Sex | -0.008 | 0.038 | -0.010 | 0.832 |
| **VCAN** | | | | | |
| Unadjusted | S1P | 0.005 | 0.062 | 0.003 | 0.942 |
| Age and sex adjusted | S1P | -8.684x10-5 | 0.062 | 0.000 | 0.999 |
| | Age | 0.003 | 0.002 | 0.064 | 0.177 |
| | Sex | -0.004 | 0.037 | -0.005 | 0.914 |
| **TSHB** | | | | | |
| Unadjusted | S1P | -0.189 | 0.110 | -0.082 | 0.086 |
| Age and sex adjusted | S1P | -0.191 | 0.111 | -0.082 | 0.087 |
| | Age | 0.001 | 0.004 | 0.015 | 0.760 |
| | Sex | -0.002 | 0.066 | -0.001 | 0.981 |
| **BAG6** | | | | | |
| **Unadjusted** | **S1P** | **0.590** | **0.096** | **0.278** | **2.03x10-9** |
| **Age and sex adjusted** | **S1P** | **0.586** | **0.097** | **0.276** | **3.71x10-9** |
| | **Age** | **0.001** | **0.004** | **0.010** | **0.819** |
| | **Sex** | **-0.016** | **0.058** | **-0.013** | **0.785** |
| **NECTIN2** | | | | | |
| Unadjusted | S1P | 0.178 | 0.068 | 0.124 | 0.009 |

| | | | | | |
|---|---|---|---|---|---|
| Age and sex adjusted | S1P | 0.168 | 0.068 | 0.117 | 0.014 |
| | Age | 0.001 | 0.003 | 0.017 | 0.710 |
| | Sex | -0.044 | 0.041 | -0.051 | 0.279 |
| **USP8** | | | | | |
| **Unadjusted** | **S1P** | **0.585** | **0.122** | **0.232** | **2.22x10-6** |
| **Age and sex adjusted** | **S1P** | **0.577** | **0.123** | **0.229** | **3.80x10-6** |
| | **Age** | **0.005** | **0.005** | **0.051** | **0.293** |
| | **Sex** | **-0.014** | **0.072** | **-0.010** | **0.845** |
| **FKBP4** | | | | | |
| **Unadjusted** | **S1P** | **0.604** | **0.102** | **0.285** | **5.87x10-9** |
| **Age and sex adjusted** | **S1P** | **0.595** | **0.103** | **0.281** | **1.41x10-8** |
| | **Age** | **-0.002** | **0.004** | **-0.029** | **0.545** |
| | **Sex** | **-0.050** | **0.060** | **-0.040** | **0.409** |
| **SDC4** | | | | | |
| **Unadjusted** | **S1P** | **0.861** | **0.104** | **0.364** | **1.44x10-15** |
| **Age and sex adjusted** | **S1P** | **0.853** | **0.105** | **0.361** | **4.05x10-15** |
| | **Age** | **-0.003** | **0.004** | **-0.037** | **0.395** |
| | **Sex** | **-0.060** | **0.063** | **-0.042** | **0.340** |
| **PAG1** | | | | | |
| **Unadjusted** | **S1P** | **0.609** | **0.131** | **0.215** | **4.10x10-6** |
| **Age and sex adjusted** | **S1P** | **0.593** | **0.131** | **0.210** | **7.92x10-6** |
| | **Age** | **0.012** | **0.005** | **0.110** | **0.017** |
| | **Sex** | **-0.007** | **0.078** | **-0.004** | **0.924** |
| **KYAT1** | | | | | |
| **Unadjusted** | **S1P** | **0.706** | **0.101** | **0.312** | **1.26x10-11** |
| **Age and sex adjusted** | **S1P** | **0.680** | **0.102** | **0.301** | **7.18x10-11** |
| | **Age** | **-0.002** | **0.004** | **-0.023** | **0.609** |
| | **Sex** | **-0.142** | **0.061** | **-0.105** | **0.020** |
| **DAB2** | | | | | |
| **Unadjusted** | **S1P** | **1.196** | **0.182** | **0.296** | **1.54x10-10** |
| **Age and sex adjusted** | **S1P** | **1.190** | **0.184** | **0.295** | **2.84x10-10** |
| | **Age** | **0.003** | **0.007** | **0.021** | **0.637** |
| | **Sex** | **-0.013** | **0.110** | **-0.005** | **0.905** |
| **NPDC1** | | | | | |
| Unadjusted | S1P | 0.045 | 0.067 | 0.031 | 0.507 |
| Age and sex adjusted | S1P | 0.034 | 0.068 | 0.024 | 0.617 |
| | Age | 0.005 | 0.003 | 0.090 | 0.056 |
| | Sex | -0.025 | 0.040 | -0.029 | 0.541 |
| **METRNL** | | | | | |
| Unadjusted | S1P | 0.011 | 0.058 | 0.009 | 0.849 |

| | | | | | |
|---|---|---|---|---|---|
| Age and sex adjusted | S1P | 0.012 | 0.058 | 0.010 | 0.831 |
| | Age | 0.005 | 0.002 | 0.112 | 0.018 |
| | Sex | 0.039 | 0.035 | 0.054 | 0.258 |
| **ROR1** | | | | | |
| Unadjusted | S1P | -0.058 | 0.068 | -0.040 | 0.392 |
| Age and sex adjusted | S1P | -0.067 | 0.068 | -0.047 | 0.326 |
| | Age | 0.006 | 0.003 | 0.099 | 0.036 |
| | Sex | -0.012 | 0.041 | -0.014 | 0.772 |

CVD II PANEL

| Model | | Unstandardized Coefficients | | Standardized Coefficients | p |
|---|---|---|---|---|---|
| | | β | Std. error | Beta | |
| **BMP-6** | | | | | |
| **Unadjusted** | **S1P** | **0.415** | **0.096** | **0.200** | **1.99x10-5** |
| **Age and sex adjusted** | **S1P** | **0.400** | **0.096** | **0.193** | **3.82x10-5** |
| | **Age** | **-0.008** | **0.004** | **-0.098** | **0.034** |
| | **Sex** | **-0.125** | **0.058** | **-0.100** | **0.031** |
| **ANG-1** | | | | | |
| **Unadjusted** | **S1P** | **0.638** | **0.103** | **0.282** | **1.17x10-9** |
| **Age and sex adjusted** | **S1P** | **0.649** | **0.103** | **0.287** | **6.45x10-10** |
| | **Age** | **-0.011** | **0.004** | **-0.130** | **0.004** |
| | **Sex** | **-0.014** | **0.061** | **-0.010** | **0.823** |
| **ADM** | | | | | |
| Unadjusted | S1P | 0.301 | 0.122 | 0.123 | 0.014 |
| Age and sex adjusted | S1P | 0.255 | 0.122 | 0.104 | 0.037 |
| | Age | 0.014 | 0.005 | 0.147 | 0.003 |
| | Sex | -0.106 | 0.073 | -0.072 | 0.147 |
| **CD40-L** | | | | | |
| **Unadjusted** | **S1P** | **1.010** | **0.132** | **0.340** | **1.45x10-13** |
| **Age and sex adjusted** | **S1P** | **1.039** | **0.133** | **0.350** | **4.36x10-14** |
| | **Age** | **-0.007** | **0.005** | **-0.063** | **0.159** |
| | **Sex** | **0.107** | **0.080** | **0.060** | **0.180** |
| **PIGF** | | | | | |
| Unadjusted | S1P | 0.142 | 0.063 | 0.107 | 0.024 |
| Age and sex adjusted | S1P | 0.089 | 0.060 | 0.067 | 0.138 |
| | Age | 0.004 | 0.002 | 0.078 | 0.080 |
| | Sex | -0.251 | 0.036 | -0.315 | 0.000 |
| **ADAM-TS13** | | | | | |
| Unadjusted | S1P | 0.041 | 0.054 | 0.036 | 0.444 |
| Age and sex adjusted | S1P | 0.047 | 0.054 | 0.042 | 0.383 |

| | | | | | |
|---|---|---|---|---|---|
| | Age | -0.003 | 0.002 | -0.066 | 0.163 |
| | Sex | 0.014 | 0.032 | 0.020 | 0.675 |
| **Protein BOC** | | | | | |
| Unadjusted | S1P | 0.066 | 0.059 | 0.053 | 0.266 |
| Age and sex adjusted | S1P | 0.050 | 0.058 | 0.040 | 0.391 |
| | Age | -0.005 | 0.002 | -0.111 | 0.017 |
| | Sex | -0.112 | 0.035 | -0.150 | 0.001 |
| **SRC** | | | | | |
| **Unadjusted** | **S1P** | **-0.329** | **0.056** | **-0.267** | **8.99x10-9** |
| **Age and sex adjusted** | **S1P** | **-0.328** | **0.057** | **-0.267** | **1.25x10-8** |
| | **Age** | **-0.003** | **0.002** | **-0.053** | **0.245** |
| | **Sex** | **-0.013** | **0.034** | **-0.017** | **0.707** |
| **IL-1ra** | | | | | |
| Unadjusted | S1P | 0.226 | 0.123 | 0.087 | 0.066 |
| Age and sex adjusted | S1P | 0.203 | 0.123 | 0.078 | 0.101 |
| | Age | 0.008 | 0.005 | 0.076 | 0.107 |
| | Sex | -0.071 | 0.074 | -0.046 | 0.334 |
| **IL6** | | | | | |
| Unadjusted | S1P | 0.404 | 0.117 | 0.163 | 0.001 |
| Age and sex adjusted | S1P | 0.346 | 0.116 | 0.140 | 0.003 |
| | Age | 0.009 | 0.004 | 0.098 | 0.036 |
| | Sex | -0.240 | 0.069 | -0.163 | 0.001 |
| **TNFRSF10A** | | | | | |
| Unadjusted | S1P | 0.028 | 0.046 | 0.029 | 0.552 |
| Age and sex adjusted | S1P | 0.018 | 0.047 | 0.019 | 0.696 |
| | Age | 0.002 | 0.002 | 0.059 | 0.224 |
| | Sex | -0.033 | 0.028 | -0.057 | 0.239 |
| **STK4** | | | | | |
| Unadjusted | S1P | 0.252 | 0.073 | 0.170 | 0.001 |
| Age and sex adjusted | S1P | 0.257 | 0.074 | 0.173 | 0.001 |
| | Age | -6.448x10-5 | 0.003 | -0.001 | 0.982 |
| | Sex | 0.025 | 0.045 | 0.028 | 0.576 |
| **IDUA** | | | | | |
| **Unadjusted** | **S1P** | **0.437** | **0.072** | **0.275** | **3.44x10-9** |
| **Age and sex adjusted** | **S1P** | **0.437** | **0.073** | **0.274** | **5.08x10-9** |
| | **Age** | **-0.001** | **0.003** | **-0.014** | **0.755** |
| | **Sex** | **-0.008** | **0.044** | **-0.009** | **0.848** |
| **TNFRSF11A** | | | | | |
| Unadjusted | S1P | 0.135 | 0.069 | 0.093 | 0.050 |
| Age and sex adjusted | S1P | 0.123 | 0.069 | 0.085 | 0.075 |
| | Age | 0.002 | 0.003 | 0.043 | 0.366 |

| | | | | | |
|---|---|---|---|---|---|
| | Sex | -0.045 | 0.041 | -0.051 | 0.279 |
| **PAR-1** | | | | | |
| **Unadjusted** | **S1P** | **0.442** | **0.068** | **0.295** | **1.93x10-10** |
| **Age and sex adjusted** | **S1P** | **0.444** | **0.068** | **0.296** | **2.38x10-10** |
| | **Age** | **-0.003** | **0.003** | **-0.051** | **0.264** |
| | **Sex** | **-0.010** | **0.041** | **-0.012** | **0.800** |
| **TRAIL-R2** | | | | | |
| Unadjusted | S1P | 0.077 | 0.060 | 0.061 | 0.195 |
| Age and sex adjusted | S1P | 0.041 | 0.058 | 0.032 | 0.484 |
| | Age | 0.010 | 0.002 | 0.208 | 0.000 |
| | Sex | -0.127 | 0.035 | -0.168 | 0.000 |
| **PRSS27** | | | | | |
| Unadjusted | S1P | 0.170 | 0.077 | 0.103 | 0.028 |
| Age and sex adjusted | S1P | 0.172 | 0.078 | 0.104 | 0.028 |
| | Age | -0.005 | 0.003 | -0.086 | 0.070 |
| | Sex | -0.025 | 0.047 | -0.026 | 0.588 |
| **TIE2** | | | | | |
| Unadjusted | S1P | 0.133 | 0.052 | 0.120 | 0.011 |
| Age and sex adjusted | S1P | 0.117 | 0.052 | 0.105 | 0.025 |
| | Age | -0.004 | 0.002 | -0.096 | 0.038 |
| | Sex | -0.109 | 0.031 | -0.164 | 0.000 |
| **TF** | | | | | |
| Unadjusted | S1P | 0.039 | 0.061 | 0.030 | 0.527 |
| Age and sex adjusted | S1P | 0.008 | 0.060 | 0.006 | 0.890 |
| | Age | 0.006 | 0.002 | 0.130 | 0.005 |
| | Sex | -0.117 | 0.036 | -0.152 | 0.001 |
| **IL1RL2** | | | | | |
| Unadjusted | S1P | 0.113 | 0.066 | 0.081 | 0.087 |
| Age and sex adjusted | S1P | 0.108 | 0.066 | 0.077 | 0.106 |
| | Age | 0.003 | 0.003 | 0.050 | 0.291 |
| | Sex | -0.011 | 0.040 | -0.013 | 0.791 |
| **PDGF subunit B** | | | | | |
| **Unadjusted** | **S1P** | **0.565** | **0.073** | **0.345** | **6.02x10-14** |
| **Age and sex adjusted** | **S1P** | **0.572** | **0.073** | **0.349** | **3.01x10-14** |
| | **Age** | **-0.009** | **0.003** | **-0.136** | **0.002** |
| | **Sex** | **-0.017** | **0.044** | **-0.017** | **0.696** |
| **IL27** | | | | | |
| Unadjusted | S1P | 0.015 | 0.059 | 0.012 | 0.803 |
| Age and sex adjusted | S1P | 0.018 | 0.059 | 0.014 | 0.768 |
| | Age | 0.001 | 0.002 | 0.016 | 0.731 |
| | Sex | 0.020 | 0.035 | 0.027 | 0.578 |

| | | | | | |
|---|---|---|---|---|---|
| **IL-17D** | | | | | |
| Unadjusted | S1P | 0.023 | 0.058 | 0.020 | 0.684 |
| Age and sex adjusted | S1P | 0.016 | 0.058 | 0.013 | 0.785 |
| | Age | 0.006 | 0.002 | 0.121 | 0.012 |
| | Sex | -0.004 | 0.035 | -0.005 | 0.913 |
| **CXCL1** | | | | | |
| **Unadjusted** | **S1P** | **0.417** | **0.094** | **0.206** | **1.09x10-5** |
| **Age and sex adjusted** | **S1P** | **0.441** | **0.094** | **0.218** | **3.56x10-6** |
| | **Age** | **0.002** | **0.004** | **0.031** | **0.497** |
| | **Sex** | **0.140** | **0.056** | **0.115** | **0.013** |
| **LOX-1** | | | | | |
| **Unadjusted** | **S1P** | **0.391** | **0.075** | **0.238** | **3.42x10-7** |
| **Age and sex adjusted** | **S1P** | **0.361** | **0.075** | **0.220** | **2.27x10-6** |
| | **Age** | **0.004** | **0.003** | **0.059** | **0.197** |
| | **Sex** | **-0.129** | **0.045** | **-0.131** | **0.004** |
| **Gal-9** | | | | | |
| Unadjusted | S1P | 0.109 | 0.060 | 0.086 | 0.069 |
| Age and sex adjusted | S1P | 0.082 | 0.060 | 0.064 | 0.172 |
| | Age | 0.006 | 0.002 | 0.121 | 0.009 |
| | Sex | -0.107 | 0.036 | -0.140 | 0.003 |
| **GIF** | | | | | |
| Unadjusted | S1P | 0.104 | 0.124 | 0.040 | 0.401 |
| Age and sex adjusted | S1P | 0.062 | 0.124 | 0.024 | 0.618 |
| | Age | 0.015 | 0.005 | 0.144 | 0.002 |
| | Sex | -0.131 | 0.074 | -0.083 | 0.078 |
| **IL-18** | | | | | |
| **Unadjusted** | **S1P** | **0.483** | **0.090** | **0.246** | **1.38x10-7** |
| **Age and sex adjusted** | **S1P** | **0.411** | **0.087** | **0.209** | **2.92x10-6** |
| | **Age** | **0.004** | **0.003** | **0.048** | **0.272** |
| | **Sex** | **-0.353** | **0.052** | **-0.300** | **0.000** |
| **PIgR** | | | | | |
| Unadjusted | S1P | 0.073 | 0.040 | 0.086 | 0.068 |
| Age and sex adjusted | S1P | 0.067 | 0.040 | 0.079 | 0.098 |
| | Age | 0.003 | 0.002 | 0.088 | 0.061 |
| | Sex | -0.015 | 0.024 | -0.030 | 0.521 |
| **RAGE** | | | | | |
| Unadjusted | S1P | -0.027 | 0.058 | -0.022 | 0.640 |
| Age and sex adjusted | S1P | -0.020 | 0.059 | -0.016 | 0.737 |
| | Age | -0.002 | 0.002 | -0.044 | 0.349 |
| | Sex | 0.026 | 0.035 | 0.035 | 0.464 |
| **SOD2** | | | | | |

| | | | | | |
|---|---|---|---|---|---|
| **Unadjusted** | **S1P** | **0.254** | **0.042** | **0.275** | **3.38x10-9** |
| **Age and sex adjusted** | **S1P** | **0.245** | **0.042** | **0.265** | **9.21x10-9** |
| | **Age** | **-0.004** | **0.002** | **-0.115** | **0.011** |
| | **Sex** | **-0.072** | **0.025** | **-0.131** | **0.004** |
| **CTRC** | | | | | |
| Unadjusted | S1P | -0.031 | 0.107 | -0.014 | 0.769 |
| Age and sex adjusted | S1P | -0.010 | 0.108 | -0.005 | 0.924 |
| | Age | 0.003 | 0.004 | 0.040 | 0.403 |
| | Sex | 0.131 | 0.064 | 0.097 | 0.042 |
| **SPON2** | | | | | |
| Unadjusted | S1P | 0.078 | 0.045 | 0.081 | 0.085 |
| Age and sex adjusted | S1P | 0.076 | 0.046 | 0.080 | 0.096 |
| | Age | 0.000 | 0.002 | 0.004 | 0.931 |
| | Sex | -0.008 | 0.027 | -0.014 | 0.771 |
| **GH** | | | | | |
| Unadjusted | S1P | -0.292 | 0.355 | -0.039 | 0.411 |
| Age and sex adjusted | S1P | 0.160 | 0.295 | 0.021 | 0.588 |
| | Age | 0.028 | 0.011 | 0.095 | 0.016 |
| | Sex | 2.509 | 0.177 | 0.559 | 0.000 |
| **FS** | | | | | |
| Unadjusted | S1P | 0.298 | 0.092 | 0.152 | 0.001 |
| Age and sex adjusted | S1P | 0.262 | 0.091 | 0.134 | 0.004 |
| | Age | 0.012 | 0.003 | 0.162 | 0.000 |
| | Sex | -0.111 | 0.054 | -0.094 | 0.043 |
| **GLO1** | | | | | |
| **Unadjusted** | **S1P** | **0.783** | **0.111** | **0.318** | **5.50x10-12** |
| **Age and sex adjusted** | **S1P** | **0.782** | **0.112** | **0.318** | **9.08x10-12** |
| | **Age** | **-0.002** | **0.004** | **-0.023** | **0.608** |
| | **Sex** | **-0.018** | **0.067** | **-0.012** | **0.787** |
| **CD84** | | | | | |
| **Unadjusted** | **S1P** | **0.499** | **0.067** | **0.345** | **6.29x10-13** |
| **Age and sex adjusted** | **S1P** | **0.504** | **0.067** | **0.349** | **4.57x10-13** |
| | **Age** | **-0.005** | **0.003** | **-0.090** | **0.054** |
| | **Sex** | **0.003** | **0.041** | **0.003** | **0.941** |
| **SERPINA12** | | | | | |
| Unadjusted | S1P | -0.185 | 0.171 | -0.054 | 0.280 |
| Age and sex adjusted | S1P | -0.112 | 0.170 | -0.033 | 0.512 |
| | Age | -0.008 | 0.007 | -0.062 | 0.209 |
| | Sex | 0.316 | 0.103 | 0.152 | 0.002 |
| **REN** | | | | | |
| Unadjusted | S1P | 0.275 | 0.121 | 0.107 | 0.024 |

| | | | | | |
|---|---|---|---|---|---|
| Age and sex adjusted | S1P | 0.188 | 0.114 | 0.073 | 0.099 |
| | Age | -0.013 | 0.004 | -0.127 | 0.004 |
| | Sex | -0.526 | 0.068 | -0.341 | 0.000 |
| **DECR1** | | | | | |
| **Unadjusted** | **S1P** | **0.889** | **0.159** | **0.255** | **4.27x10-8** |
| **Age and sex adjusted** | **S1P** | **0.925** | **0.160** | **0.266** | **1.40x10-8** |
| | **Age** | **-0.014** | **0.006** | **-0.104** | **0.024** |
| | **Sex** | **0.099** | **0.096** | **0.048** | **0.301** |
| **MERTK** | | | | | |
| Unadjusted | S1P | 0.150 | 0.073 | 0.098 | 0.039 |
| Age and sex adjusted | S1P | 0.115 | 0.072 | 0.075 | 0.112 |
| | Age | 0.004 | 0.003 | 0.061 | 0.189 |
| | Sex | -0.161 | 0.043 | -0.175 | 0.000 |
| **KIM-1** | | | | | |
| Unadjusted | S1P | 0.462 | 0.123 | 0.176 | 1.84x10-4 |
| Age and sex adjusted | S1P | 0.401 | 0.118 | 0.153 | 0.001 |
| | Age | 0.029 | 0.005 | 0.284 | 0.000 |
| | Sex | -0.139 | 0.071 | -0.088 | 0.050 |
| **THBS2** | | | | | |
| Unadjusted | S1P | 0.088 | 0.048 | 0.086 | 0.070 |
| Age and sex adjusted | S1P | 0.091 | 0.049 | 0.089 | 0.064 |
| | Age | -0.001 | 0.002 | -0.035 | 0.458 |
| | Sex | 0.008 | 0.029 | 0.012 | 0.798 |
| **TM** | | | | | |
| Unadjusted | S1P | 0.192 | 0.062 | 0.145 | 0.002 |
| Age and sex adjusted | S1P | 0.155 | 0.061 | 0.117 | 0.011 |
| | Age | 0.001 | 0.002 | 0.026 | 0.575 |
| | Sex | -0.185 | 0.036 | -0.233 | 0.000 |
| **VSIG2** | | | | | |
| Unadjusted | S1P | 0.061 | 0.058 | 0.052 | 0.294 |
| Age and sex adjusted | S1P | 0.044 | 0.058 | 0.037 | 0.447 |
| | Age | 0.009 | 0.002 | 0.201 | 0.000 |
| | Sex | -0.031 | 0.035 | -0.043 | 0.379 |
| **AMBP** | | | | | |
| Unadjusted | S1P | 0.053 | 0.051 | 0.049 | 0.297 |
| Age and sex adjusted | S1P | 0.039 | 0.051 | 0.036 | 0.449 |
| | Age | 0.001 | 0.002 | 0.027 | 0.564 |
| | Sex | -0.068 | 0.031 | -0.105 | 0.028 |
| **PRELP** | | | | | |
| Unadjusted | S1P | 0.087 | 0.046 | 0.089 | 0.060 |
| Age and sex adjusted | S1P | 0.083 | 0.047 | 0.084 | 0.077 |
| | Age | 0.004 | 0.002 | 0.111 | 0.019 |

| | | | | | |
|---|---|---|---|---|---|
| | Sex | 0.001 | 0.028 | 0.001 | 0.982 |
| **HO-1** | | | | | |
| Unadjusted | S1P | 0.209 | 0.098 | 0.101 | 0.033 |
| Age and sex adjusted | S1P | 0.145 | 0.096 | 0.070 | 0.130 |
| | Age | 0.004 | 0.004 | 0.055 | 0.227 |
| | Sex | -0.303 | 0.057 | -0.243 | 0.000 |
| **XCL1** | | | | | |
| Unadjusted | S1P | 0.144 | 0.117 | 0.058 | 0.218 |
| Age and sex adjusted | S1P | 0.112 | 0.117 | 0.045 | 0.339 |
| | Age | -0.003 | 0.004 | -0.027 | 0.571 |
| | Sex | -0.183 | 0.070 | -0.123 | 0.010 |
| **IL16** | | | | | |
| Unadjusted | S1P | 0.225 | 0.071 | 0.148 | 0.002 |
| Age and sex adjusted | S1P | 0.187 | 0.071 | 0.123 | 0.008 |
| | Age | 0.003 | 0.003 | 0.052 | 0.264 |
| | Sex | -0.176 | 0.042 | -0.193 | 0.000 |
| **SORT1** | | | | | |
| **Unadjusted** | **S1P** | **0.378** | **0.054** | **0.316** | **7.37x10-12** |
| **Age and sex adjusted** | **S1P** | **0.370** | **0.054** | **0.309** | **2.93x10-11** |
| | **Age** | **0.002** | **0.002** | **0.049** | **0.281** |
| | **Sex** | **-0.031** | **0.032** | **-0.043** | **0.338** |
| **CEACAM8** | | | | | |
| Unadjusted | S1P | 0.245 | 0.070 | 0.165 | 0.001 |
| Age and sex adjusted | S1P | 0.222 | 0.070 | 0.149 | 0.002 |
| | Age | -0.001 | 0.003 | -0.018 | 0.697 |
| | Sex | -0.129 | 0.042 | -0.144 | 0.002 |
| **PTX3** | | | | | |
| Unadjusted | S1P | -0.083 | 0.075 | -0.054 | 0.268 |
| Age and sex adjusted | S1P | -0.083 | 0.075 | -0.054 | 0.269 |
| | Age | -0.007 | 0.003 | -0.116 | 0.017 |
| | Sex | -0.043 | 0.045 | -0.046 | 0.341 |
| **PSGL1** | | | | | |
| Unadjusted | S1P | 0.099 | 0.052 | 0.091 | 0.056 |
| Age and sex adjusted | S1P | 0.085 | 0.052 | 0.078 | 0.101 |
| | Age | 0.000 | 0.002 | -0.006 | 0.897 |
| | Sex | -0.073 | 0.031 | -0.112 | 0.019 |
| **CCL17** | | | | | |
| **Unadjusted** | **S1P** | **0.778** | **0.146** | **0.245** | **1.54x10-7** |
| **Age and sex adjusted** | **S1P** | **0.761** | **0.147** | **0.240** | **3.44x10-7** |
| | **Age** | **0.007** | **0.006** | **0.054** | **0.238** |
| | **Sex** | **-0.045** | **0.088** | **-0.024** | **0.611** |

| | | | | | |
|---|---|---|---|---|---|
| **MMP-7** | | | | | |
| Unadjusted | S1P | 0.197 | 0.086 | 0.107 | 0.023 |
| Age and sex adjusted | S1P | 0.207 | 0.085 | 0.113 | 0.016 |
| | Age | 0.011 | 0.003 | 0.160 | 0.001 |
| | Sex | 0.123 | 0.051 | 0.112 | 0.017 |
| **IgG Fc receptor II-b** | | | | | |
| Unadjusted | S1P | 0.164 | 0.094 | 0.088 | 0.082 |
| Age and sex adjusted | S1P | 0.156 | 0.095 | 0.084 | 0.100 |
| | Age | 0.008 | 0.004 | 0.113 | 0.026 |
| | Sex | -0.030 | 0.057 | -0.026 | 0.603 |
| **ITGB1BP2** | | | | | |
| **Unadjusted** | **S1P** | **1.323** | **0.135** | **0.422** | **1.14x10-20** |
| **Age and sex adjusted** | **S1P** | **1.353** | **0.136** | **0.431** | **3.48x10-21** |
| | **Age** | **-0.004** | **0.005** | **-0.030** | **0.491** |
| | **Sex** | **0.128** | **0.081** | **0.068** | **0.117** |
| **DCN** | | | | | |
| Unadjusted | S1P | -0.004 | 0.060 | -0.003 | 0.948 |
| Age and sex adjusted | S1P | -0.023 | 0.060 | -0.018 | 0.703 |
| | Age | 0.004 | 0.002 | 0.086 | 0.069 |
| | Sex | -0.073 | 0.036 | -0.096 | 0.045 |
| **Dkk-1** | | | | | |
| **Unadjusted** | **S1P** | **0.649** | **0.081** | **0.354** | **1.20x10-14** |
| **Age and sex adjusted** | **S1P** | **0.659** | **0.082** | **0.359** | **6.51x10-15** |
| | **Age** | **-0.007** | **0.003** | **-0.100** | **0.025** |
| | **Sex** | **0.008** | **0.049** | **0.008** | **0.864** |
| **LPL** | | | | | |
| Unadjusted | S1P | 0.000 | 0.112 | 0.000 | 0.998 |
| Age and sex adjusted | S1P | 0.067 | 0.109 | 0.029 | 0.537 |
| | Age | 0.004 | 0.004 | 0.038 | 0.403 |
| | Sex | 0.369 | 0.065 | 0.261 | 0.000 |
| **PRSS8** | | | | | |
| Unadjusted | S1P | 0.195 | 0.080 | 0.114 | 0.016 |
| Age and sex adjusted | S1P | 0.123 | 0.077 | 0.072 | 0.108 |
| | Age | 0.007 | 0.003 | 0.102 | 0.022 |
| | Sex | -0.328 | 0.046 | -0.320 | 0.000 |
| **AGRP** | | | | | |
| Unadjusted | S1P | 0.046 | 0.067 | 0.033 | 0.488 |
| Age and sex adjusted | S1P | 0.046 | 0.066 | 0.033 | 0.483 |
| | Age | -0.012 | 0.003 | -0.222 | 0.000 |
| | Sex | -0.077 | 0.039 | -0.091 | 0.050 |
| **HB-EGF** | | | | | |

| | | | | | |
|---|---|---|---|---|---|
| **Unadjusted** | **S1P** | **0.859** | **0.103** | **0.366** | **1.25x10-15** |
| **Age and sex adjusted** | **S1P** | **0.853** | **0.103** | **0.363** | **1.65x10-15** |
| | **Age** | **-0.012** | **0.004** | **-0.127** | **0.004** |
| | **Sex** | **-0.102** | **0.062** | **-0.073** | **0.099** |
| **GDF-2** | | | | | |
| Unadjusted | S1P | 0.003 | 0.107 | 0.001 | 0.981 |
| Age and sex adjusted | S1P | 0.058 | 0.105 | 0.026 | 0.581 |
| | Age | 0.003 | 0.004 | 0.038 | 0.407 |
| | Sex | 0.312 | 0.063 | 0.231 | 0.000 |
| **FABP2** | | | | | |
| Unadjusted | S1P | 0.154 | 0.118 | 0.062 | 0.193 |
| Age and sex adjusted | S1P | 0.122 | 0.119 | 0.049 | 0.305 |
| | Age | 0.005 | 0.005 | 0.054 | 0.257 |
| | Sex | -0.135 | 0.071 | -0.090 | 0.060 |
| **THPO** | | | | | |
| **Unadjusted** | **S1P** | **0.314** | **0.053** | **0.273** | **4.75x10-9** |
| **Age and sex adjusted** | **S1P** | **0.317** | **0.053** | **0.275** | **4.93x10-9** |
| | **Age** | **-0.002** | **0.002** | **-0.042** | **0.365** |
| | **Sex** | **0.001** | **0.032** | **0.001** | **0.975** |
| **MARCO** | | | | | |
| Unadjusted | S1P | 0.158 | 0.048 | 0.152 | 0.001 |
| Age and sex adjusted | S1P | 0.136 | 0.048 | 0.132 | 0.005 |
| | Age | 0.003 | 0.002 | 0.081 | 0.079 |
| | Sex | -0.090 | 0.029 | -0.145 | 0.002 |
| **MMP-12** | | | | | |
| Unadjusted | S1P | -0.055 | 0.106 | -0.025 | 0.601 |
| Age and sex adjusted | S1P | -0.073 | 0.105 | -0.033 | 0.490 |
| | Age | 0.013 | 0.004 | 0.152 | 0.001 |
| | Sex | -0.010 | 0.063 | -0.007 | 0.879 |
| **ACE2** | | | | | |
| Unadjusted | S1P | 0.315 | 0.100 | 0.147 | 0.002 |
| Age and sex adjusted | S1P | 0.222 | 0.095 | 0.104 | 0.020 |
| | Age | 0.012 | 0.004 | 0.145 | 0.001 |
| | Sex | -0.408 | 0.057 | -0.318 | 0.000 |
| **PD-L2** | | | | | |
| Unadjusted | S1P | -0.084 | 0.101 | -0.040 | 0.405 |
| Age and sex adjusted | S1P | -0.084 | 0.101 | -0.040 | 0.406 |
| | Age | -0.004 | 0.004 | -0.054 | 0.255 |
| | Sex | -0.039 | 0.060 | -0.031 | 0.518 |
| **CTSL1** | | | | | |
| Unadjusted | S1P | 0.136 | 0.066 | 0.097 | 0.041 |

| | | | | | |
|---|---|---|---|---|---|
| Age and sex adjusted | S1P | 0.100 | 0.065 | 0.071 | 0.127 |
| | Age | 0.007 | 0.003 | 0.137 | 0.003 |
| | Sex | -0.141 | 0.039 | -0.167 | 0.000 |
| **Hoscar** | | | | | |
| Unadjusted | S1P | 0.098 | 0.053 | 0.087 | 0.066 |
| Age and sex adjusted | S1P | 0.091 | 0.054 | 0.081 | 0.091 |
| | Age | -0.001 | 0.002 | -0.013 | 0.775 |
| | Sex | -0.041 | 0.032 | -0.060 | 0.206 |
| **TNFRSF13B** | | | | | |
| Unadjusted | S1P | 0.058 | 0.077 | 0.036 | 0.453 |
| Age and sex adjusted | S1P | 0.060 | 0.077 | 0.037 | 0.438 |
| | Age | -0.004 | 0.003 | -0.059 | 0.217 |
| | Sex | -0.010 | 0.046 | -0.010 | 0.832 |
| **TGM2** | | | | | |
| Unadjusted | S1P | 0.216 | 0.127 | 0.080 | 0.090 |
| Age and sex adjusted | S1P | 0.171 | 0.127 | 0.064 | 0.179 |
| | Age | 0.005 | 0.005 | 0.051 | 0.277 |
| | Sex | -0.199 | 0.076 | -0.123 | 0.009 |
| **LEP** | | | | | |
| Unadjusted | S1P | 0.147 | 0.172 | 0.040 | 0.396 |
| Age and sex adjusted | S1P | 0.303 | 0.160 | 0.083 | 0.059 |
| | Age | 0.003 | 0.006 | 0.022 | 0.611 |
| | Sex | 0.850 | 0.096 | 0.392 | 0.000 |
| **HSP 27** | | | | | |
| Unadjusted | S1P | -0.175 | 0.061 | -0.136 | 0.004 |
| Age and sex adjusted | S1P | -0.170 | 0.061 | -0.131 | 0.006 |
| | Age | -0.003 | 0.002 | -0.059 | 0.213 |
| | Sex | 0.012 | 0.037 | 0.015 | 0.751 |
| **CD4** | | | | | |
| Unadjusted | S1P | 0.024 | 0.061 | 0.018 | 0.698 |
| Age and sex adjusted | S1P | 0.014 | 0.061 | 0.011 | 0.815 |
| | Age | 0.000 | 0.002 | -0.005 | 0.911 |
| | Sex | -0.049 | 0.037 | -0.064 | 0.179 |
| **NEMO** | | | | | |
| **Unadjusted** | **S1P** | **0.847** | **0.107** | **0.352** | **1.57x10-14** |
| **Age and sex adjusted** | **S1P** | **0.845** | **0.108** | **0.352** | **2.99x10-14** |
| | **Age** | **-0.003** | **0.004** | **-0.030** | **0.501** |
| | **Sex** | **-0.026** | **0.064** | **-0.018** | **0.689** |
| **VEGF-D** | | | | | |
| Unadjusted | S1P | 0.055 | 0.068 | 0.038 | 0.424 |
| Age and sex adjusted | S1P | 0.075 | 0.068 | 0.052 | 0.270 |
| | Age | 0.002 | 0.003 | 0.028 | 0.556 |

| | | | | | |
|---|---|---|---|---|---|
| | Sex | 0.117 | 0.041 | 0.135 | 0.004 |

### CVD III PANEL

| Model | | Unstandardized Coefficients | | Standardized Coefficients | p |
|---|---|---|---|---|---|
| | | β | Std. error | Beta | |
| **TNFRSF14** | | | | | |
| | **S1P** | **0.434** | **0.075** | **0.264** | **1.26x10-8** |
| **Age and sex adjusted** | **S1P** | **0.423** | **0.075** | **0.257** | **3.49x10-8** |
| | **Age** | **0.004** | **0.003** | **0.064** | **0.162** |
| | **Sex** | **-0.029** | **0.045** | **-0.029** | **0.524** |
| **LDL receptor** | | | | | |
| Unadjusted | S1P | 0.320 | 0.113 | 0.132 | 0.005 |
| Age and sex adjusted | S1P | 0.263 | 0.112 | 0.109 | 0.020 |
| | Age | 0.009 | 0.004 | 0.092 | 0.046 |
| | Sex | -0.238 | 0.067 | -0.164 | 0.000 |
| **ITGB2** | | | | | |
| Unadjusted | S1P | 0.050 | 0.096 | 0.025 | 0.601 |
| Age and sex adjusted | S1P | 0.055 | 0.097 | 0.027 | 0.573 |
| | Age | 0.002 | 0.004 | 0.024 | 0.607 |
| | Sex | 0.035 | 0.058 | 0.028 | 0.551 |
| **IL-17RA** | | | | | |
| Unadjusted | S1P | 0.303 | 0.085 | 0.166 | 3.94x10-4 |
| Age and sex adjusted | S1P | 0.280 | 0.085 | 0.153 | 0.001 |
| | Age | 0.000 | 0.003 | -0.004 | 0.927 |
| | Sex | -0.120 | 0.051 | -0.110 | 0.019 |
| **TNF-R2** | | | | | |
| Unadjusted | S1P | 0.145 | 0.080 | 0.086 | 0.069 |
| Age and sex adjusted | S1P | 0.116 | 0.080 | 0.069 | 0.145 |
| | Age | 0.008 | 0.003 | 0.122 | 0.009 |
| | Sex | -0.098 | 0.048 | -0.097 | 0.040 |
| **MMP-9** | | | | | |
| **Unadjusted** | **S1P** | **0.595** | **0.118** | **0.231** | **6.72x10-7** |
| **Age and sex adjusted** | **S1P** | **0.541** | **0.117** | **0.211** | **5.21x10-6** |
| | **Age** | **0.001** | **0.005** | **0.007** | **0.871** |
| | **Sex** | **-0.267** | **0.070** | **-0.174** | **0.000** |
| **IL2-RA** | | | | | |
| Unadjusted | S1P | 0.172 | 0.094 | 0.086 | 0.068 |
| Age and sex adjusted | S1P | 0.146 | 0.095 | 0.073 | 0.125 |
| | Age | 0.006 | 0.004 | 0.077 | 0.101 |
| | Sex | -0.099 | 0.056 | -0.082 | 0.081 |

| | | | | | |
|---|---|---|---|---|---|
| **OPG** | | | | | |
| Unadjusted | S1P | 0.187 | 0.079 | 0.110 | 0.019 |
| Age and sex adjusted | S1P | 0.177 | 0.078 | 0.104 | 0.024 |
| | Age | 0.014 | 0.003 | 0.213 | 0.000 |
| | Sex | 0.039 | 0.047 | 0.038 | 0.407 |
| **ALCAM** | | | | | |
| Unadjusted | S1P | 0.147 | 0.071 | 0.097 | 0.039 |
| Age and sex adjusted | S1P | 0.157 | 0.071 | 0.104 | 0.028 |
| | Age | 0.005 | 0.003 | 0.092 | 0.050 |
| | Sex | 0.084 | 0.042 | 0.093 | 0.048 |
| **TFF3** | | | | | |
| Unadjusted | S1P | -0.080 | 0.129 | -0.029 | 0.536 |
| Age and sex adjusted | S1P | -0.041 | 0.129 | -0.015 | 0.749 |
| | Age | 0.004 | 0.005 | 0.034 | 0.468 |
| | Sex | 0.217 | 0.077 | 0.133 | 0.005 |
| **SELP** | | | | | |
| **Unadjusted** | **S1P** | **0.920** | **0.104** | **0.384** | **2.32x10-17** |
| **Age and sex adjusted** | **S1P** | **0.918** | **0.105** | **0.384** | **5.31x10-17** |
| | **Age** | **0.002** | **0.004** | **0.017** | **0.690** |
| | **Sex** | **0.001** | **0.063** | **0.000** | **0.992** |
| **CSTB** | | | | | |
| **Unadjusted** | **S1P** | **0.556** | **0.103** | **0.247** | **1.01x10-7** |
| **Age and sex adjusted** | **S1P** | **0.549** | **0.103** | **0.244** | **1.67x10-7** |
| | **Age** | **0.009** | **0.004** | **0.109** | **0.017** |
| | **Sex** | **0.019** | **0.062** | **0.014** | **0.754** |
| **CD163** | | | | | |
| Unadjusted | S1P | 0.334 | 0.100 | 0.155 | 0.001 |
| Age and sex adjusted | S1P | 0.290 | 0.100 | 0.135 | 0.004 |
| | Age | 0.012 | 0.004 | 0.139 | 0.003 |
| | Sex | -0.151 | 0.060 | -0.117 | 0.012 |
| **Gal-3** | | | | | |
| Unadjusted | S1P | 0.205 | 0.084 | 0.114 | 0.015 |
| Age and sex adjusted | S1P | 0.216 | 0.084 | 0.120 | 0.010 |
| | Age | 0.010 | 0.003 | 0.140 | 0.003 |
| | Sex | 0.114 | 0.050 | 0.106 | 0.023 |
| **GRN** | | | | | |
| Unadjusted | S1P | 0.227 | 0.075 | 0.141 | 0.003 |
| Age and sex adjusted | S1P | 0.218 | 0.076 | 0.135 | 0.004 |
| | Age | 0.003 | 0.003 | 0.048 | 0.305 |
| | Sex | -0.029 | 0.045 | -0.030 | 0.523 |
| **MEPE** | | | | | |

| | | | | | |
|---|---|---|---|---|---|
| Unadjusted | S1P | 0.236 | 0.097 | 0.114 | 0.016 |
| Age and sex adjusted | S1P | 0.220 | 0.096 | 0.106 | 0.023 |
| | Age | -0.011 | 0.004 | -0.141 | 0.002 |
| | Sex | -0.150 | 0.058 | -0.121 | 0.010 |
| **BLM hydrolase** | | | | | |
| **Unadjusted** | **S1P** | **0.542** | **0.078** | **0.313** | **9.90x10-12** |
| **Age and sex adjusted** | **S1P** | **0.525** | **0.078** | **0.303** | **5.51x10-11** |
| | **Age** | **0.001** | **0.003** | **0.012** | **0.795** |
| | **Sex** | **-0.083** | **0.047** | **-0.080** | **0.075** |
| **PLC** | | | | | |
| Unadjusted | S1P | 0.193 | 0.079 | 0.114 | 0.016 |
| Age and sex adjusted | S1P | 0.169 | 0.079 | 0.100 | 0.034 |
| | Age | 0.008 | 0.003 | 0.122 | 0.009 |
| | Sex | -0.071 | 0.047 | -0.070 | 0.135 |
| **LTBR** | | | | | |
| Unadjusted | S1P | 0.205 | 0.077 | 0.124 | 0.008 |
| Age and sex adjusted | S1P | 0.192 | 0.078 | 0.116 | 0.014 |
| | Age | 0.005 | 0.003 | 0.081 | 0.084 |
| | Sex | -0.034 | 0.046 | -0.034 | 0.468 |
| **Notch 3** | | | | | |
| Unadjusted | S1P | 0.105 | 0.087 | 0.057 | 0.227 |
| Age and sex adjusted | S1P | 0.117 | 0.086 | 0.064 | 0.175 |
| | Age | 0.010 | 0.003 | 0.142 | 0.002 |
| | Sex | 0.124 | 0.052 | 0.113 | 0.016 |
| **TIMP4** | | | | | |
| Unadjusted | S1P | 0.245 | 0.094 | 0.122 | 0.009 |
| Age and sex adjusted | S1P | 0.261 | 0.091 | 0.130 | 0.004 |
| | Age | 0.017 | 0.003 | 0.224 | 0.000 |
| | Sex | 0.188 | 0.054 | 0.157 | 0.001 |
| **CNTN1** | | | | | |
| Unadjusted | S1P | 0.081 | 0.073 | 0.052 | 0.269 |
| Age and sex adjusted | S1P | 0.106 | 0.073 | 0.068 | 0.147 |
| | Age | 0.003 | 0.003 | 0.047 | 0.316 |
| | Sex | 0.144 | 0.043 | 0.156 | 0.001 |
| **CDH5** | | | | | |
| Unadjusted | S1P | 0.121 | 0.094 | 0.060 | 0.200 |
| Age and sex adjusted | S1P | 0.112 | 0.095 | 0.056 | 0.240 |
| | Age | 0.001 | 0.004 | 0.010 | 0.829 |
| | Sex | -0.041 | 0.057 | -0.034 | 0.471 |
| **TLT-2** | | | | | |
| **Unadjusted** | **S1P** | **0.344** | **0.088** | **0.181** | **1.10x10-4** |

| | | | | | |
|---|---|---|---|---|---|
| **Age and sex adjusted** | **S1P** | **0.354** | **0.089** | **0.186** | **8.05x10-5** |
| | **Age** | **0.000** | **0.003** | **0.004** | **0.939** |
| | **Sex** | **0.053** | **0.053** | **0.046** | **0.322** |
| **FABP4** | | | | | |
| Unadjusted | S1P | 0.277 | 0.137 | 0.095 | 0.044 |
| Age and sex adjusted | S1P | 0.302 | 0.134 | 0.103 | 0.025 |
| | Age | 0.022 | 0.005 | 0.199 | 0.000 |
| | Sex | 0.267 | 0.080 | 0.153 | 0.001 |
| **TFPI** | | | | | |
| Unadjusted | S1P | 0.277 | 0.077 | 0.168 | 3.30x10-4 |
| Age and sex adjusted | S1P | 0.240 | 0.076 | 0.146 | 0.002 |
| | Age | 0.007 | 0.003 | 0.106 | 0.021 |
| | Sex | -0.147 | 0.045 | -0.149 | 0.001 |
| **PAI** | | | | | |
| **Unadjusted** | **S1P** | **1.045** | **0.110** | **0.409** | **1.15x10-19** |
| **Age and sex adjusted** | **S1P** | **1.018** | **0.111** | **0.399** | **1.30x10-18** |
| | **Age** | **0.003** | **0.004** | **0.028** | **0.510** |
| | **Sex** | **-0.119** | **0.066** | **-0.078** | **0.072** |
| **CCL24** | | | | | |
| Unadjusted | S1P | 0.330 | 0.163 | 0.095 | 0.044 |
| Age and sex adjusted | S1P | 0.270 | 0.163 | 0.078 | 0.098 |
| | Age | -0.002 | 0.006 | -0.014 | 0.759 |
| | Sex | -0.314 | 0.097 | -0.151 | 0.001 |
| **TR** | | | | | |
| Unadjusted | S1P | 0.033 | 0.101 | 0.015 | 0.744 |
| Age and sex adjusted | S1P | 0.045 | 0.102 | 0.021 | 0.658 |
| | Age | -0.003 | 0.004 | -0.034 | 0.468 |
| | Sex | 0.044 | 0.061 | 0.035 | 0.468 |
| **TNFRSF10C** | | | | | |
| Unadjusted | S1P | 0.300 | 0.103 | 0.136 | 0.004 |
| Age and sex adjusted | S1P | 0.315 | 0.104 | 0.143 | 0.002 |
| | Age | 0.000 | 0.004 | -0.001 | 0.976 |
| | Sex | 0.077 | 0.062 | 0.058 | 0.216 |
| **GDF-15** | | | | | |
| Unadjusted | S1P | 0.328 | 0.103 | 0.149 | 0.001 |
| Age and sex adjusted | S1P | 0.271 | 0.097 | 0.123 | 0.006 |
| | Age | 0.028 | 0.004 | 0.329 | 0.000 |
| | Sex | -0.121 | 0.058 | -0.092 | 0.038 |
| **SELE** | | | | | |
| **Unadjusted** | **S1P** | **0.521** | **0.102** | **0.234** | **5.02x10-7** |
| **Age and sex adjusted** | **S1P** | **0.462** | **0.101** | **0.208** | **5.64x10-6** |

| | | | | | |
|---|---|---|---|---|---|
| | **Age** | **0.001** | **0.004** | **0.016** | **0.713** |
| | **Sex** | **-0.286** | **0.060** | **-0.215** | **0.000** |
| **AZU1** | | | | | |
| **Unadjusted** | **S1P** | **0.404** | **0.086** | **0.217** | **3.13x10-6** |
| **Age and sex adjusted** | **S1P** | **0.369** | **0.085** | **0.199** | **1.99x10-5** |
| | **Age** | **0.002** | **0.003** | **0.033** | **0.475** |
| | **Sex** | **-0.161** | **0.051** | **-0.145** | **0.002** |
| **DLK-1** | | | | | |
| Unadjusted | S1P | -0.014 | 0.118 | -0.005 | 0.907 |
| Age and sex adjusted | S1P | -0.073 | 0.118 | -0.029 | 0.535 |
| | Age | 0.006 | 0.005 | 0.064 | 0.167 |
| | Sex | -0.262 | 0.070 | -0.174 | 0.000 |
| **MPO** | | | | | |
| Unadjusted | S1P | 0.295 | 0.079 | 0.174 | 2.04x10-4 |
| Age and sex adjusted | S1P | 0.288 | 0.080 | 0.169 | 3.34x10-4 |
| | Age | 0.000 | 0.003 | 0.004 | 0.938 |
| | Sex | -0.036 | 0.048 | -0.035 | 0.452 |
| **CXCL16** | | | | | |
| Unadjusted | S1P | 0.149 | 0.074 | 0.094 | 0.046 |
| Age and sex adjusted | S1P | 0.147 | 0.075 | 0.093 | 0.051 |
| | Age | 0.003 | 0.003 | 0.048 | 0.311 |
| | Sex | 0.009 | 0.045 | 0.009 | 0.846 |
| **IL-6RA** | | | | | |
| Unadjusted | S1P | 0.254 | 0.081 | 0.147 | 0.002 |
| Age and sex adjusted | S1P | 0.236 | 0.081 | 0.136 | 0.004 |
| | Age | 0.004 | 0.003 | 0.053 | 0.255 |
| | Sex | -0.070 | 0.048 | -0.067 | 0.151 |
| **RETN** | | | | | |
| Unadjusted | S1P | 0.218 | 0.088 | 0.116 | 0.014 |
| Age and sex adjusted | S1P | 0.237 | 0.088 | 0.126 | 0.008 |
| | Age | 0.002 | 0.003 | 0.029 | 0.533 |
| | Sex | 0.110 | 0.053 | 0.098 | 0.038 |
| **IGFBP-1** | | | | | |
| Unadjusted | S1P | -0.086 | 0.166 | -0.024 | 0.604 |
| Age and sex adjusted | S1P | 0.031 | 0.158 | 0.009 | 0.846 |
| | Age | 0.013 | 0.006 | 0.096 | 0.031 |
| | Sex | 0.673 | 0.095 | 0.319 | 0.000 |
| **CHIT1** | | | | | |
| Unadjusted | S1P | 0.181 | 0.184 | 0.048 | 0.326 |
| Age and sex adjusted | S1P | 0.107 | 0.175 | 0.028 | 0.540 |
| | Age | 0.049 | 0.007 | 0.335 | 0.000 |
| | Sex | -0.142 | 0.101 | -0.064 | 0.162 |

| | | | | | |
|---|---|---|---|---|---|
| **TR-AP** | | | | | |
| **Unadjusted** | **S1P** | **0.399** | **0.087** | **0.211** | **5.94x10-6** |
| **Age and sex adjusted** | **S1P** | **0.332** | **0.084** | **0.176** | **9.46x10-5** |
| | **Age** | **0.009** | **0.003** | **0.126** | **0.005** |
| | **Sex** | **-0.278** | **0.050** | **-0.247** | **0.000** |
| **CCL22** | | | | | |
| Unadjusted | S1P | 0.501 | 0.197 | 0.119 | 0.011 |
| Age and sex adjusted | S1P | 0.457 | 0.198 | 0.109 | 0.021 |
| | Age | -0.003 | 0.008 | -0.018 | 0.706 |
| | Sex | -0.236 | 0.118 | -0.094 | 0.046 |
| **PI3** | | | | | |
| Unadjusted | S1P | 0.259 | 0.096 | 0.126 | 0.007 |
| Age and sex adjusted | S1P | 0.215 | 0.095 | 0.105 | 0.025 |
| | Age | 0.007 | 0.004 | 0.088 | 0.057 |
| | Sex | -0.183 | 0.057 | -0.150 | 0.001 |
| **Ep-CAM** | | | | | |
| Unadjusted | S1P | 0.007 | 0.179 | 0.002 | 0.969 |
| Age and sex adjusted | S1P | 0.030 | 0.181 | 0.008 | 0.869 |
| | Age | 0.005 | 0.007 | 0.034 | 0.478 |
| | Sex | 0.145 | 0.108 | 0.064 | 0.179 |
| **AP-N** | | | | | |
| Unadjusted | S1P | 0.203 | 0.076 | 0.125 | 0.008 |
| Age and sex adjusted | S1P | 0.186 | 0.077 | 0.115 | 0.015 |
| | Age | 0.003 | 0.003 | 0.047 | 0.319 |
| | Sex | -0.067 | 0.046 | -0.069 | 0.144 |
| **AXL** | | | | | |
| Unadjusted | S1P | 0.151 | 0.080 | 0.089 | 0.060 |
| Age and sex adjusted | S1P | 0.115 | 0.080 | 0.067 | 0.150 |
| | Age | 0.003 | 0.003 | 0.042 | 0.372 |
| | Sex | -0.165 | 0.048 | -0.162 | 0.001 |
| **IL-1RT1** | | | | | |
| Unadjusted | S1P | 0.159 | 0.072 | 0.104 | 0.027 |
| Age and sex adjusted | S1P | 0.146 | 0.072 | 0.095 | 0.044 |
| | Age | 0.004 | 0.003 | 0.073 | 0.118 |
| | Sex | -0.040 | 0.043 | -0.043 | 0.359 |
| **MMP-2** | | | | | |
| Unadjusted | S1P | 0.183 | 0.085 | 0.101 | 0.032 |
| Age and sex adjusted | S1P | 0.177 | 0.085 | 0.098 | 0.038 |
| | Age | 0.008 | 0.003 | 0.116 | 0.014 |
| | Sex | 0.021 | 0.051 | 0.020 | 0.675 |
| **FAS** | | | | | |
| Unadjusted | S1P | 0.288 | 0.078 | 0.172 | 2.42x10-4 |

| | | | | | |
|---|---|---|---|---|---|
| Age and sex adjusted | S1P | 0.239 | 0.076 | 0.143 | 0.002 |
| | Age | 0.012 | 0.003 | 0.191 | 0.000 |
| | Sex | -0.169 | 0.045 | -0.169 | 0.000 |
| **MB** | | | | | |
| Unadjusted | S1P | 0.092 | 0.115 | 0.038 | 0.425 |
| Age and sex adjusted | S1P | -0.004 | 0.111 | -0.002 | 0.971 |
| | Age | 0.007 | 0.004 | 0.076 | 0.092 |
| | Sex | -0.443 | 0.066 | -0.303 | 0.000 |
| **TNFSF13B** | | | | | |
| Unadjusted | S1P | 0.255 | 0.084 | 0.142 | 0.003 |
| Age and sex adjusted | S1P | 0.266 | 0.084 | 0.148 | 0.002 |
| | Age | 0.007 | 0.003 | 0.107 | 0.021 |
| | Sex | 0.100 | 0.050 | 0.093 | 0.046 |
| **PRTN3** | | | | | |
| Unadjusted | S1P | 0.276 | 0.094 | 0.138 | 0.003 |
| Age and sex adjusted | S1P | 0.268 | 0.095 | 0.133 | 0.005 |
| | Age | 0.002 | 0.004 | 0.020 | 0.666 |
| | Sex | -0.033 | 0.057 | -0.027 | 0.561 |
| **PCSK9** | | | | | |
| Unadjusted | S1P | -0.020 | 0.099 | -0.010 | 0.841 |
| Age and sex adjusted | S1P | -0.013 | 0.100 | -0.006 | 0.899 |
| | Age | -0.005 | 0.004 | -0.068 | 0.161 |
| | Sex | -0.009 | 0.059 | -0.007 | 0.880 |
| **U-PAR** | | | | | |
| Unadjusted | S1P | 0.295 | 0.081 | 0.170 | 2.94x10-4 |
| Age and sex adjusted | S1P | 0.295 | 0.081 | 0.170 | 2.92x10-4 |
| | Age | 0.009 | 0.003 | 0.135 | 0.004 |
| | Sex | 0.055 | 0.048 | 0.053 | 0.253 |
| **OPN** | | | | | |
| Unadjusted | S1P | 0.349 | 0.098 | 0.165 | 4.33x10-4 |
| Age and sex adjusted | S1P | 0.317 | 0.098 | 0.150 | 0.001 |
| | Age | 0.011 | 0.004 | 0.140 | 0.003 |
| | Sex | -0.091 | 0.059 | -0.072 | 0.122 |
| **CTSD** | | | | | |
| **Unadjusted** | **S1P** | **0.467** | **0.086** | **0.248** | **1.03x10-7** |
| **Age and sex adjusted** | **S1P** | **0.396** | **0.083** | **0.210** | **2.42x10-6** |
| | **Age** | **0.014** | **0.003** | **0.188** | **0.000** |
| | **Sex** | **-0.277** | **0.050** | **-0.245** | **0.000** |
| **PGLYRP1** | | | | | |
| Unadjusted | S1P | 0.309 | 0.092 | 0.156 | 0.001 |
| Age and sex adjusted | S1P | 0.293 | 0.093 | 0.148 | 0.002 |
| | Age | 0.003 | 0.004 | 0.035 | 0.450 |

| | | | | | |
|---|---|---|---|---|---|
| | Sex | -0.062 | 0.056 | -0.053 | 0.261 |
| **CPA1** | | | | | |
| Unadjusted | S1P | 0.100 | 0.121 | 0.039 | 0.407 |
| Age and sex adjusted | S1P | 0.060 | 0.120 | 0.023 | 0.618 |
| | Age | 0.018 | 0.005 | 0.181 | 0.000 |
| | Sex | -0.095 | 0.071 | -0.062 | 0.185 |
| **JAMA** | | | | | |
| **Unadjusted** | **S1P** | **0.911** | **0.105** | **0.380** | **5.64x10-17** |
| **Age and sex adjusted** | **S1P** | **0.928** | **0.105** | **0.387** | **2.83x10-17** |
| | **Age** | **-0.001** | **0.004** | **-0.011** | **0.797** |
| | **Sex** | **0.082** | **0.063** | **0.057** | **0.192** |
| **Gal-4** | | | | | |
| Unadjusted | S1P | 0.236 | 0.099 | 0.112 | 0.018 |
| Age and sex adjusted | S1P | 0.231 | 0.099 | 0.109 | 0.021 |
| | Age | 0.008 | 0.004 | 0.094 | 0.046 |
| | Sex | 0.024 | 0.059 | 0.019 | 0.686 |
| **IL-1RT2** | | | | | |
| Unadjusted | S1P | 0.278 | 0.075 | 0.172 | 2.48x10-4 |
| Age and sex adjusted | S1P | 0.247 | 0.075 | 0.152 | 0.001 |
| | Age | 0.000 | 0.003 | -0.004 | 0.935 |
| | Sex | -0.158 | 0.045 | -0.163 | 0.000 |
| **SHPS-1** | | | | | |
| Unadjusted | S1P | 0.082 | 0.086 | 0.045 | 0.339 |
| Age and sex adjusted | S1P | 0.067 | 0.086 | 0.037 | 0.436 |
| | Age | 0.008 | 0.003 | 0.110 | 0.020 |
| | Sex | -0.028 | 0.051 | -0.026 | 0.586 |
| **CCL15** | | | | | |
| Unadjusted | S1P | 0.267 | 0.102 | 0.123 | 0.009 |
| Age and sex adjusted | S1P | 0.255 | 0.103 | 0.117 | 0.013 |
| | Age | 0.006 | 0.004 | 0.072 | 0.124 |
| | Sex | -0.025 | 0.061 | -0.019 | 0.687 |
| **CASP-3** | | | | | |
| **Unadjusted** | **S1P** | **1.111** | **0.119** | **0.403** | **0.000** |
| **Age and sex adjusted** | **S1P** | **1.120** | **0.120** | **0.406** | **0.000** |
| | **Age** | **0.005** | **0.005** | **0.044** | **0.309** |
| | **Sex** | **0.073** | **0.072** | **0.044** | **0.311** |
| **CPB1** | | | | | |
| Unadjusted | S1P | 0.126 | 0.116 | 0.051 | 0.281 |
| Age and sex adjusted | S1P | 0.081 | 0.115 | 0.033 | 0.483 |
| | Age | 0.018 | 0.004 | 0.185 | 0.000 |
| | Sex | -0.119 | 0.069 | -0.080 | 0.085 |

| | | | | | |
|---|---|---|---|---|---|
| **CHI3L1** | | | | | |
| Unadjusted | S1P | 0.516 | 0.139 | 0.173 | 2.24x10-4 |
| Age and sex adjusted | S1P | 0.443 | 0.135 | 0.148 | 0.001 |
| | Age | 0.027 | 0.005 | 0.236 | 0.000 |
| | Sex | -0.202 | 0.081 | -0.113 | 0.013 |
| **ST2** | | | | | |
| Unadjusted | S1P | 0.322 | 0.095 | 0.157 | 0.001 |
| Age and sex adjusted | S1P | 0.217 | 0.088 | 0.106 | 0.014 |
| | Age | 0.009 | 0.003 | 0.118 | 0.006 |
| | Sex | -0.477 | 0.053 | -0.389 | 0.000 |
| **t-PA** | | | | | |
| Unadjusted | S1P | 0.466 | 0.134 | 0.162 | 0.001 |
| Age and sex adjusted | S1P | 0.360 | 0.128 | 0.125 | 0.005 |
| | Age | 0.023 | 0.005 | 0.206 | 0.000 |
| | Sex | -0.397 | 0.077 | -0.231 | 0.000 |
| **SCGB3A2** | | | | | |
| Unadjusted | S1P | 0.163 | 0.127 | 0.061 | 0.198 |
| Age and sex adjusted | S1P | 0.213 | 0.126 | 0.079 | 0.093 |
| | Age | 0.000 | 0.005 | -0.004 | 0.931 |
| | Sex | 0.248 | 0.075 | 0.154 | 0.001 |
| **EGFR** | | | | | |
| Unadjusted | S1P | 0.136 | 0.061 | 0.104 | 0.027 |
| Age and sex adjusted | S1P | 0.128 | 0.062 | 0.098 | 0.038 |
| | Age | 0.000 | 0.002 | -0.009 | 0.853 |
| | Sex | -0.042 | 0.037 | -0.053 | 0.259 |
| **IGFBP-7** | | | | | |
| Unadjusted | S1P | 0.260 | 0.093 | 0.130 | 0.006 |
| Age and sex adjusted | S1P | 0.213 | 0.092 | 0.107 | 0.022 |
| | Age | 0.012 | 0.004 | 0.152 | 0.001 |
| | Sex | -0.167 | 0.055 | -0.140 | 0.003 |
| **CD93** | | | | | |
| Unadjusted | S1P | 0.119 | 0.076 | 0.073 | 0.121 |
| Age and sex adjusted | S1P | 0.109 | 0.077 | 0.067 | 0.158 |
| | Age | 0.003 | 0.003 | 0.048 | 0.306 |
| | Sex | -0.031 | 0.046 | -0.032 | 0.494 |
| **IL-18BP** | | | | | |
| Unadjusted | S1P | 0.175 | 0.085 | 0.096 | 0.041 |
| Age and sex adjusted | S1P | 0.136 | 0.085 | 0.075 | 0.109 |
| | Age | 0.010 | 0.003 | 0.136 | 0.004 |
| | Sex | -0.139 | 0.051 | -0.128 | 0.006 |
| **COL1A1** | | | | | |
| Unadjusted | S1P | 0.205 | 0.078 | 0.123 | 0.009 |

| | | | | | |
|---|---|---|---|---|---|
| Age and sex adjusted | S1P | 0.218 | 0.079 | 0.131 | 0.006 |
| | Age | -0.002 | 0.003 | -0.034 | 0.462 |
| | Sex | 0.052 | 0.047 | 0.052 | 0.267 |
| **PON3** | | | | | |
| Unadjusted | S1P | 0.065 | 0.113 | 0.027 | 0.566 |
| Age and sex adjusted | S1P | 0.113 | 0.113 | 0.047 | 0.317 |
| | Age | -0.005 | 0.004 | -0.057 | 0.225 |
| | Sex | 0.212 | 0.067 | 0.148 | 0.002 |
| **CTSZ** | | | | | |
| Unadjusted | S1P | 0.333 | 0.085 | 0.181 | 1.14x10-4 |
| Age and sex adjusted | S1P | 0.292 | 0.085 | 0.158 | 0.001 |
| | Age | 0.008 | 0.003 | 0.107 | 0.019 |
| | Sex | -0.159 | 0.051 | -0.144 | 0.002 |
| **MMP-3** | | | | | |
| Unadjusted | S1P | 0.353 | 0.119 | 0.139 | 0.003 |
| Age and sex adjusted | S1P | 0.181 | 0.100 | 0.071 | 0.071 |
| | Age | 0.006 | 0.004 | 0.063 | 0.107 |
| | Sex | -0.834 | 0.060 | -0.550 | 0.000 |
| **RARRES2** | | | | | |
| **Unadjusted** | **S1P** | **0.263** | **0.065** | **0.188** | **5.69x10-5** |
| **Age and sex adjusted** | **S1P** | **0.260** | **0.065** | **0.186** | **6.86x10-5** |
| | **Age** | **0.007** | **0.002** | **0.125** | **0.007** |
| | **Sex** | **0.029** | **0.039** | **0.034** | **0.459** |
| **ICAM-2** | | | | | |
| Unadjusted | S1P | 0.187 | 0.085 | 0.103 | 0.028 |
| Age and sex adjusted | S1P | 0.179 | 0.086 | 0.099 | 0.038 |
| | Age | 0.001 | 0.003 | 0.018 | 0.706 |
| | Sex | -0.035 | 0.051 | -0.032 | 0.494 |
| **KLK6** | | | | | |
| Unadjusted | S1P | 0.171 | 0.083 | 0.097 | 0.039 |
| Age and sex adjusted | S1P | 0.153 | 0.084 | 0.087 | 0.067 |
| | Age | 0.003 | 0.003 | 0.047 | 0.313 |
| | Sex | -0.071 | 0.050 | -0.067 | 0.154 |
| **PDGF subunit A** | | | | | |
| **Unadjusted** | **S1P** | **1.015** | **0.112** | **0.392** | **4.94x10-18** |
| **Age and sex adjusted** | **S1P** | **1.036** | **0.113** | **0.400** | **1.98x10-18** |
| | **Age** | **-0.003** | **0.004** | **-0.032** | **0.456** |
| | **Sex** | **0.089** | **0.068** | **0.058** | **0.186** |
| **TNF-R1** | | | | | |
| Unadjusted | S1P | 0.170 | 0.076 | 0.105 | 0.025 |
| Age and sex adjusted | S1P | 0.146 | 0.076 | 0.090 | 0.055 |

| | | | | | |
|---|---|---|---|---|---|
| | Age | 0.007 | 0.003 | 0.110 | 0.019 |
| | Sex | -0.081 | 0.045 | -0.084 | 0.074 |
| **IGFBP-2** | | | | | |
| Unadjusted | S1P | 0.033 | 0.120 | 0.013 | 0.787 |
| Age and sex adjusted | S1P | 0.052 | 0.118 | 0.020 | 0.660 |
| | Age | 0.018 | 0.005 | 0.185 | 0.000 |
| | Sex | 0.210 | 0.071 | 0.138 | 0.003 |
| **Vwf** | | | | | |
| Unadjusted | S1P | 0.086 | 0.168 | 0.024 | 0.607 |
| Age and sex adjusted | S1P | 0.070 | 0.168 | 0.020 | 0.678 |
| | Age | 0.022 | 0.006 | 0.162 | 0.001 |
| | Sex | 0.052 | 0.100 | 0.024 | 0.604 |
| **PECAM-1** | | | | | |
| **Unadjusted** | **S1P** | **0.798** | **0.089** | **0.389** | **8.57x10-18** |
| **Age and sex adjusted** | **S1P** | **0.786** | **0.090** | **0.383** | **4.43x10-17** |
| | **Age** | **0.002** | **0.003** | **0.025** | **0.569** |
| | **Sex** | **-0.048** | **0.054** | **-0.039** | **0.375** |
| **CCL16** | | | | | |
| Unadjusted | S1P | 0.390 | 0.107 | 0.169 | 3.08x10-4 |
| Age and sex adjusted | S1P | 0.312 | 0.104 | 0.135 | 0.003 |
| | Age | 0.013 | 0.004 | 0.151 | 0.001 |
| | Sex | -0.308 | 0.062 | -0.224 | 0.000 |

**Supplemental Table S5 - List of proteins from four proteomic panels that reveal significant sex-specific associations with plasma S1P.** The association-determining sex is highlighted in pink for differences with single comparisons and orange for differences using Bonferroni-corrected P-values of 0.05/299. For the inflammation panel, we found additional sex-specific associations with panel-specific Bonferroni corrected multiple comparisons (0.05/64 ($P \leq 7.81x10^{-4}$) marked in yellow. N=444.

| Inflammation panel | Male (N=216) | | | Female (N=228) | | |
|---|---|---|---|---|---|---|
| | Pearson's r | β | P | Pearson's r | β | P |
| IL8 | 0.071 | 0.169 | 0.297 | 0.228 | 0.341 | 0.001 |
| uPA | 0.047 | 0.053 | 0.493 | 0.163 | 0.176 | 0.014 |
| FGF-21 | 0.093 | 0.480 | 0.173 | 0.179 | 0.814 | 0.007 |
| IL6 | 0.097 | 0.279 | 0.154 | 0.169 | 0.388 | 0.011 |
| TRAIL | 0.043 | 0.056 | 0.534 | 0.223 | 0.251 | 0.001 |
| CCL4 | 0.106 | 0.256 | 0.122 | 0.119 | 0.255 | 0.073 |
| IL-10RB | 0.135 | 0.186 | 0.047 | 0.071 | 0.073 | 0.288 |
| MCP-2 | 0.149 | 0.369 | 0.028 | 0.201 | 0.462 | 0.002 |
| CCL20 | 0.158 | 0.495 | 0.020 | -0.039 | -0.312 | 0.556 |
| CD244 | 0.160 | 0.235 | 0.019 | 0.245 | 0.308 | $1.84x10^{-4}$ |
| MMP-1 | 0.258 | 0.911 | $1.28x10^{-4}$ | 0.202 | 0.663 | 0.002 |
| CD40 | 0.200 | 0.345 | 0.003 | 0.254 | 0.382 | $1.02x10^{-4}$ |
| TWEAK | 0.039 | 0.050 | 0.571 | 0.246 | 0.283 | $1.78x10^{-4}$ |

| Metabolism panel | Pearson's r | β | P | Pearson's r | β | P |
|---|---|---|---|---|---|---|
| CTSO | 0.156 | 0.215 | 0.022 | 0.075 | 0.098 | 0.253 |
| GRAP2 | 0.326 | 1.620 | $9.22x10^{-7}$ | 0.265 | 1.251 | $4.10x10^{-5}$ |
| LILRA5 | 0.110 | 0.170 | 0.106 | 0.159 | 0.245 | 0.015 |
| SNAP23 | 0.330 | 1.111 | $6.49x10^{-7}$ | 0.264 | 0.862 | $4.40x10^{-5}$ |
| NOMO1 | 0.163 | 0.251 | 0.016 | 0.098 | 0.137 | 0.136 |
| ENPP7 | 0.218 | 0.657 | 0.001 | 0.112 | 0.337 | 0.088 |
| CDH2 | 0.144 | 0.256 | 0.035 | 0.069 | 0.108 | 0.300 |
| IGFBPL1 | 0.154 | 0.231 | 0.023 | 0.040 | 0.050 | 0.541 |
| SUMF2 | 0.284 | 0.654 | $2.10x10^{-5}$ | 0.196 | 0.435 | 0.003 |
| ENO2 | 0.250 | 0.601 | $1.97x10^{-4}$ | 0.316 | 0.677 | $7.97x10^{-7}$ |
| **CVDII panel** | **Pearson's r** | **β** | **P** | **Pearson's r** | **β** | **P** |
| IL6 | 0.106 | 0.298 | 0.122 | 0.199 | 0.414 | 0.003 |
| TNFRSF11A | 0.033 | 0.046 | 0.630 | 0.132 | 0.201 | 0.044 |
| PAR-1 | 0.256 | 0.373 | $1.36x10^{-4}$ | 0.323 | 0.503 | $5.13x10^{-7}$ |
| TIE2 | 0.040 | 0.039 | 0.561 | 0.149 | 0.179 | 0.023 |
| FGF-21 | 0.088 | 0.506 | 0.197 | 0.189 | 0.948 | 0.004 |
| FS | 0.117 | 0.211 | 0.086 | 0.162 | 0.340 | 0.014 |
| KIM-1 | 0.220 | 0.583 | 0.001 | 0.115 | 0.301 | 0.081 |
| TM | 0.061 | 0.067 | 0.373 | 0.166 | 0.240 | 0.012 |
| PRELP | 0.035 | 0.033 | 0.609 | 0.136 | 0.138 | 0.039 |
| IL16 | 0.123 | 0.161 | 0.071 | 0.132 | 0.219 | 0.045 |
| MMP-7 | 0.096 | 0.180 | 0.158 | 0.144 | 0.259 | 0.028 |
| MARCO | 0.125 | 0.127 | 0.067 | 0.147 | 0.153 | 0.026 |
| LOX-1 | 0.194 | 0.305 | 0.004 | 0.250 | 0.423 | $1.21x10^{-4}$ |
| IL18 | 0.157 | 0.283 | 0.021 | 0.278 | 0.538 | $1.80x10^{-5}$ |
| GLO1 | 0.241 | 0.619 | $3.41x10^{-4}$ | 0.389 | 0.929 | $8.93x10^{-10}$ |
| **CVDIII panel** | **Pearson's r** | **β** | **P** | **Pearson's r** | **β** | **P** |
| IL-17RA | 0.225 | 0.421 | 0.001 | 0.083 | 0.148 | 0.204 |
| OPG | 0.204 | 0.358 | 0.003 | 0.027 | 0.044 | 0.686 |
| ALCAM | 0.143 | 0.225 | 0.035 | 0.073 | 0.107 | 0.266 |
| CD163 | 0.176 | 0.389 | 0.009 | 0.108 | 0.227 | 0.098 |
| GRN | 0.199 | 0.336 | 0.003 | 0.074 | 0.115 | 0.260 |
| LTBR | 0.157 | 0.267 | 0.021 | 0.084 | 0.136 | 0.201 |
| TNFRSF10C | 0.116 | 0.253 | 0.088 | 0.166 | 0.371 | 0.011 |
| GDF-15 | 0.187 | 0.443 | 0.006 | 0.088 | 0.179 | 0.180 |
| AZU1 | 0.181 | 0.359 | 0.007 | 0.225 | 0.383 | 0.001 |
| CXCL16 | 0.142 | 0.242 | 0.037 | 0.044 | 0.066 | 0.498 |
| RETN | 0.083 | 0.164 | 0.223 | 0.172 | 0.311 | 0.008 |
| PI3 | 0.081 | 0.170 | 0.235 | 0.138 | 0.273 | 0.034 |
| AP-N | 0.162 | 0.260 | 0.017 | 0.075 | 0.125 | 0.249 |
| IL-1RT1 | 0.151 | 0.238 | 0.026 | 0.048 | 0.072 | 0.468 |
| FAS | 0.214 | 0.381 | 0.001 | 0.090 | 0.139 | 0.169 |

| | | | | | | |
|---|---|---|---|---|---|---|
| **PRTN3** | 0.101 | 0.220 | 0.137 | 0.179 | 0.169 | 0.009 |
| **PGLYRP1** | 0.130 | 0.141 | 0.043 | 0.172 | 0.305 | 0.013 |
| **Gal-4** | 0.168 | 0.389 | 0.010 | 0.067 | 0.103 | 0.440 |
| **CCL15** | 0.144 | 0.312 | 0.039 | 0.103 | 0.218 | 0.126 |
| **ST2** | 0.176 | 0.373 | 0.007 | 0.061 | 0.094 | 0.409 |
| **IGFBP-7** | 0.144 | 0.313 | 0.023 | 0.097 | 0.149 | 0.244 |
| **IL-18BP** | 0.132 | 0.263 | 0.035 | 0.045 | 0.042 | 0.722 |
| **COL1A1** | 0.106 | 0.195 | 0.086 | 0.152 | 0.234 | 0.033 |
| **CTSZ** | 0.213 | 0.435 | 0.001 | 0.113 | 0.178 | 0.123 |
| **MMP-3** | 0.158 | 0.373 | 0.012 | -0.001 | 0.017 | 0.899 |
| **ICAM-2** | 0.154 | 0.302 | 0.021 | 0.050 | 0.067 | 0.553 |
| **CTSD** | 0.276 | 0.567 | $4.0x10^{-5}$ | 0.189 | 0.271 | 0.009 |
